# Which children and young people are at higher risk of severe disease and death after SARS-CoV-2 infection: a systematic review and individual patient meta-analysis

**DOI:** 10.1101/2021.06.30.21259763

**Authors:** R Harwood, H Yan, N Talawila Da Camara, C Smith, J Ward, C Tudur-Smith, M Linney, M Clark, E Whittaker, D Saatci, PJ Davis, K Luyt, ES Draper, S Kenny, L K Fraser, R.M Viner

## Abstract

**Background:** We aimed to use individual patient data to describe pre-existing factors associated with severe disease, primarily admission to critical care, and death secondary to SARS-CoV-2 infection in children and young people (CYP) in hospital.

**Methods:** We searched Pubmed, European PMC, Medline and Embase for case series and cohort studies that included all CYP admitted to hospital with ≥30 CYP with SARS-CoV-2 or ≥5 CYP with PIMS-TS or MIS-C. Eligible studies contained 1) details of age, sex, ethnicity or co-morbidities, and 2) an outcome which included admission to critical care, mechanical invasive ventilation, cardiovascular support, or death. Studies reporting outcomes in more restricted grouping of co-morbidities were eligible for narrative review. Authors of eligible studies were approached for individual patient data (IPD). We used random effects meta-analyses for aggregate study-level data and multilevel mixed effect models for IPD data to examine risk factors (age, sex, comorbidities) associated with admission to critical care and death. Data shown are odds ratios and 95% confidence intervals (CI).

**Findings:** 81 studies were included, 57 in the meta-analysis (of which 22 provided IPD) and 26 in the narrative synthesis. Most studies had an element of bias in their design or reporting. Sex was not associated with critical care or death. Compared with CYP aged 1-4 years, infants had increased odds of admission to critical care (OR 1.63 (95% CI 1.40-1.90)) and death (OR 2.08 (1.57-2.86)). Odds of death were increased amongst CYP over 10 years (10-14 years OR 2.15 (1.54-2.98); >14 years OR 2.15 (1.61-2.88)).

Number of comorbid conditions was associated with increased odds of admission to critical care and death for COVID-19 in a dose-related fashion. For critical care admission odds ratios were: 1 comorbidity 1.49 (1.45-1.53); 2 comorbidities 2.58 (2.41-2.75); ≥3 comorbidities 2.97 (2.04-4.32), and for death: 1 comorbidity 2.15 (1.98-2.34); 2 comorbidities 4.63 (4.54-4.74); ≥3 co-morbidities 4.98 (3.78-6.65). Odds of admission to critical care were increased for all co-morbidities apart from asthma (0.92 (0.91-0.94)) and malignancy (0.85 (0.17-4.21)) with an increased odds of death in all co-morbidities considered apart from asthma. Neurological and cardiac comorbidities were associated with the greatest increase in odds of severe disease or death. Obesity increased the odds of severe disease and death independently of other comorbidities.

**Interpretation:** Hospitalised CYP at greatest vulnerability of severe disease or death from SARS-CoV-2 infection are infants, teenagers, those with cardiac or neurological conditions, or 2 or more comorbid conditions, and those who are obese. These groups should be considered higher priority for vaccination and for protective shielding when appropriate. Whilst odds ratios were high, the absolute increase in risk for most comorbidities was small compared to children without underlying conditions.

**Funding:** RH is in receipt of a funded fellowship from Kidney Research UK. JW is in receipt of a Medical Research Council Fellowship.

**Putting Research Into Context:** *Evidence before this study:* The risk factors for severe disease following SARS-CoV-2 infection in adults has been extensively studied and reported, with good evidence that increasing age, non-white ethnicity, male gender and co-morbidities increase the risk. SARS-CoV-2 infection in children and young people (CYP) infrequently results in hospital admission and very rarely causes severe disease and death, making it difficult to discern the impact of a range of potential risk factors for severe disease in the many small to moderate sized published studies. More recent larger publications have aimed to address this question in specific populations but the global experience has not been described. We searched Pubmed, European PMC, Medline and Embase from the 1^st^ January 2020 to 21^st^ May 2021 for case series and cohort studies that included all CYP admitted to hospital with 30 children with reverse transcriptase-PCR confirmed SARS-CoV-2 or 5 CYP defined as having PIMS-TS or MIS-C. 57 studies met the eligibility criteria for meta-analysis.

*Added value of this study:* To our knowledge, this is the first meta-analysis to use individual patient data to compare the odds and risk of critical care admission and death in CYP with COVID-19 and PIMS-TS. We find that the odds of severe disease in hospitalised children is increased in those with multiple co-morbidities, cardiac and neurological co-morbidities and those who are obese. However, the additional risk compared to children without co-morbidity is small.

*Implications of all the available evidence:* Severe COVID-19 and PIMS-TS, whilst rare, can occur in CYP. We have identified pre-existing risk factors for severe disease after SARS-CoV-2 and recommend that those with co-orbidities which place them in the highest risk groups are prioritised for vaccination.

## Introduction

Children and young people (CYP) have suffered fewer direct effects of the COVID-19 pandemic than adults, as the vast majority of CYP experience few if any symptoms of SARS-CoV-2 infection.(1) However a small minority experience more severe disease (2) and small numbers of deaths have been documented.(3) As severe outcomes amongst CYP are uncommon, our understanding of which are at risk from SARS-CoV-2 is limited, in contrast to adults. Yet identification of CYP at the highest risk from infection and its sequelae is essential for guiding clinicians, families and policymakers about who can safely attend school, and for identifying groups to be prioritised for vaccination.

SARS-CoV-2 infection in CYP has two primary manifestations. The first is acute COVID-19 disease, an acute illness caused by current infection with the virus and often characterised by respiratory symptoms. The second is a delayed inflammatory condition referred to as Paediatric Inflammatory Multisystem Syndrome Temporally associated with SARS-CoV-2 (PIMS-TS) or Multisystem Inflammatory Syndrome in Children (MIS-C)(4–6). Postulated risk factors for developing more severe COVID-19 or PIMS-TS / MIS-C include existing co-morbid conditions, age, sex, ethnicity, socio-economic group, and geographical location (7–10). Existing systematic evaluations are not useful for guiding policy as reviews were undertaken early in the pandemic (11–13), included highly heterogeneous groups and a wide range of outcomes from very small studies(14), and failed to distinguish between acute COVID-19 and PIMS-TS/MIS-C. Rapid growth in the literature over the past year provides an opportunity to synthesize findings, and better inform policy decisions about vaccination and protective shielding of vulnerable CYP.

We undertook a high-quality systematic review and meta-analysis of the literature from the first pandemic year to identify which CYP were at increased risk of severe disease or death in CYP admitted to hospital with SARS-CoV-2 infection or PIMS-TS / MIS-C. The study was limited to hospitalised CYP to enable the baseline denominator characteristics to be more accurately defined, particularly co-morbidities, and because in itself, hospital admission may be an indicator of severity. We limited our review to pre-specified potential risk-factors (co-morbidities, age, sex, ethnicity and deprivation), plus a limited number of outcomes denoting severe disease (critical care admission, need for mechanical invasive ventilation or cardiovascular support) and death. These are relevant to policy on protective shielding and school attendance, and potential vaccination strategies for CYP.

## Methods

The protocol for this systematic review and meta-analysis was published on PROSPERO (CRD42021235338) on the 5^th^ February, 2021. We report findings according to the PRISMA 2020 guidelines(15) (Supplementary information 1).

### Search

We performed a systematic search of four major databases: PubMed, European PMC, Scopus and Embase for relevant studies on COVID-19 in children and young people up to 21 years of age, published between the 1^st^ January 2020 and the 29^th^ January 2021 and updated the search on the 21^st^ May 2021. Searches were limited to English only and included key search concepts relating to COVID-19 OR SARS-CoV-2 OR PIMS-TS OR MIS-C AND Child OR Young person OR neonate (full search strategy in supplementary information 1). References of published systematic reviews and included studies were checked for additional studies.

### Study Selection

Two reviewers selected studies using a two-stage process. All titles and abstracts were reviewed independently in duplicate by a team of five reviewers to determine eligibility. Full texts of articles were reviewed if inclusion was not clear in the abstract. Disagreements were discussed between the two reviewers and a decision made about inclusion or exclusion of the study. We excluded studies if the data were duplicated elsewhere, as reported by the study authors, and prioritised the studies which gave comparative data on the risk factors and outcomes of interest; if both did so, we used the larger study.

Inclusion criteria were as follows:

1. Observational studies of any type of CYP under 21 years of age who had been admitted to hospital with a finding of SARS-CoV-2 infection at or during admission by reverse transcriptase polymerase chain reaction (RT-PCR) OR who had been identified clinically as having PIMS-TS or MIS-C. Studies were eligible from institutions or from defined populations.
2. Data were provided on any of the following potential risk factors: age, sex, ethnicity, co-morbidity and socioeconomic deprivation.
3. Studies that included all admitted CYP in a population or institution regardless of co-morbidity were eligible for inclusion in the meta-analysis if they included 30 children with COVID-19 or 5 children with PIMS-TS or MIS-C. 30 children with COVID-19 was selected as the minimum a-priori to account for the outcomes of admission to critical care and death being rare, with previous systematic reviews suggesting severe COVID-19 occurs in approximately 2.5% of children(16). Studies of a single pre-existing co-morbidity were included in the systematic review if they included 5 children but not included in the meta-analysis.
4. Studies which reported one of the following outcomes as a proxy for severe disease:
  1. Need for invasive ventilation during hospital stay (not including during anaesthesia for surgical procedures)
  2. Need for cardiovascular support (vasopressors, inotropes +/− ECMO)
  3. Need for critical/intensive care
  4. Death after diagnosis of SARS-CoV-2 infection or PIMS-TS/MIS-C. We initially intended to include other identifiers indicative of severe disease including use of pharmacological therapy and length of stay in critical care, but discarded these during the search as they were rarely and inconsistently reported. In analyses, CYP who did not have an indicator of severe disease but had COVID-19 or PIMS-TS/MIS-C and were admitted to hospital were used as the comparator group.

### Data Extraction for meta-analysis and Study Quality Assessment

Data on risk factors and outcome variables were extracted from individual studies by one reviewer using a pre-designed data collection form and extraction was cross-checked by a second reviewer in 10% of studies. Authors of studies from the first search (to January 2021) were contacted by email and asked to provide either additional aggregated data demonstrating the relationship between predictor and outcome variables or IPD. Time did not allow these to be requested for studies identified in the second search (to May 2021). IPD were shared by authors using a standardised data collection form and checked for consistency with the original publication. Any queries from sharing authors or the study team were discussed over email or by a virtual video call. Eligible studies not supplying individual patient data in a way that enabled the relationship between risk factors and outcomes to be analysed or that did not provide aggregate or individual patient data were excluded from the meta-analysis.

### Assessment of Bias

We assessed the studies for bias using the Newcastle-Ottawa Scale (17) to assess the quality of observational studies. Studies were scored according to selection of participants, comparability, and outcome. The description of comparator cohorts was deemed present when analyses comparing two groups of outcomes were described within the publication.

### Analysis

Meta-analyses were undertaken separately for COVID-19 and PIMS-TS/MIS-C to examine the association of each clinical outcome with sex (female sex was the reference group), age-group (1-4 years as reference group) and comorbidities (children without any comorbidity were the reference group). Children who were RT-PCR positive for SARS-CoV-2 but met the criteria for PIMS-TS or MIS-C were included in the latter group.

Meta-analyses were conducted in two ways. First, we undertook a random-effects meta-analysis of reported study-level data using RevMan 5 software (18) to estimate pooled odds-ratios for each outcome (death, intensive care admission, mechanical invasive ventilation and cardiovascular support). We refer to this analysis as the aggregate meta-analysis. Age categories were described as <1 year, 1-4 years, 5-9 years, 10-14 years and over 15 years and age 1-4 years used as the reference group. When studies reported a different age grouping, the group was used in the range which had the greatest cross-over of years. Co-morbidity data were compared using the presence and absence of individual co-morbidities. We calculated the I^2^ statistic as a measure of heterogeneity and report prediction intervals. Funnel plots were examined to assess the evidence for publication bias. We then performed a sensitivity analysis by excluding the largest study of patients with COVID-19. The second set of meta-analyses were undertaken on the IPD, using multi-level logistic mixed-effects models in Stata 16 (StataCorp. College Station, TX) including a random effect for study, with models for co-morbidities adjusted for age and sex. After each model we calculated the predicted probability for each outcome amongst those with and without each comorbidity using the margins post estimation command. We did this to estimate risk difference for admission to critical care or death among CYP with comorbidities compared to those without. As a sensitivity analysis, a two-stage meta-analysis was conducted using study-level estimates calculated from the IPD data. A further sensitivity analysis for both the aggregate and IPD meta-analyses was performed by excluding one very large study.(19) Eligible studies which included only CYP with specific comorbidities were not included in the meta-analyses but included in a narrative synthesis. Data displayed are odds ratio (95% confidence interval) and absolute risk difference (95% confidence interval).

## Results

Figure 1 shows the search flow. 23,050 reports were identified. After excluding duplicates and ineligible studies, 81 studies were included in the review. 57 studies were included in the meta-analysis, including a total of 21,412 children (see Table 1). Nine studies were from Asia, fifteen from Europe, one from Africa, twenty-one from North America and eight from South America. One study had global recruitment.

**Table 1.**
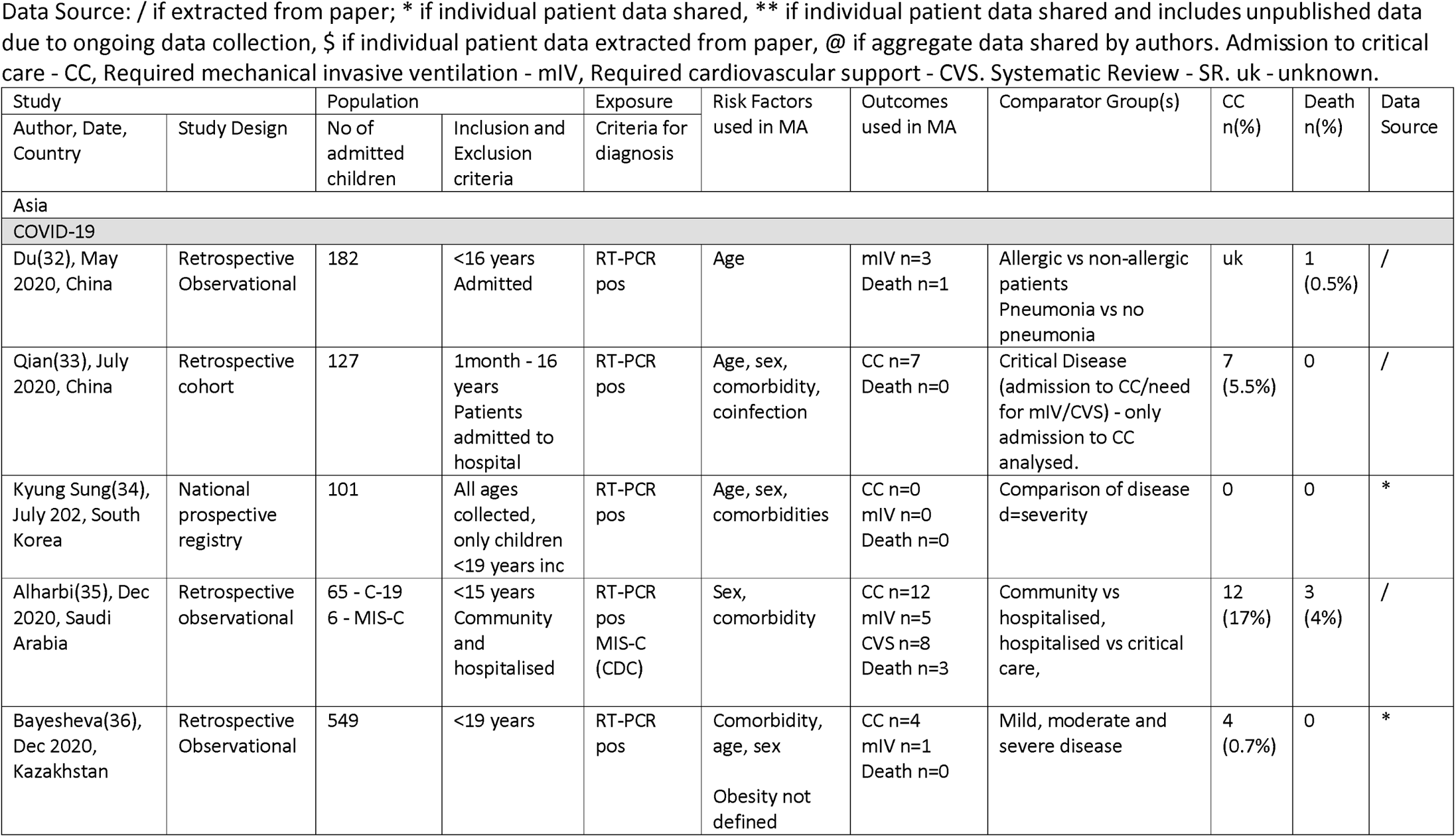
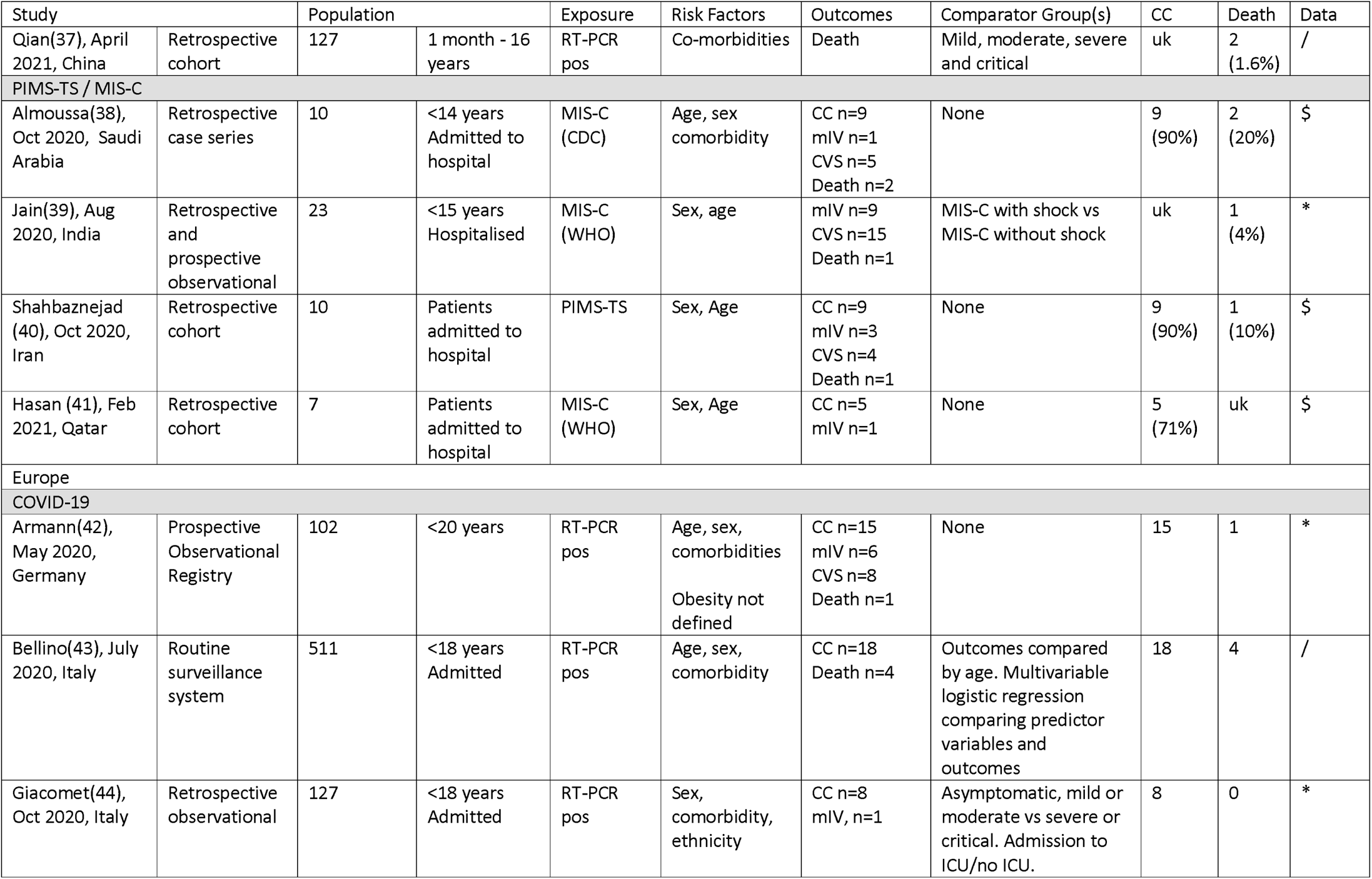
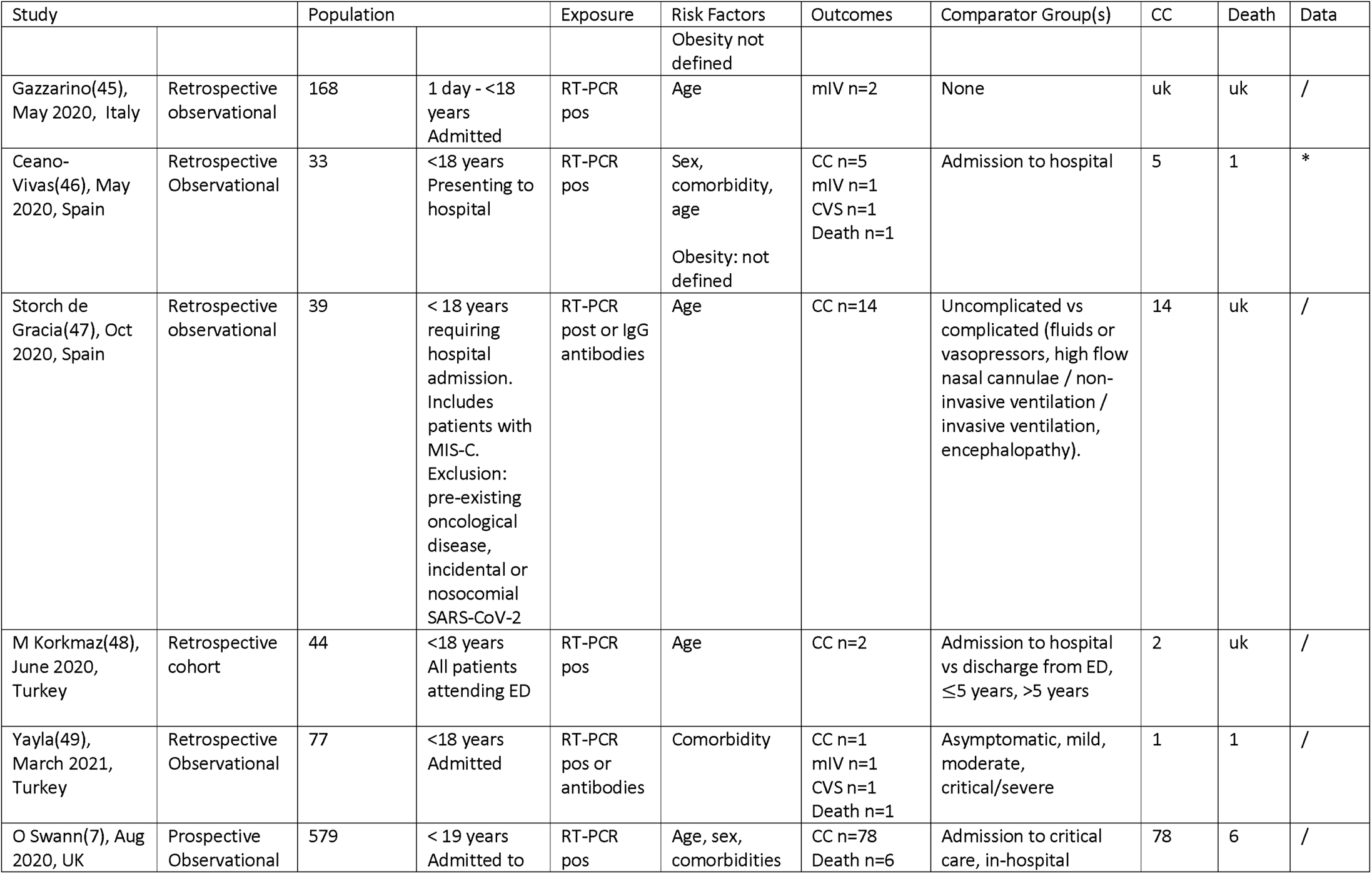
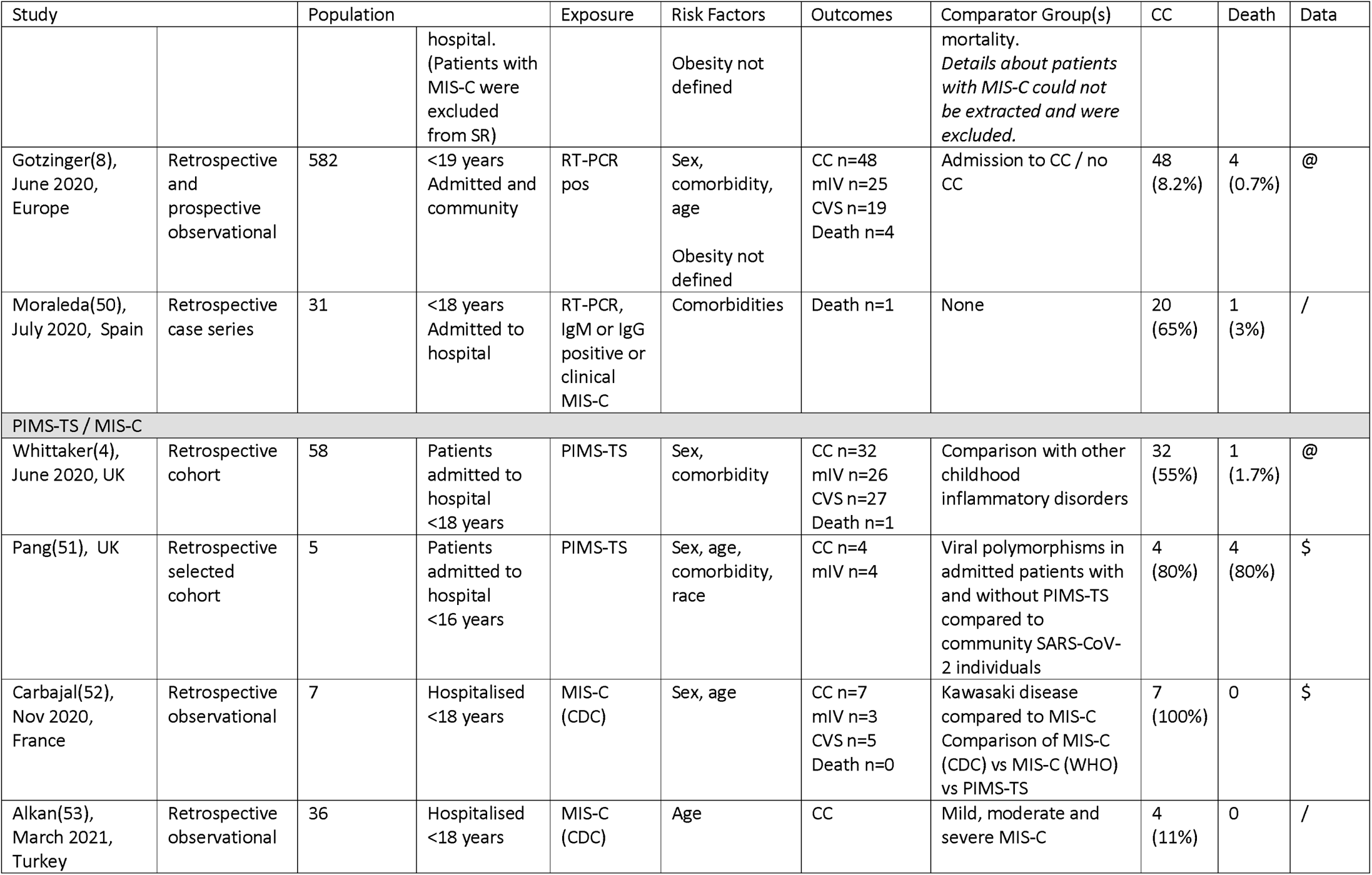
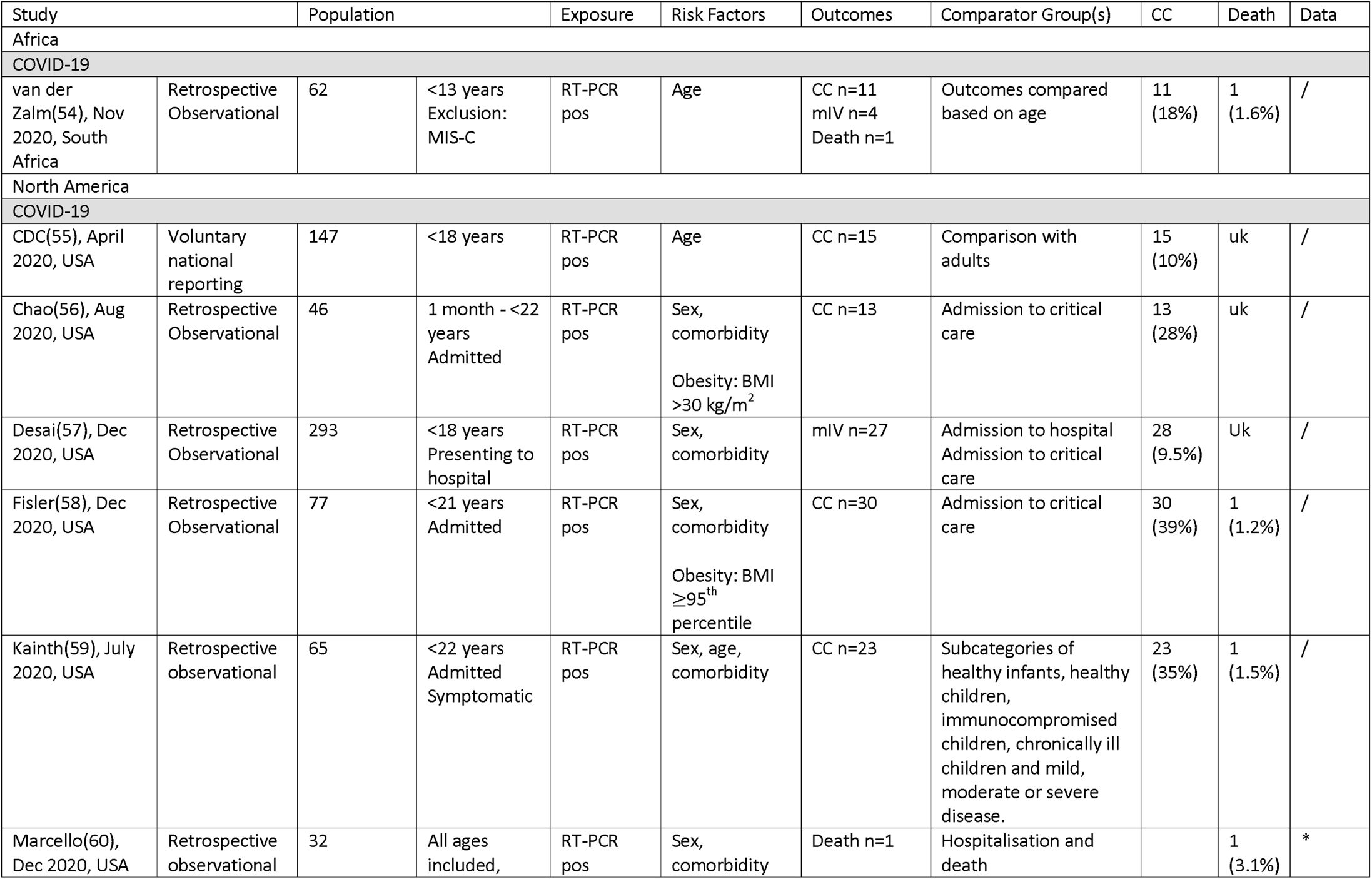
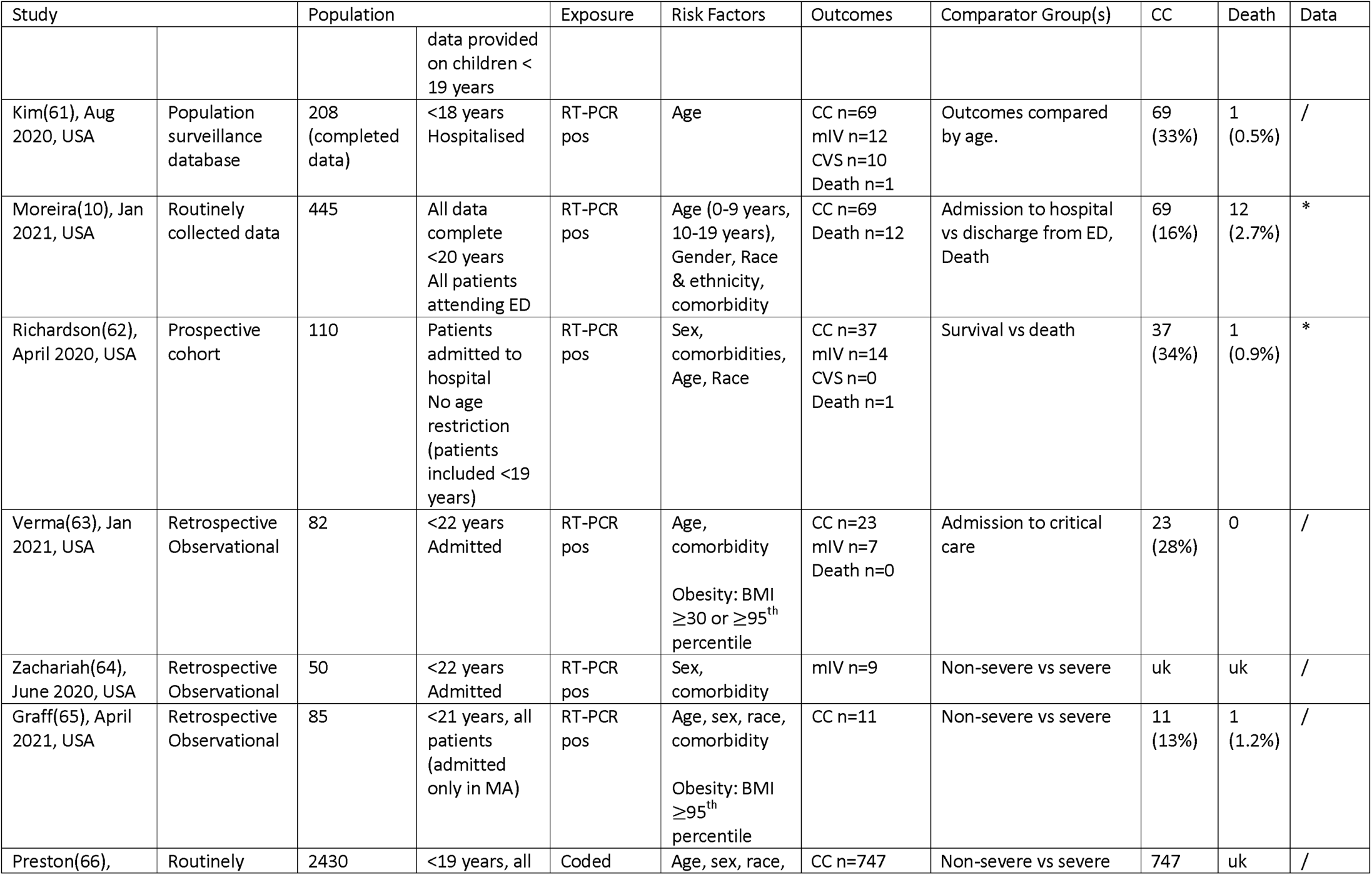
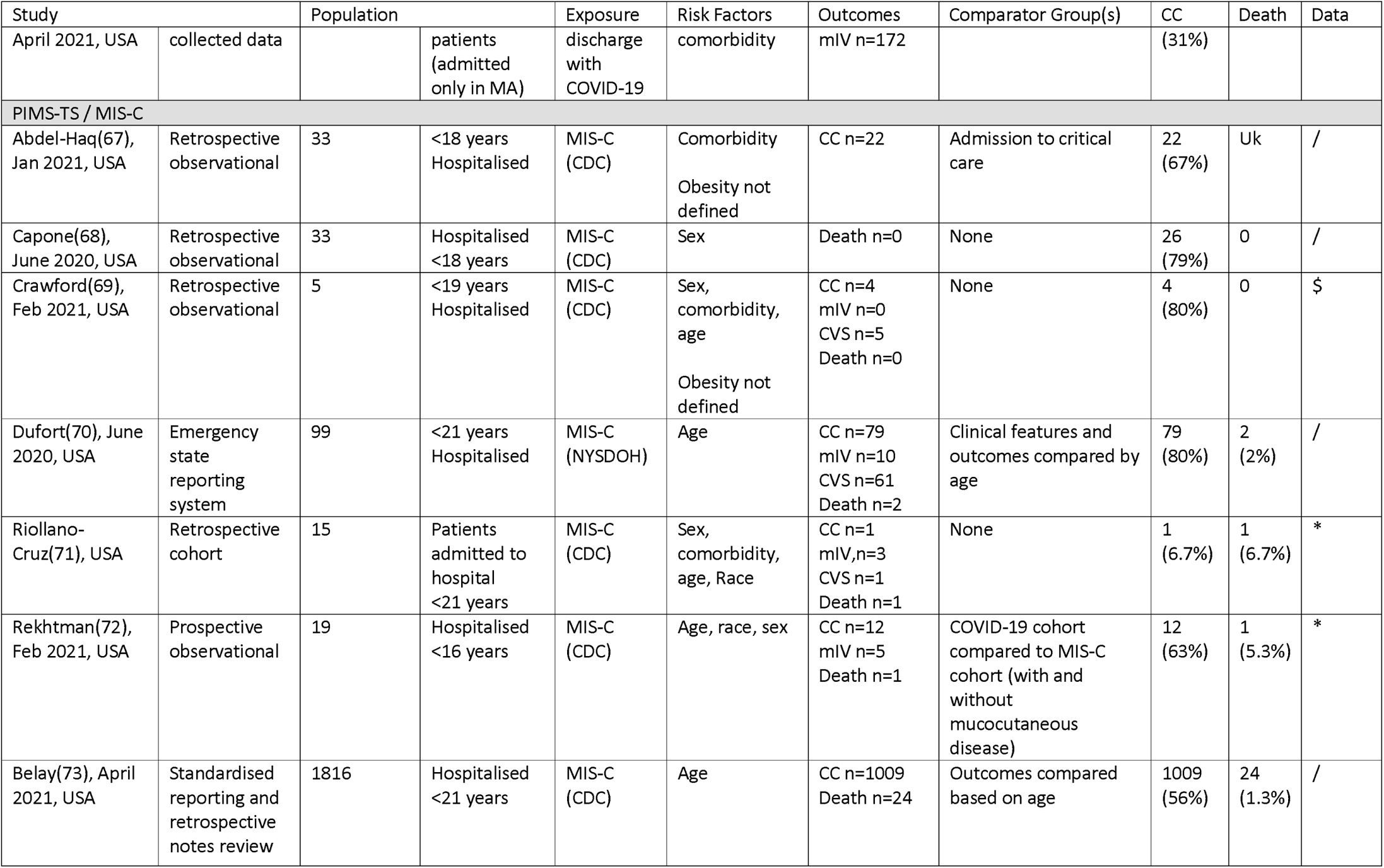
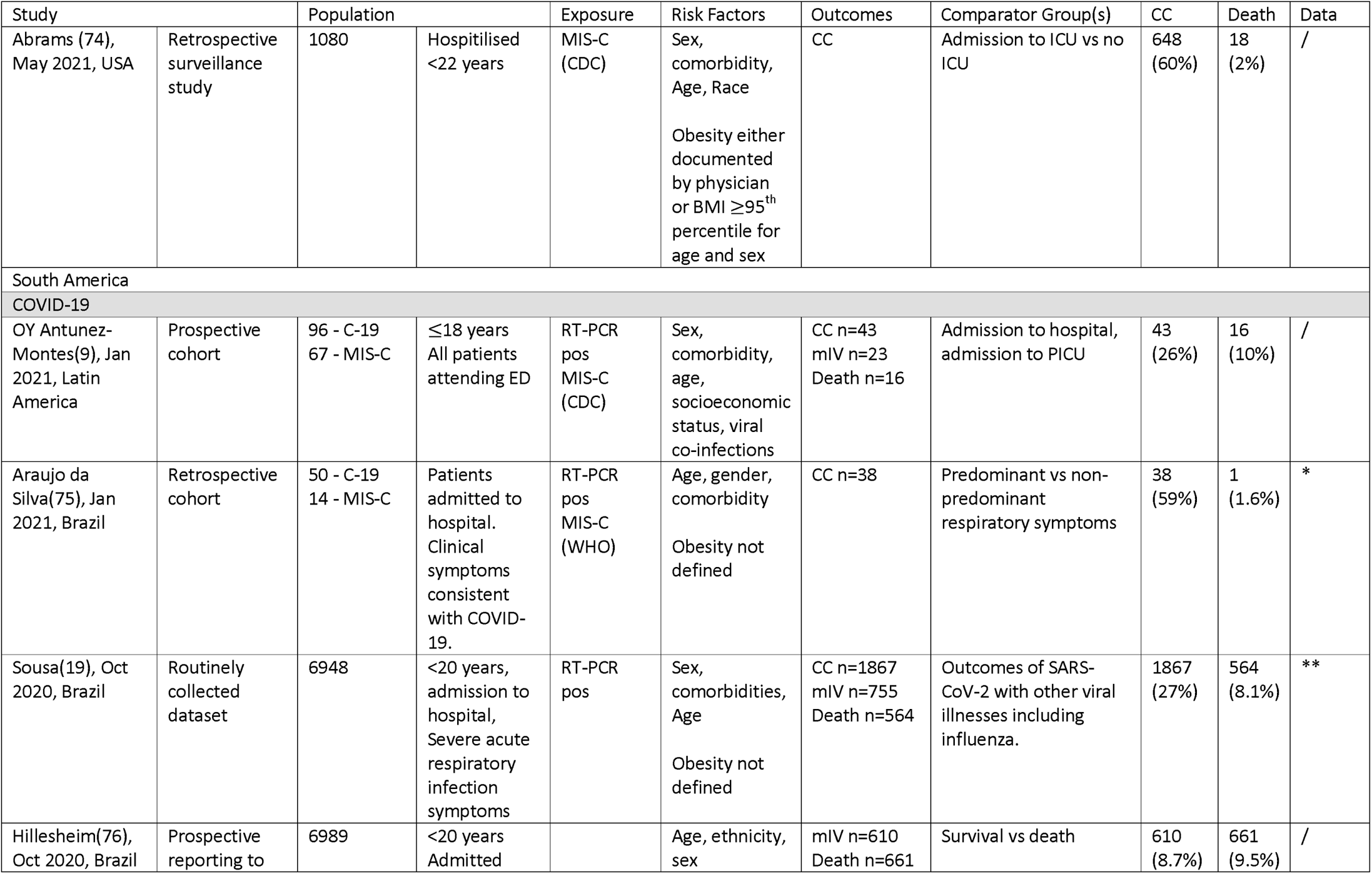
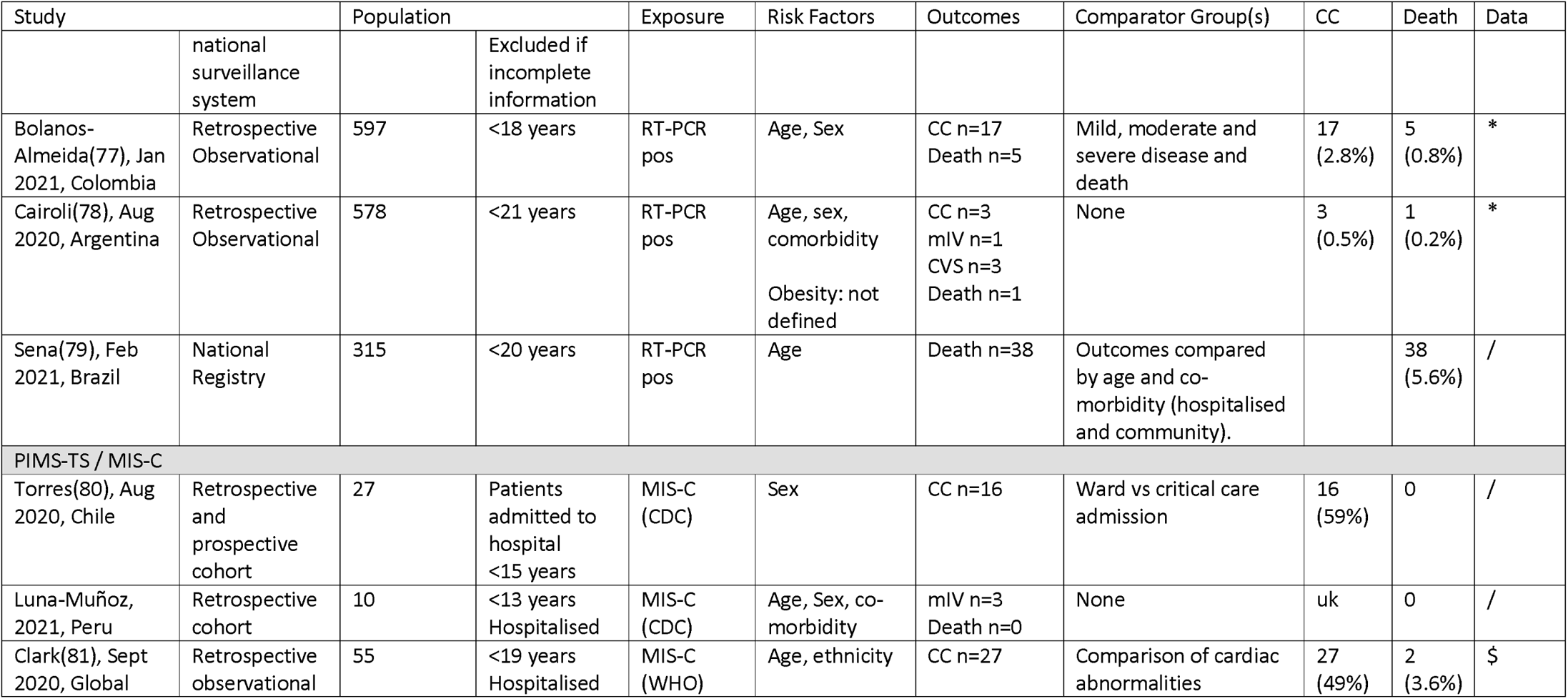
Study characteristics of ‘All comer’ studies for CYP with COVID-19 and PIMS-TS or MIS-C included in meta-analyses, grouped by region of origin

Data from 22 studies (40% of those in the meta-analysis) was included in the IPD meta-analyses, totalling 10,022 children. 26 studies reporting individual co-morbidities were eligible for inclusion in the narrative synthesis. Most studies eligible for inclusion in the meta-analysis were at considerable risk of bias (Figure 2).

We discuss findings from the aggregate and IPD meta-analyses for each set of risk factors and clinical outcomes below. Detailed data from included studies and pooled estimates from the aggregate meta-analyses are provided in Supplementary Table 1. Supplementary figures 1 and 2 show the sensitivity analysis with the largest study excluded. A two-stage meta-analysis using study-level estimates calculated from the IPD data is shown in supplementary figures 3 and 4.

Proportions of hospitalised children with COVID-19 admitted to critical care and who died in the aggregate analysis were 21.8% and 5.9% respectively and for PIMS-TS/MIS-C were 60.4% and 5.2%. In the IPD analysis, the proportion admitted to critical care with COVID-19 was 16.5% (6.7, 26.3) with death reported in 2.1% (−0.1, 4.3). For PIMS-TS/MIS-C, 72.6% (54.4, 90.7) were admitted to critical care and 7.41% (4.0, 10.8) died.

### Demographic risk factors for admission to critical care and death

Sex was not associated with pooled risk of admission to critical care or death in either COVID-19 or PIMS-TS in either the aggregate or IPD analyses (Figure 3). Compared with 1-4 year olds, the aggregate analysis found a higher pooled risk of critical care admission amongst 10-14 year olds and a higher risk of death amongst infants for COVID-19. In contrast, the IPD analysis found higher risk of critical care and death amongst both infants and 10-14 year olds, plus a higher odds of death amongst those >14 years for COVID-19. For PIMS-TS/MIS-C, the aggregate analysis found higher odds of critical care admission in all age-groups over 5 years, but no age-effects on risk of death. Numbers in the IPD analysis for PIMS-TS/MIS-C were very small, with no association of age-group with risk of death or critical care admission.

We were unable to assess the impact of ethnicity and socioeconomic position on clinical outcomes. The reporting of ethnicity data was highly variable and groupings were insufficiently similar across studies to allow meta-analysis. Socioeconomic position was reported by very few studies.

### Association of co-morbidities and critical care and death in aggregate meta-analysis

The aggregate meta-analysis compared those with any or specific comorbidities with all other children in each study (Figure 4). The presence of any comorbidity increased odds of critical care and death in COVID-19, with pooled odds ratios of 2.56 (1.77, 3.71) for critical care and 4.16 (1.97, 8.80) for death, both with moderate to high heterogeneity. Pooled odds ratios for PIMS-TS/MIS-C were of a similar order but with wide confidence intervals.

Pooled odds of both critical care admission and death in COVID-19 were increased in children with the following co-morbidities: cardiovascular; gastrointestinal or hepatic; neurological; chronic kidney disease; endocrine conditions, including diabetes; and metabolic conditions, including obesity. Odds ratios for critical care ranged from 2.5 to 3.1 and for death from 2.9 to 13. The presence of asthma or trisomy 21 was not associated with either outcome, while respiratory conditions were associated with increased odds of critical care but not death. There was an increased odds of death but not of critical care admission in those with malignancy, haematological conditions and immunosuppression for non-malignant reasons.

Few individual comorbidities were associated with odds of critical care or death in PIMS-TS / MIS-C, with the exception of malignancy (OR for death 183 (2.61-12,815) and metabolic diseases including obesity (OR for critical care 1.45 (1.10-1.92)).

### Association between co-morbidities and critical care and death in IPD meta-analysis

The IPD analysis compared those with each co-morbidity with children without any co-morbidity and additionally enabled analysis of risk associated with multiple comorbidities, obesity without other comorbidity, and trisomy 21 without cardiovascular disease. Figure 5 shows pooled OR for critical care and death for each comorbidity, and Figure 6 shows the risk difference estimated from the same models compared with children without comorbidities.

In IPD analysis, the presence of any comorbidity increased odds of critical care and death in COVID-19. The pooled odds ratio for admission to critical care was 1.64 (1.59-1.69), with risk difference being 4.6% (2.5, 6.7) greater than the 16.2% prevalence of critical care admission in those without comorbidities. The pooled odds of death from COVID-19 in those with any comorbidity was 2.49 (2.34-2.66), with a risk difference of 2.1% (−0.03, 4.2) above the 1.69% risk in those without comorbidity. For PIMS-TS/MIS-C, pooled odds of critical care was 12.44 (9.74-15.87) and risk difference 21.1% (4.4, 37.8) above baseline risk of 74.5%, and pooled odds of death was 11.23 (0.77, 163.22) with risk difference 21.0% (−3.4, 45.3) above baseline risk of death of 3.1%.

Number of comorbidities increased the odds of critical care and death in COVID-19 in an apparently dose-dependent fashion, with those with ≥3 comorbidities having a odds ratio of death of 4.98 (3.78, 6.56), twice that of the odds with one comorbidity. Small numbers with PIMS-TS / MIS-C meant that further analysis of co-morbidities could not be undertaken.

All individual comorbidities increased odds of admission to critical care except for malignancy and asthma, the latter associated with reduced odds (−0.6% (−1.0, −0.2). Risk differences for critical care above the risk for the no comorbidities group were highest for cardiovascular, neurological, and gastrointestinal conditions, as well as for obesity. Obesity alone, without other conditions, increased risk difference to the same level as cardiovascular or neurological conditions, although numbers were small in the obesity analyses.

Odds of death in COVID-19 in the IPD analyses was elevated in all comorbidity groups except for asthma, where there was a reduced risk (−0.6% (−0.9, −0.3)). Risk difference additional to the no comorbidity group was highest for malignancy. Trisomy 21 increased risk of death in those with or without comorbid cardiovascular disease.

### Narrative findings from studies of specific comorbidities

Twenty-six papers met the inclusion criteria for the narrative synthesis (Table 2), all reporting on the association of co-morbidity with acute COVID-19. Malignancy was the focus of sixteen of the studies, with rates of critical care admission in hospitalised patients ranging from 0-45% and of death in 0-47%. Six of the ten studies reporting deaths in this group of patients noted that some or all of the reported deaths were due to the underlying condition rather than SARS-CoV-2 infection.

**Table 2.**
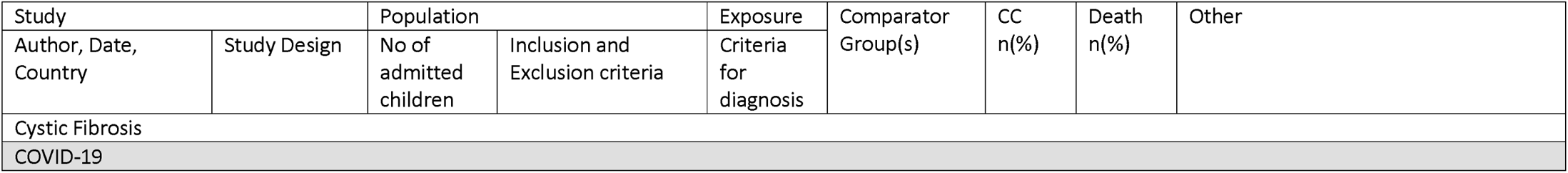
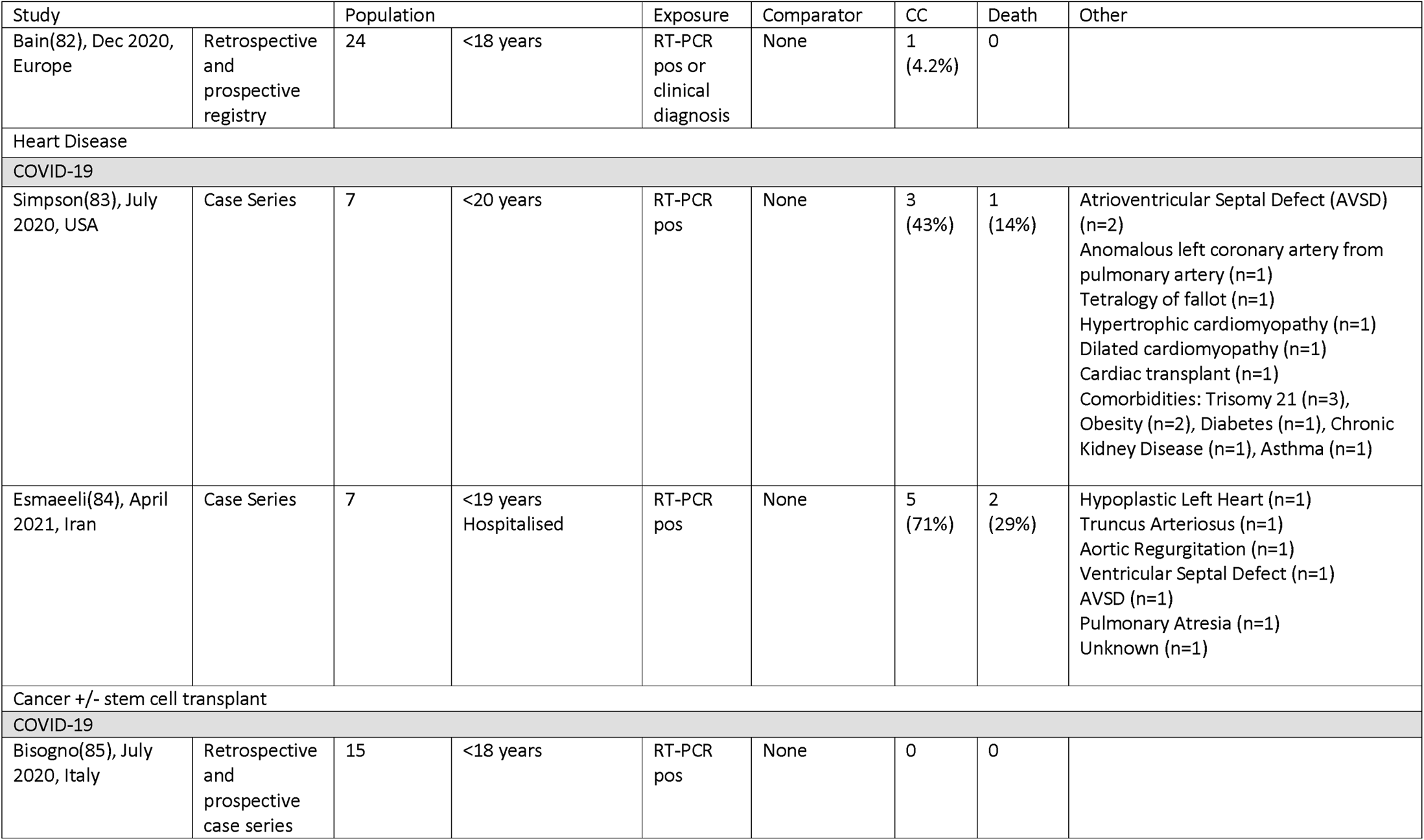
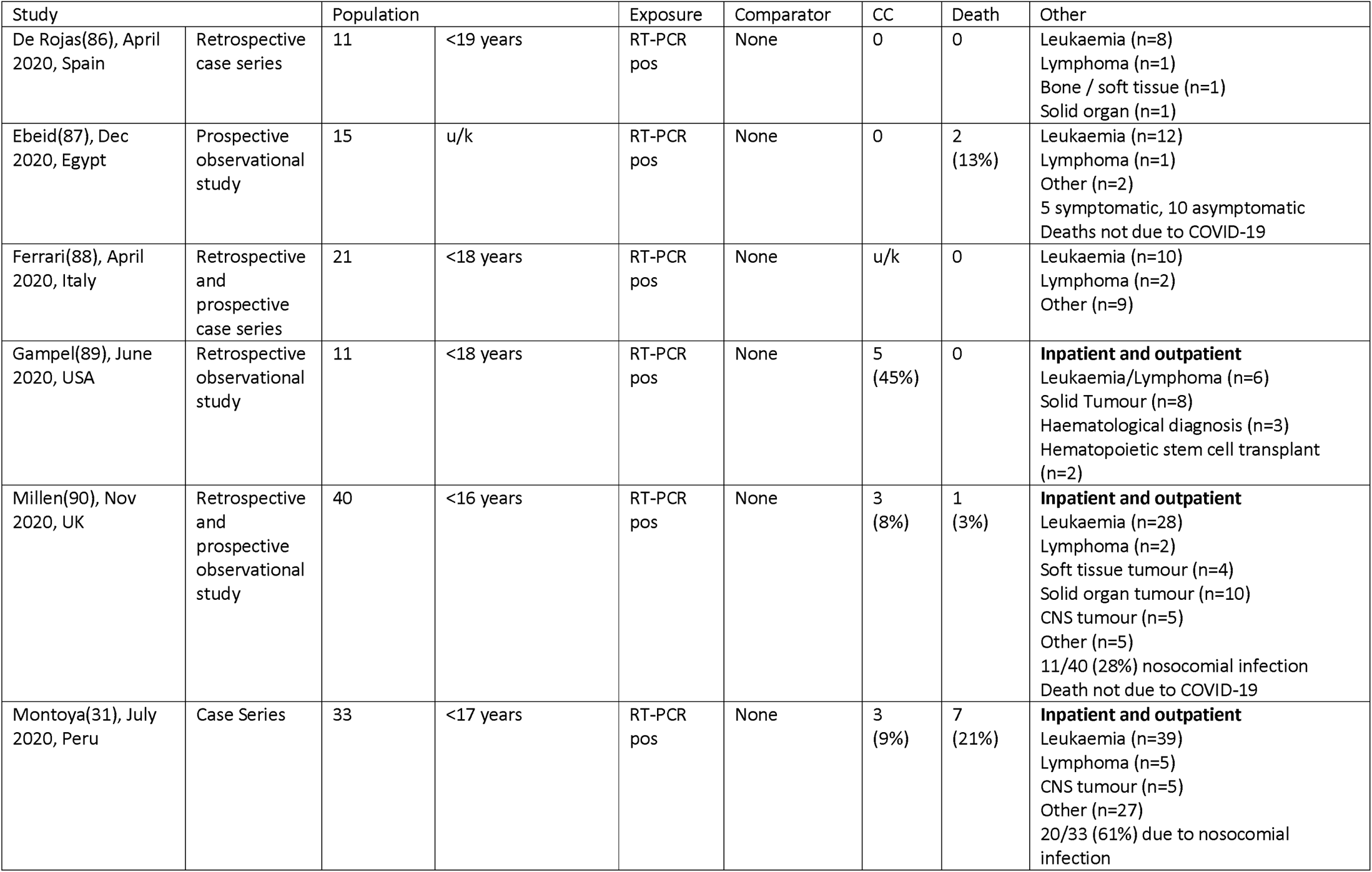
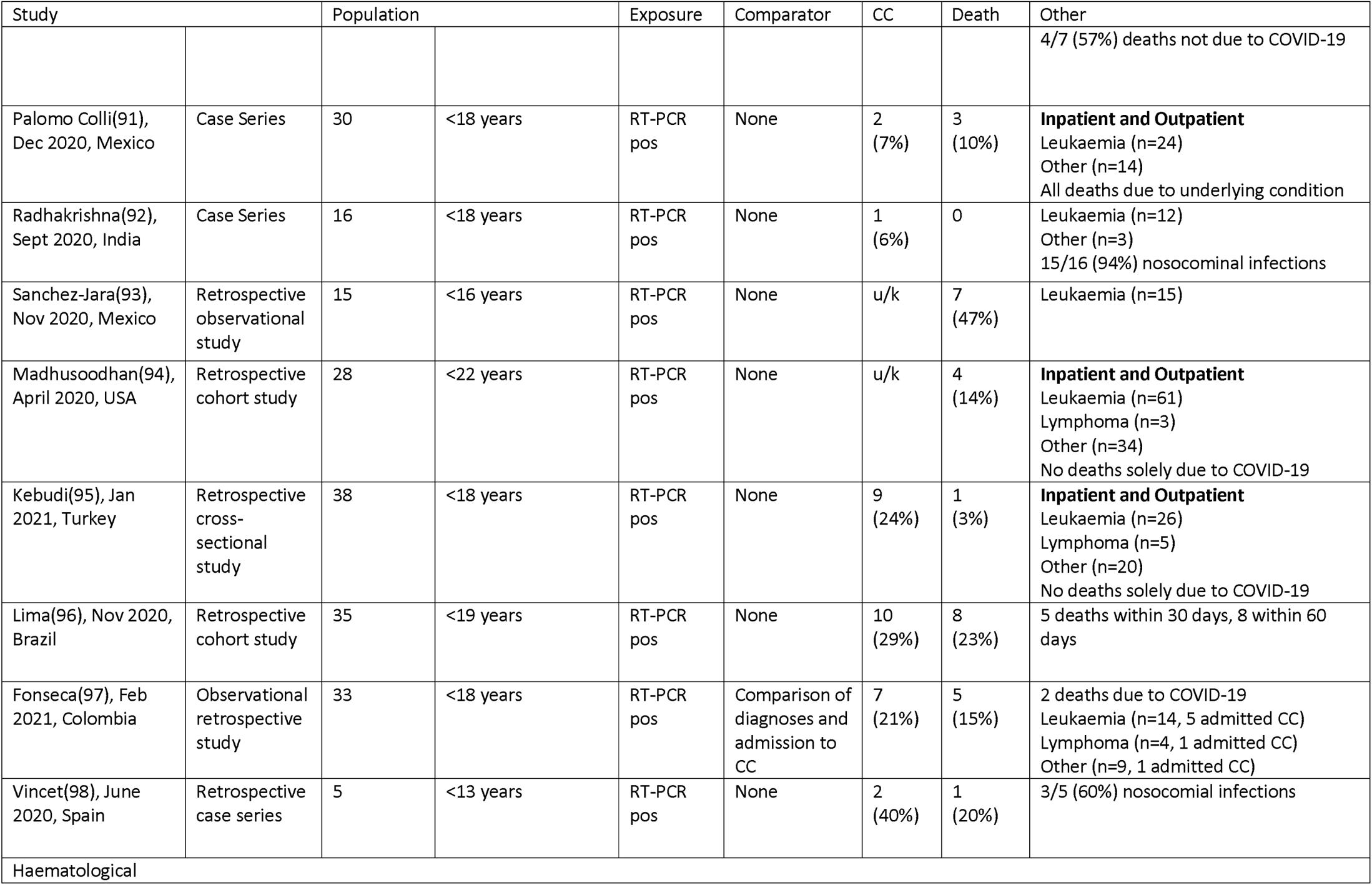
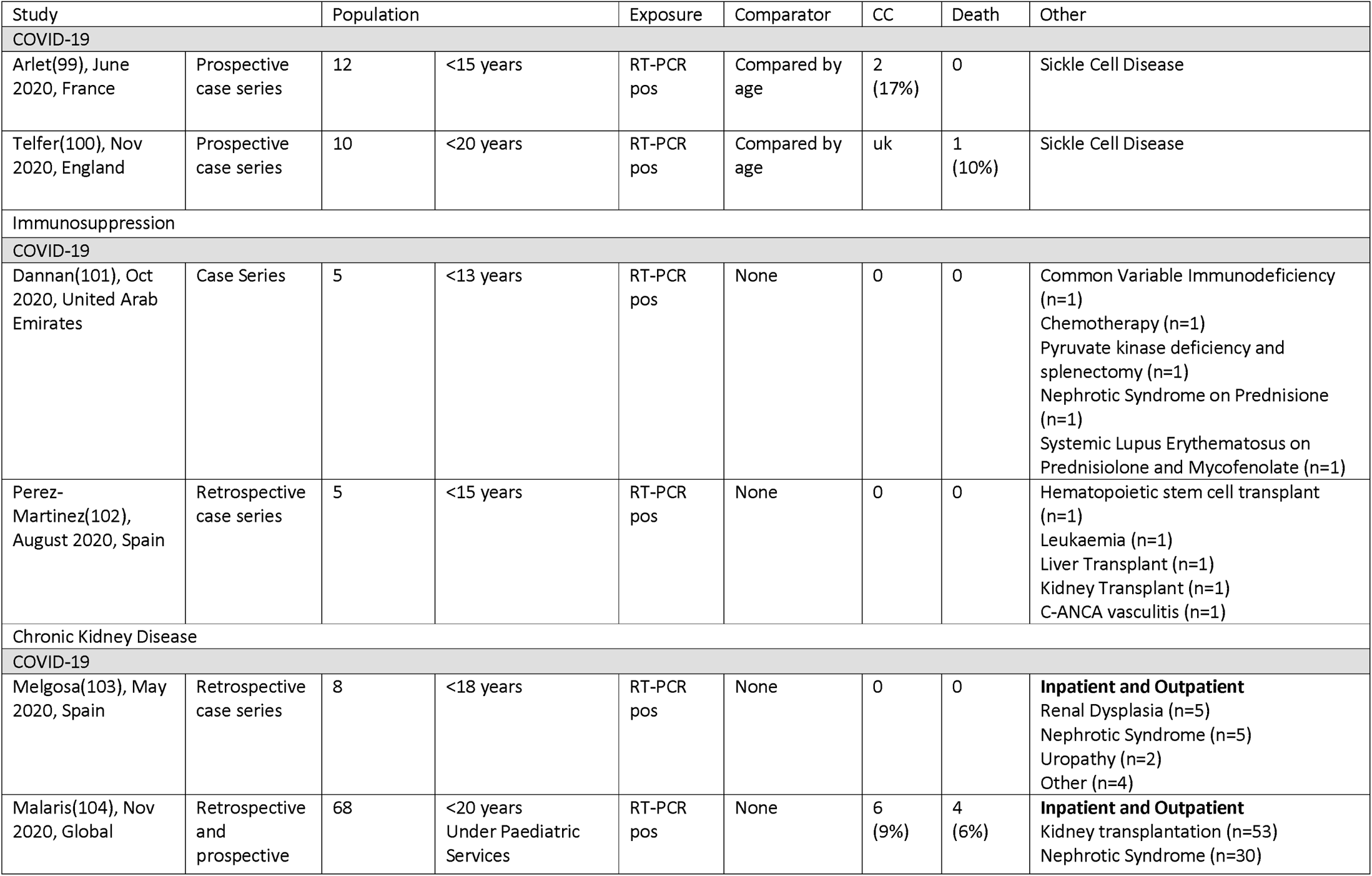
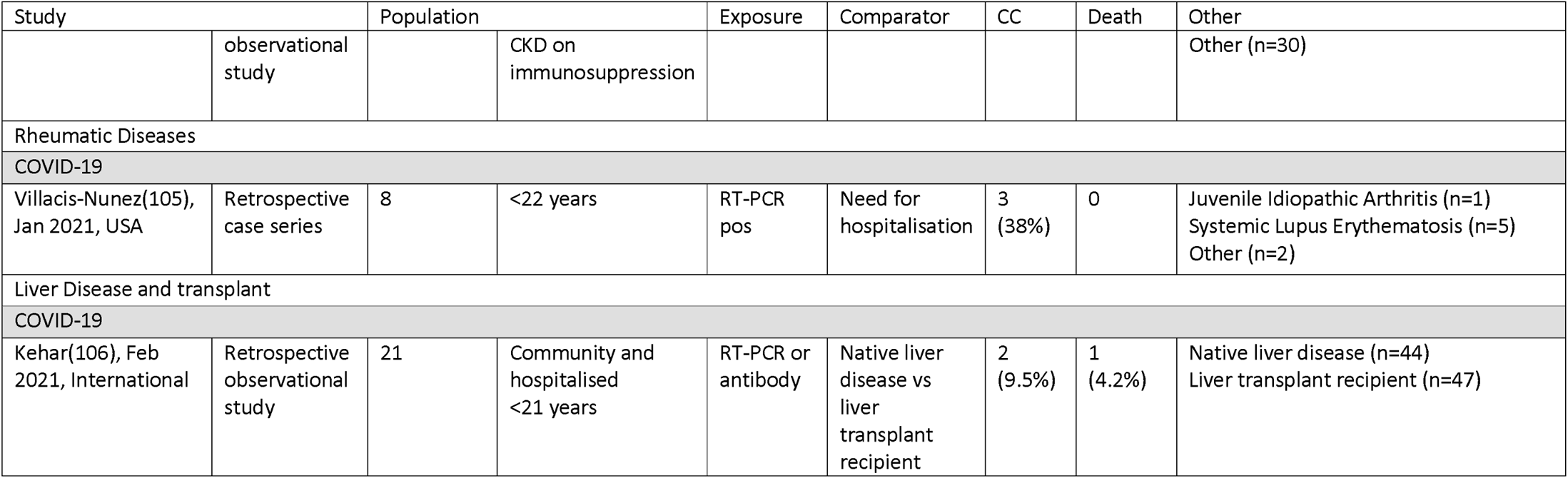
Study characteristics of comorbidity studies for CYP with COVID-19 and PIMS-TS or MIS-C. Admission to critical care - CC.

Two studies focussed on hospitalised patients with sickle cell disease. There were fewer than fifteen patients in each study, with 17% of patients being admitted to critical care in one study and reported deaths in 0-10%. Two studies looking at non-malignant immunosuppression described no children requiring critical care admission or death and a study of children with Rheumatic diseases found a rate of critical care admission of 38%. Chronic kidney disease was examined in two studies with small numbers of hospitalised patients, which describe a rate of critical care admission between 0-9% and of death between 0-6%. A study of CYP with cystic fibrosis found that 1 in 24 (4%) of those hospitalised were admitted to critical care and no deaths were described. Finally, two studies describe the association between pre-existing cardiac co-morbidity and outcome, which show a high proportion of children are admitted to critical care (43-71%) and that 14-29% are reported to die.

## Discussion

We present the first individual patient meta-analysis of risk factors for severe disease and death in CYP from both COVID-19 and PIMS-TS/MIS-C, nested within a broad systematic review and meta-analysis of published studies from the first pandemic year. Studies were of mixed quality and most were open to substantial bias; yet our meta-analyses included data from 57 studies from 19 countries, including 8 low or middle-income countries (LMIC).

Across both the aggregate and IPD analyses, we found no association between sex and odds of severe disease or death for either COVID-19 or PIMS-TS/MIS-C. We found that odds of poor outcomes was 1.6 to 2-fold higher for infants than 1-4 year olds for COVID-19 alone, but that teenagers had elevated odds of severe COVID-19 (1.4 to 2.2-fold higher odds) and particularly PIMS-TS/MIS-C (2.5 to 8-fold greater odds).

The presence of underlying comorbid conditions were the strongest risk factors for critical care admission and death. The presence of any comorbidity increased odds of poor outcomes in COVID-19 for both the aggregate and IPD analyses, increasing absolute risk of critical care admission by 4.5% (a relative increase of 28%) and risk of death by 2.5% (125% relative increase), with an even greater 21% increase in risk of death for PIMS-TS/MIS-C (6.8-fold increase in risk). Whilst one comorbidity increased absolute risk of critical care by 3.6% and death by 1.5% in COVID-19, 2 or more comorbidities dramatically increased the absolute risk increase.

All comorbidities were associated with increased risk across the two analyses, with the exception of asthma. Increase in odds of poor outcomes in COVID-19 was highest amongst those with cardiovascular, respiratory, neurological, and gastrointestinal comorbidities, each increasing absolute risk of critical care by 8-11% and risk of death by 1-3.5%. Malignancy was associated with increased of death from COVID-19, but not critical care admission in both analyses, which is counter-intuitive and raises the possibility that this reflects the high mortality rate amongst cancer survivors who may have incidentally died with SARS-CoV-2 positivity. The aggregated analysis did not suggest increased risk in those with immunosuppression (outside malignancy) or with haematological conditions when compared to children without those comorbidities, but these groups were at increased risk in the IPD analysis.

The risk factors we identified for more severe COVID-19, older age and the presence of comorbidities, particularly cardiovascular, respiratory, neurological, and gastrointestinal comorbidities, and obesity, are highly similar to those risk factors now well described for adults.(20) This suggests that risk factors for severe COVID-19 are consistent across the life-course, but previously not well understood in CYP because of the rarity of severe disease. Our findings relate to risk factors for severe disease rather than risk factors for infection, as we included only hospitalised children. It is likely that our findings may over-estimate risks of critical care and death for CYP in high income countries, as the mortality rate in our analyses (2.1% of children with COVID and 7.41% of those with PIMS-TS/MIS-C) are very much higher than national mortality rates reported from these settings (21–23). This likely reflects inclusion of studies from LMIC and publication bias towards more severe cases. Despite this the additional absolute risks related to all comorbidities was small compared with those without comorbidities.

### Comparison with the literature

Our finding of no differences in risk by sex is contrary to a large literature showing that males are more vulnerable to severe illness and death in childhood(24, 25). Whilst male sex is a known risk factor for more severe COVID-19 in adults, this excess risk arises only after middle age (26). We found that obesity, whether alone or with other conditions, markedly increased risk of critical care admission and death in the IPD analysis. Whilst numbers with obesity were very small, our findings are consistent with adult data showing obesity to be one of the strongest risk factors for severe disease in adults(27).

Previous reviews have not provided a systematic understanding of the associations of paediatric comorbidities and severe outcomes in children. Systematic reviews which were undertaken early in the pandemic highlighted some of the challenges around identifying comorbidities which were associated with severe disease, including pooled reporting of even common conditions such as asthma(28) and a focus on individual comorbidities without a comparator group(29). Our finding that children with trisomy 21 were at increased risk of critical care admission and death has not been described before, although it is consistent with previous adult data(30). We found that this risk appears to operate both through and independently of cardiovascular anomalies, indicating that all CYP with trisomy 21 are at some increased risk of severe disease.

### Strengths and limitations

We undertook a high quality systematic review and meta-analysis, including use of individual patient data to provide more detailed analysis of comorbidities, including obesity and trisomy 21. Findings were largely consistent across the aggregate and the IPD analyses, with some exceptions. Findings from a sensitivity analysis excluding the largest study and from a two-stage meta-analysis of the IPD were highly similar to those described above.

Our data are subject to a number of limitations. Twenty-two of 57 studies (39%) provided individual patient data; systematic differences between these groups may have introduced bias. There were very small numbers with PIMS-TS/MIS-C in some analyses, particularly the IPD analyses. We were unable to examine ethnicity and socioeconomic position as risk factors due to lack of data in included studies.

Included studies were highly heterogenous and from a wide range of resource settings, and it is likely that findings were influenced by differing national approaches to hospitalisation of infected children and by differences in availability and use of resources including intensive care beds. A number of East Asian countries hospitalised all children who were SARS-CoV-2 positive, regardless of symptoms, whilst other countries limited hospitalisation to symptomatic children or those with significant illness. Policies on admission to and access to critical care likely also differed between countries(31). The novel nature of PIMS-TS/MIS-C also likely influenced critical care admission thresholds for this condition. Definitions of comorbidities were also heterogenous across studies and some of our comorbidity groups may be subject to misclassification bias. The definition of obesity in most studies related to severe or extreme obesity rather than the more common condition of being overweight, yet obesity was undefined in a number of studies.

We were unable to separate the increased risk for severe disease related to comorbidities from the underlying risks of illness and death seen in these comorbidities in uninfected children, as all children in our study had SARS-CoV-2. Yet studies of infected hospitalised cohorts are essential to understand how rare congenital or acquired comorbidities may influence risk of severe disease or death from SARS-CoV-2

Whilst our review examined comorbidities as risk factors in more detail than previous studies, there were limited data on sub-types of comorbidities, e.g. whether neurological problems were epilepsy or more complex neurodisability, and on combinations of comorbidities. Our finding that cardiovascular, neurological, and gastrointestinal conditions were associated with the highest risk of poor outcome, a risk similar to having 2 or more comorbidities, may reflect that these conditions were more likely to be comorbid with others. Given the low risk to children requiring hospital admission or critical care as a direct consequence of SARS-CoV-2 infection, it is likely that a significant number of reported cases were coincidental cases of SARS-CoV-2 seropositivity reflecting population prevalence.

### Conclusions and implications

When children are admitted to hospital with SARS-CoV-2 infection, those at greatest risk of severe disease or death are teenagers, those with cardiac or neurological conditions, or 2 or more comorbid conditions, and children who are significantly obese. These groups should be considered higher priority for vaccination and for protective shielding when appropriate. Our findings suggest that established risk factors for severe disease in adults also operate in the early life-course. Whilst odd ratios for poor outcomes were increased for nearly all comorbidities in children, the absolute increase in risk for most comorbidities was small compared to children without underlying conditions. This emphasises that our findings should be understood within the broader context that risk of severe disease and death from COVID-19 and PIMS-TS/MIS-C in hospitalised children is very low compared with adults.

Our study quantifies the additional risk related to comorbidities in infected children, however it is likely that some or all of this risk relates to the underlying condition rather than SARS-CoV-2 infection. Further population-based research using comparator groups which identify the risk of severe disease due to COVID-19 and the underlying risk due to comorbidity is required to develop a safe approach to shielding and vaccination for children.

## Supporting information

PRISMA

## Data Availability

All data are from published studies and are summarised in our manuscript

## Acknowledgements

We thank the authors who shared patient level data to enable their inclusion in this study (Supplementary Information 1) and the Royal College of Paediatrics and Child Health who assisted with data extraction.

## Contributions

Study Design: RH, NT, CS, JW, C T-S, ML, MC, EW, PJD, KL, ESD, SK, LF and RMV, Literature search, identification of papers and data extraction: RH, HY, NT, CS, JW, SK and LF, Data analysis: RH, CT-S and RV, First Draft: RH, Review and editing: All authors

