## Supplement for "Which children and young people are at higher risk of severe disease and death after SARS-CoV-2 infection: a systematic review and individual patient meta-analysis"

**Supplementary Material**

**Full Search Terms**

Search Terms used for Pubmed:

PubMed: ((COVID-19[Supplementary Concept] OR severe acute respiratory syndrome coronavirus 2[Supplementary Concept] OR Coronavirus Infections[MeSH Terms] OR  Wuhan coronavirus[Text Word] OR Wuhan seafood market pneumonia virus[Text Word] OR COVID19*[Text Word] OR COVID-19*[Text Word] OR COVID-2019*[Text Word] OR coronavirus disease 2019[Text Word] OR SARS-CoV-2[Text Word] OR SARS2[Text Word] OR 2019-nCoV*[Text Word] OR 2019 novel coronavirus[Text Word] OR severe acute respiratory syndrome coronavirus 2[Text Word] OR 2019 novel coronavirus infection[Text Word] OR coronavirus disease 2019[Text Word] OR coronavirus disease-19[Text Word] OR novel coronavirus[Text Word] OR coronavirus[Text Word] OR SARS-CoV-19[Text Word] OR PIMS-TS[Text Word] OR paediatric inflammatory multisystem syndrome [Text Word] OR MIS-C[Text Word] OR multi-system inflammatory syndrome in children[Text Word]) AND (minors [Text Word]OR minors* [Text Word]OR boy [Text Word]OR boys[Text Word] OR girl*[Text Word] OR kid[Text Word] OR kids[Text Word] OR child[Text Word] OR child*[Text Word] OR children*[Text Word] OR schoolchild*[Text Word] OR schoolchild[Text Word] OR school child [Text Word] OR school child*[Text Word] OR adolescen* [Text Word]OR juvenil* [Text Word]OR youth* [Text Word]OR teen* [Text Word]OR under*age* [Text Word]OR pubescen* [Text Word] OR pediatrics[mh] OR pediatric* [Text Word]OR paediatric* [Text Word]OR pediatric* [Text Word]))

Filters:
Language: English only
Article type: Clinical study, clinical trial, comparative study, journal article, multicentre study, observational study, pragmatic clinical trial, randomized control trial
Age: Child: birth-18 years, Newborn: birth-1 month, Infant: birth-23 months, Infant: 1-23 months, preschool child: 2-5 years, child: 6-12 years, adolescent: 13-18 years

### Sensitivity analysis - association between demographics and outcome

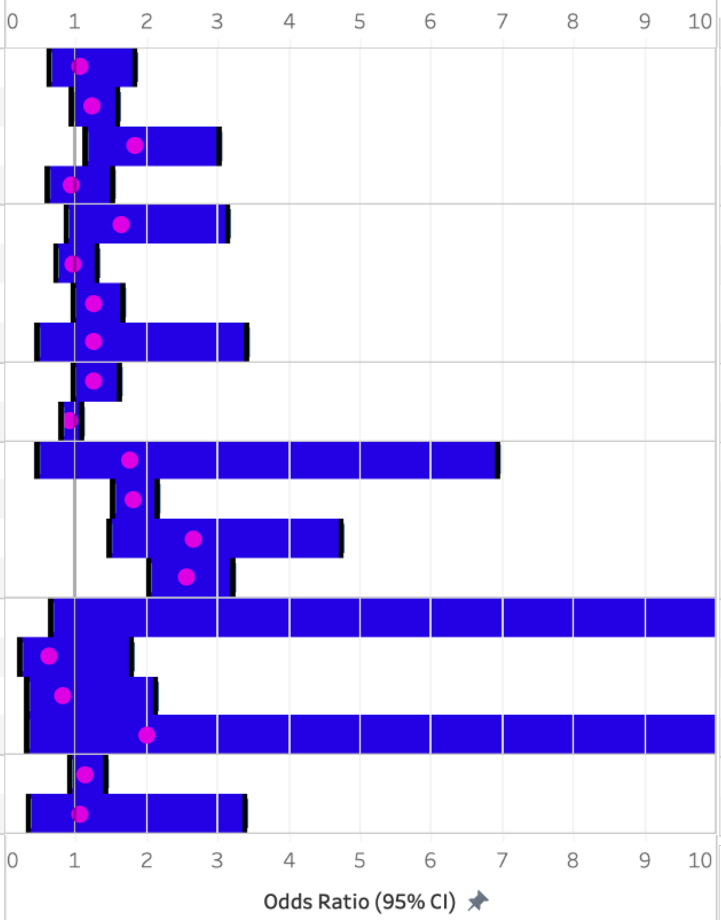


| | | | LCI | OR | UCI | I2 | p |
|---|---|---|---|---|---|---|---|
| COVID-19 | Critical Care Admission | <1 year | 0.62 | 1.07 | 1.83 | 58.46 | 0.81 |
| | | 5-9 years | 0.94 | 1.22 | 1.59 | 0.00 | 0.14 |
| | | 10-14 years | 1.13 | 1.84 | 3.02 | 25.47 | 0.02 |
| | | >14 years | 0.60 | 0.95 | 1.51 | 58.32 | 0.83 |
| | Death | <1 year | 0.87 | 1.65 | 3.14 | 27.49 | 0.13 |
| | | 5-9 years | 0.73 | 0.97 | 1.30 | 0.00 | 0.85 |
| | | 10-14 years | 0.96 | 1.27 | 1.67 | 0.00 | 0.09 |
| | | >14 years | 0.46 | 1.25 | 3.40 | 78.56 | 0.66 |
| | Sex | Admission to critical care | 0.97 | 1.25 | 1.61 | 32.51 | 0.09 |
| | | Death | 0.79 | 0.92 | 1.08 | 0.00 | 0.32 |
| PIMS-TS / MIS-C | Critical Care Admission | <1 year | 0.45 | 1.77 | 6.94 | 0.00 | 0.41 |
| | | 5-9 years | 1.51 | 1.81 | 2.16 | 0.00 | 0.00 |
| | | 10-14 years | 1.48 | 2.65 | 4.74 | 21.32 | 0.00 |
| | | >14 years | 2.03 | 2.56 | 3.21 | 0.00 | 0.00 |
| | Death | <1 year | 0.64 | 14.50 | 328.46 | 0.00 | 0.09 |
| | | 5-9 years | 0.22 | 0.63 | 1.78 | 0.00 | 0.38 |
| | | 10-14 years | 0.31 | 0.82 | 2.13 | 0.00 | 0.68 |
| | | >14 years | 0.31 | 2.01 | 12.87 | 38.13 | 0.46 |
| | Sex | Admission to critical care | 0.92 | 1.15 | 1.43 | 0.00 | 0.23 |
| | | Death | 0.34 | 1.07 | 3.38 | 0.00 | 0.09 |

Supplementary Figure 1. Sensitivity analysis of aggregated demographic data used in meta-analysis. Largest study included in the meta-analysis excluded.

Sensitivity analysis - association between co-morbidities and outcome

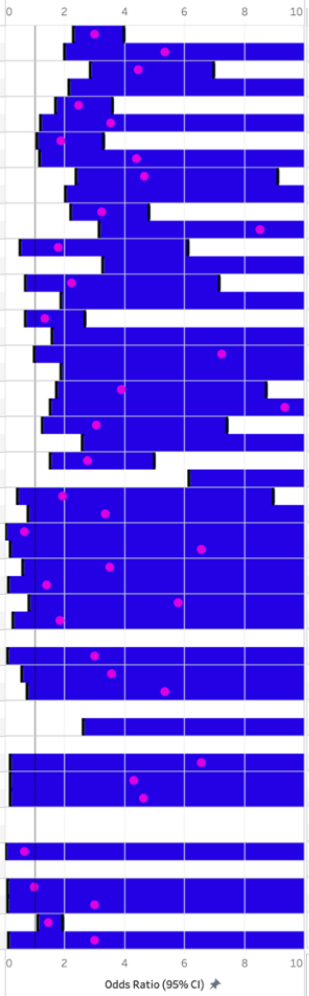


| | | | LCI | OR | UCI | I2 | p |
|---|---|---|---|---|---|---|---|
| COVID-19 | Any Co-morbidity | Admission to critical care | 2.27 | 3.01 | 3.97 | 19.41 | 0.00 |
| | | Death | 1.99 | 5.33 | 14.26 | 10.20 | 0.00 |
| | Cardiovascular | Admission to critical care | 2.84 | 4.44 | 6.96 | 0.00 | 0.00 |
| | | Death | 2.15 | 16.62 | 128.70 | 66.14 | 0.01 |
| | Respiratory | Admission to critical care | 1.68 | 2.46 | 3.59 | 0.00 | 0.00 |
| | | Death | 1.18 | 3.53 | 10.55 | 0.00 | 0.02 |
| | Asthma | Admission to critical care | 1.08 | 1.88 | 3.29 | 0.00 | 0.03 |
| | | Death | 1.16 | 4.38 | 16.50 | 0.00 | 0.03 |
| | Gastrointestinal / Liver | Admission to critical care | 2.37 | 4.65 | 9.12 | 0.00 | 0.00 |
| | | Death | 2.01 | 13.06 | 84.84 | 0.00 | 0.01 |
| | Neurological | Admission to critical care | 2.18 | 3.24 | 4.81 | 0.00 | 0.00 |
| | | Death | 3.14 | 8.51 | 23.08 | 0.00 | 0.00 |
| | Malignancy | Admission to critical care | 0.51 | 1.77 | 6.11 | 58.59 | 0.36 |
| | | Death | 3.27 | 10.76 | 35.41 | 30.99 | 0.00 |
| | Haematological | Admission to critical care | 0.70 | 2.23 | 7.14 | 0.00 | 0.18 |
| | | Death | 1.88 | 11.74 | 73.56 | 0.00 | 0.01 |
| | Immunosuppression | Admission to critical care | 0.68 | 1.35 | 2.68 | 26.87 | 0.40 |
| | | Death | 1.57 | 11.95 | 90.74 | 16.07 | 0.02 |
| | T21 | Admission to critical care | 0.99 | 7.25 | 53.10 | 31.57 | 0.05 |
| | | Death | 1.87 | 17.07 | 156.14 | 0.00 | 0.01 |
| | CKD | Admission to critical care | 1.73 | 3.88 | 8.72 | 0.00 | 0.00 |
| | | Death | 1.52 | 9.35 | 57.39 | 0.00 | 0.02 |
| | Endocrine including diabetes | Admission to critical care | 1.26 | 3.06 | 7.41 | 17.65 | 0.01 |
| | | Death | 2.57 | 13.17 | 67.45 | 0.00 | 0.00 |
| | Metabolic inc obesity | Admission to critical care | 1.51 | 2.75 | 4.98 | 17.25 | 0.00 |
| | | Death | 6.14 | 28.54 | 132.61 | 0.00 | 0.00 |
| PIMS-TS / MIS-C | Any Co-morbidity | Admission to critical care | 0.41 | 1.93 | 8.97 | 59.89 | 0.40 |
| | | Death | 0.79 | 3.36 | 14.40 | 0.00 | 0.10 |
| | Cardiovascular | Admission to critical care | 0.02 | 0.65 | 21.18 | 0.00 | 0.81 |
| | | Death | 0.18 | 6.56 | 239.09 | 0.00 | 0.31 |
| | Respiratory | Admission to critical care | 0.59 | 3.50 | 21.00 | 16.37 | 0.17 |
| | | Death | 0.12 | 1.41 | 16.70 | 0.00 | 0.79 |
| | Asthma | Admission to critical care | 0.80 | 5.78 | 41.84 | 13.73 | 0.08 |
| | | Death | 0.26 | 1.84 | 12.77 | 0.00 | 0.54 |
| | Gastrointestinal / Liver | Admission to critical care | | | | | |
| | | Death | 0.08 | 3.00 | 111.78 | 0.00 | 0.55 |
| | Neurological | Admission to critical care | 0.58 | 3.57 | 22.08 | 0.00 | 0.17 |
| | | Death | 0.74 | 5.33 | 38.45 | 0.00 | 0.10 |
| | Malignancy | Admission to critical care | | | | | |
| | | Death | 2.61 | 183.00 | 12,814.00 | 0.00 | 0.02 |
| | Haematological | Admission to critical care | | | | | |
| | | Death | 0.18 | 6.56 | 239.09 | 0.00 | 0.31 |
| | Immunosuppression | Admission to critical care | 0.17 | 4.31 | 109.74 | 0.00 | 0.38 |
| | | Death | 0.16 | 4.63 | 130.71 | 0.00 | 0.37 |
| | T21 | Admission to critical care | | | | | |
| | | Death | | | | | |
| | CKD | Admission to critical care | 0.02 | 0.65 | 21.18 | 0.00 | 0.81 |
| | | Death | | | | | |
| | Endocrine including diabetes | Admission to critical care | 0.09 | 0.98 | 11.17 | 0.00 | 0.98 |
| | | Death | 0.08 | 3.00 | 111.78 | 0.00 | 0.55 |
| | Metabolic inc obesity | Admission to critical care | 1.10 | 1.45 | 1.92 | 0.00 | 0.01 |
| | | Death | 0.12 | 3.00 | 73.64 | 0.00 | 0.50 |

Supplementary Figure 2. Sensitivity analysis of aggregated co-morbidity data used in meta-analysis. Largest study included in the meta-analysis excluded.

Meta-analysis of studies providing IPD - Association between demographics and severe outcome

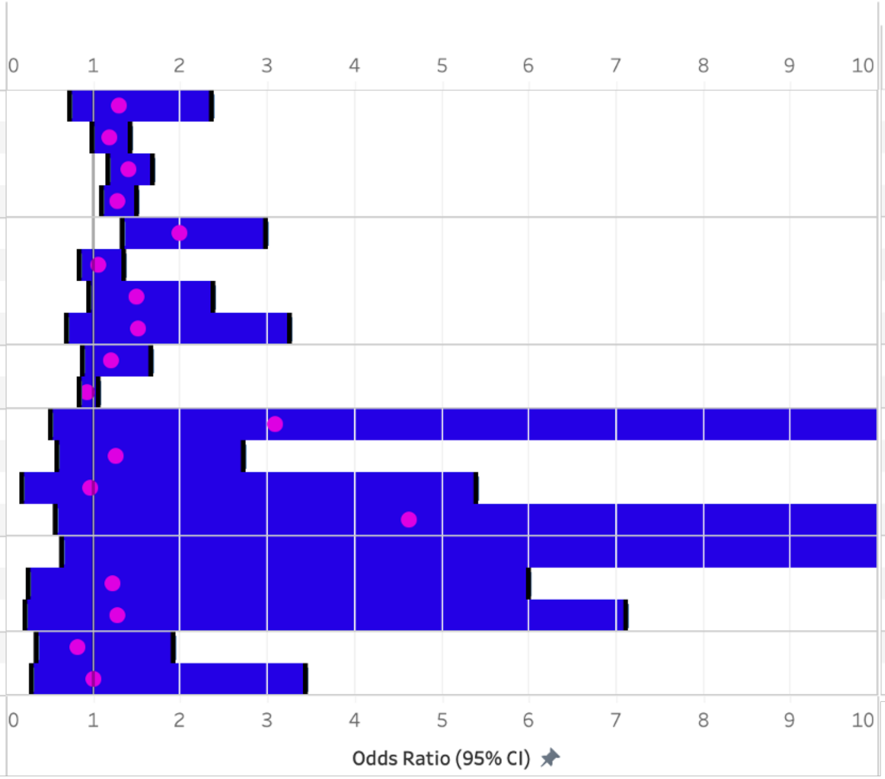


| Group | Sub cat | Outcome | LCI | OR | UCI | I2 | p |
|---|---|---|---|---|---|---|---|
| | | | Measure | | | | |
| COVID-19 | Critical Care Admission | <1 year | 0.72 | 1.31 | 2.36 | 44.65 | 0.38 |
| | | 5-9 years | 0.99 | 1.18 | 1.42 | 0.00 | 0.07 |
| | | 10-14 years | 1.17 | 1.40 | 1.68 | 0.00 | 0.00 |
| | | >14 years | 1.09 | 1.28 | 1.49 | 0.00 | 0.00 |
| | Death | <1 year | 1.34 | 1.99 | 2.98 | 39.71 | 0.00 |
| | | 5-9 years | 0.85 | 1.07 | 1.34 | 0.00 | 0.57 |
| | | 10-14 years | 0.95 | 1.50 | 2.38 | 41.20 | 0.08 |
| | | >14 years | 0.70 | 1.51 | 3.25 | 83.64 | 0.29 |
| | Sex | Admission to critical care | 0.88 | 1.21 | 1.67 | 40.32 | 0.23 |
| | | Death | 0.84 | 0.94 | 1.05 | 0.00 | 0.28 |
| PIMS-TS / MIS-C | Critical Care Admission | <1 year | 0.51 | 3.10 | 18.68 | 0.00 | 0.22 |
| | | 5-9 years | 0.58 | 1.26 | 2.72 | 0.00 | 0.55 |
| | | 10-14 years | 0.18 | 0.97 | 5.40 | 44.93 | 0.98 |
| | | >14 years | 0.57 | 4.63 | 37.46 | 0.00 | 0.15 |
| | Death | <1 year | 0.64 | 14.50 | 328.46 | 0.00 | 0.09 |
| | | 5-9 years | 0.25 | 1.22 | 5.99 | 0.00 | 0.80 |
| | | 10-14 years | 0.23 | 1.27 | 7.11 | 0.00 | 0.78 |
| | Sex | Admission to critical care | 0.35 | 0.82 | 1.92 | 0.00 | 0.65 |
| | | Death | 0.29 | 1.00 | 3.44 | 0.00 | 1.00 |

Supplementary Figure 3. Meta-analysis of demographics of individual patient data.

### Meta-analysis of studies providing IPD - Association between co-morbidities and severe outcome

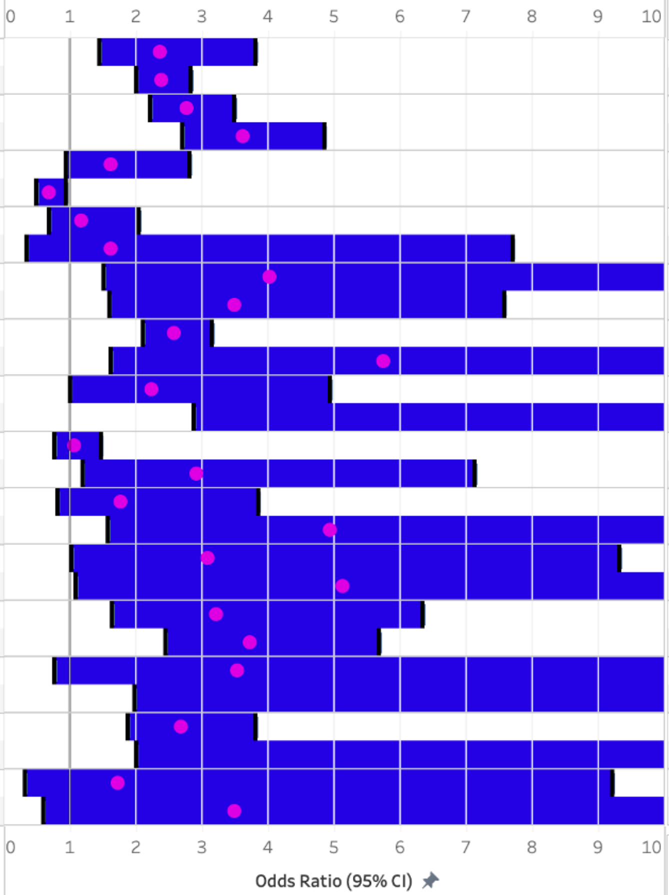


| Group | Sub cat | Outcome | LCI | OR | UCI | I2 | p |
|---|---|---|---|---|---|---|---|
| COVID-19 | Any Co-morbidity | Admission to critical care | 1.45 | 2.35 | 3.81 | 54.20 | 0.00 |
| | | Death | 2.01 | 2.39 | 2.84 | 0.00 | 0.00 |
| | Cardiovascular | Admission to critical care | 2.21 | 2.77 | 3.48 | 0.00 | 0.00 |
| | | Death | 2.70 | 3.62 | 4.85 | 0.00 | 0.00 |
| | Respiratory | Admission to critical care | 0.94 | 1.63 | 2.81 | 43.55 | 0.08 |
| | | Death | 0.49 | 0.68 | 0.95 | 0.00 | 0.02 |
| | Asthma | Admission to critical care | 0.68 | 1.18 | 2.05 | 23.30 | 0.56 |
| | | Death | 0.34 | 1.63 | 7.71 | 52.82 | 0.54 |
| | Gastrointestinal / Liver | Admission to critical care | 1.51 | 4.02 | 10.71 | 21.84 | 0.01 |
| | | Death | 1.60 | 3.48 | 7.58 | 0.00 | 0.00 |
| | Neurological | Admission to critical care | 2.10 | 2.57 | 3.14 | 0.00 | 0.00 |
| | | Death | 1.61 | 5.74 | 20.52 | 44.92 | 0.01 |
| | Malignancy | Admission to critical care | 1.01 | 2.23 | 4.93 | 0.00 | 0.05 |
| | | Death | 2.87 | 10.72 | 40.09 | 0.00 | 0.00 |
| | Haematological | Admission to critical care | 0.77 | 1.07 | 1.47 | 0.00 | 0.70 |
| | | Death | 1.20 | 2.92 | 7.13 | 13.84 | 0.02 |
| | Immunosuppression | Admission to critical care | 0.81 | 1.77 | 3.84 | 40.83 | 0.15 |
| | | Death | 1.57 | 4.93 | 15.50 | 26.34 | 0.01 |
| | T21 | Admission to critical care | 1.02 | 3.08 | 9.32 | 43.95 | 0.05 |
| | | Death | 1.08 | 5.12 | 24.40 | 40.64 | 0.04 |
| | CKD | Admission to critical care | 1.64 | 3.22 | 6.33 | 18.28 | 0.00 |
| | | Death | 2.44 | 3.72 | 5.67 | 0.00 | 0.00 |
| | Endocrine including diabetes | Admission to critical care | 0.77 | 3.54 | 16.35 | 20.48 | 0.11 |
| | | Death | 1.97 | 12.36 | 77.44 | 0.00 | 0.01 |
| | Metabolic inc obesity | Admission to critical care | 1.88 | 2.68 | 3.81 | 0.00 | 0.00 |
| | | Death | 2.01 | 11.86 | 69.96 | 57.48 | 0.01 |
| PIMS-TS / MIS-C | Any Co-morbidity | Admission to critical care | 0.33 | 1.73 | 9.22 | 38.24 | 0.52 |
| | | Death | 0.59 | 3.48 | 20.59 | 16.62 | 0.17 |

Supplementary Figure 4. Meta-analysis of co-morbidities of individual patient data.

Supplementary Table 1. Summary of all studies included in aggregated meta-analysis including outcomes of mechanical invasive ventilation and cardiovascular support

| Name | Events 1 | Total 1 | Events 2 | Total 2 | Effect Estimate | CI Start | CI End | Weight | P(Q) | I²(Q) | Z | P(Z) |
|---|---|---|---|---|---|---|---|---|---|---|---|---|
| Sex and Severe Outcome | | | | | | | | | | | | |
| Invasive Ventilation | 605 | 6384 | 500 | 6043 | 1.17 | 0.94 | 1.46 | 100.00 | 0.31 | 10.70 | 1.41 | 0.16 |
| COVID-19 | 556 | 6233 | 465 | 5948 | 1.24 | 0.92 | 1.66 | 86.15 | 0.14 | 30.00 | 1.43 | 0.15 |
| 12 - OY Antunez-Montes et al | 2 | 110 | 1 | 71 | 1.30 | 0.12 | 14.57 | 0.81 | | | | |
| 13 - AR Araujo da Silva et al | 3 | 34 | 1 | 15 | 1.35 | 0.13 | 14.20 | 0.86 | | | | |
| 142 - S Richardson et al | 8 | 51 | 6 | 59 | 1.64 | 0.53 | 5.10 | 3.49 | | | | |
| 151 - B Sousa et al | 404 | 3591 | 351 | 3341 | 1.08 | 0.93 | 1.26 | 38.87 | | | | |
| 155 - Kyung Sung | 0 | 54 | 0 | 47 | 0.00 | 0.00 | 0.00 | 0.00 | | | | |
| 15 - JP Armann et al | 3 | 45 | 3 | 57 | 1.29 | 0.25 | 6.70 | 1.71 | | | | |
| 174 - P Zachariah et al | 4 | 27 | 5 | 23 | 0.63 | 0.15 | 2.68 | 2.18 | | | | |
| 21 - D Bayesheva et al | 1 | 311 | 0 | 241 | 2.33 | 0.09 | 57.53 | 0.47 | | | | |
| 221 - Preston et al | 100 | 1083 | 72 | 1344 | 1.80 | 1.31 | 2.46 | 24.14 | | | | |
| 28 - H Cairoli | 1 | 315 | 0 | 263 | 2.51 | 0.10 | 61.96 | 0.47 | | | | |
| 46 - M Ceano-Vivas et al | 1 | 22 | 0 | 11 | 1.60 | 0.06 | 42.63 | 0.44 | | | | |
| 54 - A Desai et al | 10 | 156 | 18 | 137 | 0.45 | 0.20 | 1.02 | 6.37 | | | | |
| 75 - V Giacomet et al | 1 | 82 | 0 | 44 | 1.64 | 0.07 | 41.05 | 0.46 | | | | |
| 79 - F Gotzinger et al | 18 | 311 | 7 | 271 | 2.32 | 0.95 | 5.63 | 5.42 | | | | |
| 9 - M Alharbi et al | 0 | 41 | 1 | 24 | 0.19 | 0.01 | 4.82 | 0.46 | | | | |
| PIMS-TS/MIS-C | 49 | 151 | 35 | 95 | 0.80 | 0.46 | 1.41 | 13.85 | 0.73 | 0.00 | 0.75 | 0.45 |
| 10 - Z Almoussa et al | 1 | 5 | 0 | 5 | 3.67 | 0.12 | 113.73 | 0.41 | | | | |
| 12 - OY Antunez-Montes et al | 15 | 43 | 9 | 24 | 0.89 | 0.32 | 2.52 | 4.10 | | | | |
| 13 - AR Araujo da Silva et al | 1 | 5 | 1 | 9 | 2.00 | 0.10 | 41.00 | 0.52 | | | | |
| 143 - M Riollano-Cruz et al | 3 | 11 | 0 | 4 | 3.71 | 0.15 | 88.75 | 0.47 | | | | |
| 147 - L Shahbaznejad et al | 2 | 6 | 1 | 4 | 1.50 | 0.09 | 25.39 | 0.60 | | | | |
| 167 - E Whittaker et al | 15 | 38 | 10 | 20 | 0.65 | 0.22 | 1.94 | 3.73 | | | | |
| 179 - J Pang et al | 3 | 3 | 1 | 2 | 7.00 | 0.17 | 291.34 | 0.34 | | | | |
| 183 - Rekhtman et al | 2 | 13 | 3 | 6 | 0.18 | 0.02 | 1.64 | 0.98 | | | | |

| Name | Events 1 | Total 1 | Events 2 | Total 2 | Effect Estimate | CI Start | CI End | Weight | P(Q) | I²(Q) | Z | P(Z) |
|---|---|---|---|---|---|---|---|---|---|---|---|---|
| 187 - R Carbajal et al | 0 | 2 | 3 | 5 | 0.14 | 0.00 | 4.47 | 0.40 | | | | |
| 244- Luna-Munoz | 2 | 7 | 1 | 3 | 0.80 | 0.04 | 14.64 | 0.5 | | | | |
| 228 - Hasan et al | 1 | 5 | 0 | 2 | 1.67 | 0.05 | 58.28 | 0.38 | | | | |
| 41 - RL Crawford et al | 0 | 5 | 0 | 0 | 0.00 | 0.00 | 0.00 | 0.00 | | | | |
| 89 - S Jain et al | 3 | 11 | 6 | 12 | 0.38 | 0.07 | 2.14 | 1.54 | | | | |
| 9 - M Alharbi et al | 3 | 4 | 1 | 2 | 3.00 | 0.08 | 107.45 | 0.37 | | | | |
| Need for inotropes/ECMO | 69 | 1009 | 54 | 828 | 0.82 | 0.47 | 1.46 | 100.00 | 0.26 | 17.85 | 0.66 | 0.51 |
| COVID-19 | 20 | 895 | 13 | 756 | 1.32 | 0.66 | 2.64 | 45.65 | 0.80 | 0.00 | 0.77 | 0.44 |
| 12 - OY Antunez-Montes et al | 1 | 110 | 0 | 71 | 1.96 | 0.08 | 48.76 | 2.95 | | | | |
| 142 - S Richardson et al | 0 | 51 | 0 | 59 | 0.00 | 0.00 | 0.00 | 0.00 | | | | |
| 15 - JP Armann et al | 5 | 45 | 3 | 57 | 2.25 | 0.51 | 9.97 | 11.16 | | | | |
| 28 - H Cairoli | 1 | 315 | 0 | 263 | 2.51 | 0.10 | 61.96 | 2.97 | | | | |
| 46 - M Ceano-Vivas et al | 1 | 22 | 0 | 11 | 1.60 | 0.06 | 42.63 | 2.84 | | | | |
| 79 - F Gotzinger et al | 11 | 311 | 8 | 271 | 1.21 | 0.48 | 3.04 | 20.90 | | | | |
| 9 - M Alharbi et al | 1 | 41 | 2 | 24 | 0.28 | 0.02 | 3.21 | 4.83 | | | | |
| PIMS-TS/MIS-C | 49 | 114 | 41 | 72 | 0.53 | 0.21 | 1.35 | 54.35 | 0.15 | 36.60 | 1.33 | 0.18 |
| 10 - Z Almoussa et al | 2 | 5 | 3 | 5 | 0.44 | 0.04 | 5.58 | 4.58 | | | | |
| 12 - OY Antunez-Montes et al | 16 | 43 | 7 | 24 | 1.44 | 0.49 | 4.22 | 17.52 | | | | |
| 147 - L Shahbaznejad et al | 2 | 6 | 2 | 4 | 0.50 | 0.04 | 6.68 | 4.39 | | | | |
| 167 - E Whittaker et al | 14 | 38 | 13 | 20 | 0.31 | 0.10 | 0.97 | 16.44 | | | | |
| 187 - R Carbajal et al | 0 | 2 | 5 | 5 | 0.02 | 0.00 | 1.21 | 1.78 | | | | |
| 41 - RL Crawford et al | 5 | 5 | 0 | 0 | 0.00 | 0.00 | 0.00 | 0.00 | | | | |
| 89 - S Jain et al | 6 | 11 | 10 | 12 | 0.24 | 0.03 | 1.65 | 7.37 | | | | |
| 9 - M Alharbi et al | 4 | 4 | 1 | 2 | 9.00 | 0.22 | 362.48 | 2.27 | | | | |
| Admission to Critical Care / ICU | 2088 | 7878 | 1745 | 7300 | 1.19 | 0.99 | 1.44 | 100.00 | 0.00 | 44.79 | 1.81 | 0.07 |
| COVID-19 | 1622 | 7121 | 1401 | 6716 | 1.19 | 0.93 | 1.52 | 77.12 | 0.00 | 58.55 | 1.41 | 0.16 |
| 122 - A Moreira et al | 63 | 206 | 42 | 239 | 2.07 | 1.32 | 3.23 | 8.54 | | | | |
| 12 - OY Antunez-Montes et al | 8 | 110 | 3 | 71 | 1.78 | 0.46 | 6.94 | 1.75 | | | | |
| 136 - G Qian et al | 5 | 86 | 2 | 39 | 1.14 | 0.21 | 6.16 | 1.18 | | | | |
| 13 - AR Araujo da Silva et al | 15 | 34 | 11 | 15 | 0.29 | 0.08 | 1.09 | 1.82 | | | | |

| Name | Events 1 | Total 1 | Events 2 | Total 2 | Effect Estimate | CI Start | CI End | Weight | P(Q) | I²(Q) | Z | P(Z) |
|---|---|---|---|---|---|---|---|---|---|---|---|---|
| 142 - S Richardson et al | 8 | 51 | 6 | 59 | 1.64 | 0.53 | 5.10 | 2.41 | | | | |
| 151 - B Sousa et al | 981 | 3591 | 885 | 3341 | 1.04 | 0.94 | 1.16 | 15.28 | | | | |
| 155 - Kyung Sung | 0 | 54 | 0 | 47 | 0.00 | 0.00 | 0.00 | 0.00 | | | | |
| 157 - O Swann et al | 49 | 326 | 29 | 253 | 1.37 | 0.84 | 2.23 | 7.76 | | | | |
| 15 - JP Armann et al | 5 | 45 | 10 | 57 | 0.59 | 0.19 | 1.86 | 2.33 | | | | |
| 21 - D Bayesheva et al | 0 | 311 | 4 | 241 | 0.08 | 0.00 | 1.58 | 0.41 | | | | |
| 221 - Preston et al | 402 | 1083 | 345 | 1344 | 1.71 | 1.44 | 2.03 | 14.15 | | | | |
| 224 - Graff et al | 6 | 39 | 5 | 27 | 0.80 | 0.22 | 2.95 | 1.89 | | | | |
| 26 - CE Bolaños-Almeida et al | 8 | 317 | 9 | 283 | 0.79 | 0.30 | 2.07 | 3.13 | | | | |
| 28 - H Cairoli | 3 | 315 | 0 | 263 | 5.90 | 0.30 | 114.79 | 0.40 | | | | |
| 32 - J Chao et al | 8 | 31 | 5 | 15 | 0.70 | 0.18 | 2.66 | 1.79 | | | | |
| 46 - M Ceano-Vivas et al | 3 | 22 | 2 | 11 | 0.71 | 0.10 | 5.03 | 0.90 | | | | |
| 67 - G Fisler et al | 13 | 33 | 17 | 40 | 0.88 | 0.34 | 2.25 | 3.28 | | | | |
| 75 - V Giacomet et al | 5 | 82 | 3 | 44 | 0.89 | 0.20 | 3.90 | 1.50 | | | | |
| 79 - F Gotzinger et al | 33 | 311 | 15 | 271 | 2.03 | 1.08 | 3.82 | 5.79 | | | | |
| 91 - M Kainth et al | 4 | 33 | 5 | 32 | 0.74 | 0.18 | 3.07 | 1.63 | | | | |
| 9 - M Alharbi et al | 3 | 41 | 3 | 24 | 0.55 | 0.10 | 2.99 | 1.18 | | | | |
| PIMS-TS/MIS-C | 466 | 757 | 344 | 584 | 1.15 | 0.92 | 1.43 | 22.88 | 0.69 | 0.00 | 1.19 | 0.23 |
| 10 - Z Almoussa et al | 5 | 5 | 4 | 5 | 3.67 | 0.12 | 113.73 | 0.30 | | | | |
| 12 - OY Antunez-Montes et al | 17 | 43 | 11 | 24 | 0.77 | 0.28 | 2.12 | 2.92 | | | | |
| 13 - AR Araujo da Silva et al | 5 | 5 | 7 | 9 | 3.67 | 0.15 | 92.65 | 0.34 | | | | |
| 147 - L Shahbaznejad et al | 5 | 6 | 4 | 4 | 0.41 | 0.01 | 12.64 | 0.30 | | | | |
| 160 - JP Torres et al | 9 | 14 | 5 | 13 | 2.88 | 0.60 | 13.75 | 1.36 | | | | |
| 167 - E Whittaker et al | 25 | 38 | 13 | 20 | 1.04 | 0.33 | 3.23 | 2.39 | | | | |
| 183 - Rekhtman et al | 8 | 13 | 4 | 6 | 0.80 | 0.10 | 6.10 | 0.83 | | | | |
| 187 - R Carbajal et al | 2 | 2 | 5 | 5 | 0.00 | 0.00 | 0.00 | 0.00 | | | | |
| 226 - Abrams et al | 372 | 602 | 275 | 476 | 1.18 | 0.93 | 1.51 | 12.67 | | | | |
| 228 - Hasan et al | 2 | 5 | 0 | 2 | 3.57 | 0.11 | 111.71 | 0.30 | | | | |
| 41 - RL Crawford et al | 4 | 5 | 0 | 0 | 0.00 | 0.00 | 0.00 | 0.00 | | | | |
| 5 - N Abdel-Haq et al | 8 | 15 | 14 | 18 | 0.33 | 0.07 | 1.47 | 1.46 | | | | |

| Name | Events 1 | Total 1 | Events 2 | Total 2 | Effect Estimate | CI Start | CI End | Weight | P(Q) | I²(Q) | Z | P(Z) |
|---|---|---|---|---|---|---|---|---|---|---|---|---|
| 9 - M Alharbi et al | 4 | 4 | 2 | 2 | 0.00 | 0.00 | 0.00 | 0.00 | | | | |
| Received Pharmacological Therapy | 161 | 640 | 109 | 498 | 0.71 | 0.44 | 1.15 | 100.00 | 0.82 | 0.00 | 1.40 | 0.16 |
| COVID-19 | 57 | 526 | 38 | 417 | 0.73 | 0.42 | 1.27 | 77.48 | 0.63 | 0.00 | 1.12 | 0.26 |
| 12 - OY Antunez-Montes et al | 11 | 110 | 5 | 71 | 0.68 | 0.23 | 2.05 | 19.05 | | | | |
| 13 - AR Araujo da Silva et al | 22 | 34 | 6 | 15 | 0.36 | 0.10 | 1.27 | 14.81 | | | | |
| 15 - JP Armann et al | 20 | 45 | 26 | 57 | 1.05 | 0.48 | 2.30 | 37.47 | | | | |
| 28 - H Cairoli | 2 | 315 | 0 | 263 | 0.24 | 0.01 | 4.98 | 2.50 | | | | |
| 46 - M Ceano-Vivas et al | 2 | 22 | 1 | 11 | 1.00 | 0.08 | 12.40 | 3.65 | | | | |
| PIMS-TS/MIS-C | 104 | 114 | 71 | 81 | 0.63 | 0.23 | 1.74 | 22.52 | 0.63 | 0.00 | 0.89 | 0.37 |
| 10 - Z Almoussa et al | 5 | 5 | 5 | 5 | 0.00 | 0.00 | 0.00 | 0.00 | | | | |
| 12 - OY Antunez-Montes et al | 42 | 43 | 23 | 24 | 0.55 | 0.03 | 9.17 | 2.91 | | | | |
| 13 - AR Araujo da Silva et al | 5 | 5 | 6 | 9 | 0.17 | 0.01 | 4.03 | 2.30 | | | | |
| 147 - L Shahbaznejad et al | 6 | 6 | 4 | 4 | 0.00 | 0.00 | 0.00 | 0.00 | | | | |
| 167 - E Whittaker et al | 30 | 38 | 14 | 20 | 0.62 | 0.18 | 2.14 | 15.19 | | | | |
| 187 - R Carbajal et al | 2 | 2 | 5 | 5 | 0.00 | 0.00 | 0.00 | 0.00 | | | | |
| 89 - S Jain et al | 10 | 11 | 12 | 12 | 3.57 | 0.13 | 97.23 | 2.12 | | | | |
| 9 - M Alharbi et al | 4 | 4 | 2 | 2 | 0.00 | 0.00 | 0.00 | 0.00 | | | | |
| Death | 620 | 10921 | 651 | 10491 | 0.94 | 0.84 | 1.05 | 100.00 | 0.99 | 0.00 | 1.08 | 0.28 |
| COVID-19 | 610 | 10758 | 647 | 10387 | 0.94 | 0.83 | 1.05 | 99.00 | 0.98 | 0.00 | 1.09 | 0.27 |
| 122 - A Moreira et al | 4 | 206 | 8 | 239 | 0.57 | 0.17 | 1.93 | 0.90 | | | | |
| 12 - OY Antunez-Montes et al | 0 | 11 | 0 | 5 | 0.00 | 0.00 | 0.00 | 0.00 | | | | |
| 13 - AR Araujo da Silva et al | 1 | 34 | 0 | 15 | 1.39 | 0.05 | 36.04 | 0.13 | | | | |
| 142 - S Richardson et al | 0 | 51 | 1 | 59 | 0.38 | 0.02 | 9.50 | 0.13 | | | | |
| 151 - B Sousa et al | 286 | 3591 | 278 | 3341 | 0.95 | 0.80 | 1.13 | 44.75 | | | | |
| 155 - Kyung Sung | 0 | 54 | 0 | 47 | 0.00 | 0.00 | 0.00 | 0.00 | | | | |
| 15 - JP Armann et al | 0 | 45 | 1 | 57 | 0.41 | 0.02 | 10.40 | 0.13 | | | | |
| 19 - S Bellino et al | 1 | 1970 | 2 | 1866 | 0.47 | 0.04 | 5.22 | 0.23 | | | | |
| 21 - D Bayesheva et al | 0 | 311 | 0 | 241 | 0.00 | 0.00 | 0.00 | 0.00 | | | | |
| 26 - CE Bolaños-Almeida et al | 2 | 317 | 3 | 283 | 0.59 | 0.10 | 3.57 | 0.41 | | | | |

| Name | Events 1 | Total 1 | Events 2 | Total 2 | Effect Estimate | CI Start | CI End | Weight | P(Q) | I²(Q) | Z | P(Z) |
|---|---|---|---|---|---|---|---|---|---|---|---|---|
| 28 - H Cairoli | 1 | 315 | 1 | 263 | 0.83 | 0.05 | 13.40 | 0.17 | | | | |
| 46 - M Ceano-Vivas et al | 1 | 22 | 0 | 11 | 1.60 | 0.06 | 42.63 | 0.12 | | | | |
| 75 - V Giacomet et al | 0 | 82 | 0 | 44 | 0.00 | 0.00 | 0.00 | 0.00 | | | | |
| 79 - F Gotzinger et al | 3 | 311 | 1 | 271 | 2.63 | 0.27 | 25.43 | 0.26 | | | | |
| 86 - D Hillesheim et al | 311 | 3383 | 350 | 3604 | 0.94 | 0.80 | 1.11 | 51.53 | | | | |
| 92 - R Marcello et al | 0 | 14 | 1 | 17 | 0.38 | 0.01 | 10.05 | 0.12 | | | | |
| 9 - M Alharbi et al | 0 | 41 | 1 | 24 | 0.19 | 0.01 | 4.82 | 0.13 | | | | |
| PIMS-TS/MIS-C | 10 | 163 | 4 | 104 | 1.07 | 0.34 | 3.38 | 1.00 | 0.72 | 0.00 | 0.11 | 0.91 |
| 10 - Z Almoussa et al | 1 | 5 | 1 | 5 | 1.00 | 0.05 | 22.18 | 0.14 | | | | |
| 12 - OY Antunez-Montes et al | 4 | 43 | 0 | 24 | 5.58 | 0.29 | 108.26 | 0.15 | | | | |
| 13 - AR Araujo da Silva et al | 0 | 5 | 0 | 9 | 0.00 | 0.00 | 0.00 | 0.00 | | | | |
| 143 - M Riollano-Cruz et al | 1 | 11 | 0 | 4 | 1.29 | 0.04 | 37.98 | 0.12 | | | | |
| 147 - L Shahbaznejad et al | 2 | 6 | 0 | 4 | 5.00 | 0.18 | 136.32 | 0.12 | | | | |
| 167 - E Whittaker et al | 1 | 38 | 0 | 20 | 1.64 | 0.06 | 42.11 | 0.13 | | | | |
| 183 - Rekhtman et al | 0 | 13 | 1 | 6 | 0.14 | 0.00 | 3.87 | 0.12 | | | | |
| 187 - R Carbajal et al | 0 | 2 | 0 | 5 | 0.00 | 0.00 | 0.00 | 0.00 | | | | |
| 29 - C Capone et al | 0 | 20 | 0 | 13 | 0.00 | 0.00 | 0.00 | 0.00 | | | | |
| 41 - RL Crawford et al | 0 | 5 | 0 | 0 | 0.00 | 0.00 | 0.00 | 0.00 | | | | |
| 89 - S Jain et al | 0 | 11 | 1 | 12 | 0.33 | 0.01 | 9.07 | 0.12 | | | | |
| 9 - M Alharbi et al | 1 | 4 | 1 | 2 | 0.33 | 0.01 | 11.94 | 0.10 | | | | |
| Any co-morbidity | | | | | | | | | | | | |
| Invasive Ventilation | 370 | 2623 | 514 | 7068 | 2.29 | 1.36 | 3.86 | 100.00 | 0.10 | 31.20 | 3.10 | 0.00 |
| COVID-19 | 355 | 2580 | 466 | 6931 | 2.73 | 1.62 | 4.62 | 74.09 | 0.22 | 22.87 | 3.76 | 0.00 |
| 12 - OY Antunez-Montes et al | 0 | 60 | 3 | 121 | 0.28 | 0.01 | 5.51 | 2.77 | | | | |
| 13 - AR Araujo da Silva et al | 4 | 31 | 0 | 19 | 6.38 | 0.32 | 125.49 | 2.77 | | | | |
| 142 - S Richardson et al | 2 | 14 | 12 | 96 | 1.17 | 0.23 | 5.86 | 7.52 | | | | |
| 151 - B Sousa et al | 318 | 1957 | 437 | 4979 | 2.02 | 1.73 | 2.35 | 25.78 | | | | |
| 155 - Kyung Sung | 0 | 5 | 0 | 96 | 0.00 | 0.00 | 0.00 | 0.00 | | | | |
| 15 - JP Armann et al | 6 | 33 | 0 | 69 | 32.85 | 1.79 | 603.18 | 2.89 | | | | |
| 170 - B Yayla et al | 0 | 5 | 1 | 72 | 4.33 | 0.16 | 119.39 | 2.28 | | | | |

| Name | Events 1 | Total 1 | Events 2 | Total 2 | Effect Estimate | CI Start | CI End | Weight | P(Q) | I²(Q) | Z | P(Z) |
|---|---|---|---|---|---|---|---|---|---|---|---|---|
| 174 - P Zachariah et al | 8 | 33 | 1 | 17 | 5.12 | 0.58 | 44.91 | 4.77 | | | | |
| 21 - D Bayesheva et al | 0 | 62 | 1 | 490 | 2.61 | 0.11 | 64.78 | 2.42 | | | | |
| 28 - H Cairoli | 1 | 191 | 0 | 387 | 6.10 | 0.25 | 150.50 | 2.43 | | | | |
| 75 - V Giacomet et al | 0 | 20 | 1 | 107 | 1.73 | 0.07 | 44.01 | 2.38 | | | | |
| 79 - F Gotzinger et al | 16 | 145 | 9 | 437 | 5.90 | 2.55 | 13.66 | 15.72 | | | | |
| 9 - M Alharbi et al | 0 | 24 | 1 | 41 | 0.55 | 0.02 | 14.07 | 2.38 | | | | |
| PIMS-TS/MIS-C | 15 | 43 | 48 | 137 | 0.83 | 0.22 | 3.09 | 25.91 | 0.06 | 51.03 | 0.28 | 0.78 |
| 10 - Z Almoussa et al | 0 | 4 | 1 | 6 | 0.41 | 0.01 | 12.64 | 2.14 | | | | |
| 12 - OY Antunez-Montes et al | 11 | 17 | 13 | 50 | 5.22 | 1.61 | 16.96 | 11.30 | | | | |
| 13 - AR Araujo da Silva et al | 0 | 5 | 2 | 9 | 0.27 | 0.01 | 6.89 | 2.39 | | | | |
| 143 - M Riollano-Cruz et al | 1 | 5 | 2 | 10 | 1.00 | 0.07 | 14.64 | 3.33 | | | | |
| 167 - E Whittaker et al | 0 | 6 | 25 | 52 | 0.08 | 0.00 | 1.55 | 2.86 | | | | |
| 179 - J Pang et al | 1 | 2 | 3 | 3 | 0.14 | 0.00 | 5.95 | 1.84 | | | | |
| 244 - Luna-Munoz et al | 0 | 1 | 3 | 9 | 0.62 | 0.02 | 19.5 | 2.00 | | | | |
| 41 - RL Crawford et al | 0 | 2 | 0 | 3 | 0.00 | 0.00 | 0.00 | 0.00 | | | | |
| 9 - M Alharbi et al | 2 | 2 | 2 | 4 | 5.00 | 0.15 | 166.59 | 2.06 | | | | |
| Cardiovascular support | 29 | 338 | 23 | 753 | 4.50 | 2.13 | 9.51 | 100.00 | 0.91 | 0.00 | 3.93 | 0.00 |
| COVID-19 | 9 | 313 | 6 | 690 | 4.07 | 1.43 | 11.53 | 51.65 | 0.68 | 0.00 | 2.64 | 0.01 |
| 12 - OY Antunez-Montes et al | 0 | 60 | 1 | 121 | 0.66 | 0.03 | 16.54 | 5.43 | | | | |
| 15 - JP Armann et al | 5 | 33 | 3 | 69 | 3.93 | 0.88 | 17.57 | 25.01 | | | | |
| 170 - B Yayla et al | 1 | 5 | 1 | 72 | 17.75 | 0.93 | 338.84 | 6.45 | | | | |
| 28 - H Cairoli | 1 | 191 | 0 | 387 | 6.10 | 0.25 | 150.50 | 5.46 | | | | |
| 9 - M Alharbi et al | 2 | 24 | 1 | 41 | 3.64 | 0.31 | 42.40 | 9.30 | | | | |
| PIMS-TS/MIS-C | 20 | 25 | 17 | 63 | 5.01 | 1.71 | 14.71 | 48.35 | 0.84 | 0.00 | 2.93 | 0.00 |
| 10 - Z Almoussa et al | 5 | 4 | 0 | 6 | 0.00 | 0.00 | 0.00 | 0.00 | | | | |
| 12 - OY Antunez-Montes et al | 11 | 17 | 12 | 50 | 5.81 | 1.77 | 19.04 | 39.78 | | | | |
| 41 - RL Crawford et al | 2 | 2 | 2 | 3 | 3.00 | 0.08 | 115.34 | 4.21 | | | | |
| 9 - M Alharbi et al | 2 | 2 | 3 | 4 | 2.14 | 0.06 | 77.54 | 4.36 | | | | |
| Admission to Critical Care | 1485 | 4570 | 1474 | 8158 | 2.55 | 1.79 | 3.64 | 100.00 | 0.00 | 70.28 | 5.14 | 0.00 |
| COVID-19 | 1444 | 4516 | 1434 | 8070 | 2.56 | 1.77 | 3.71 | 86.92 | 0.00 | 74.20 | 5.01 | 0.00 |

| Name | Events 1 | Total 1 | Events 2 | Total 2 | Effect Estimate | CI Start | CI End | Weight | P(Q) | I²(Q) | Z | P(Z) |
|---|---|---|---|---|---|---|---|---|---|---|---|---|
| 12 - OY Antunez-Montes et al | 2 | 60 | 9 | 121 | 0.43 | 0.09 | 2.05 | 3.80 | | | | |
| 13 - AR Araujo da Silva et al | 18 | 31 | 8 | 19 | 1.90 | 0.60 | 6.05 | 5.68 | | | | |
| 142 - S Richardson et al | 7 | 14 | 30 | 96 | 2.20 | 0.71 | 6.83 | 5.82 | | | | |
| 151 - B Sousa et al | 673 | 1957 | 1194 | 4979 | 1.66 | 1.48 | 1.86 | 13.91 | | | | |
| 155 - Kyung Sung | 0 | 5 | 0 | 96 | 0.00 | 0.00 | 0.00 | 0.00 | | | | |
| 157 - O Swann et al | 53 | 258 | 25 | 322 | 3.07 | 1.85 | 5.10 | 10.97 | | | | |
| 15 - JP Armann et al | 10 | 33 | 5 | 69 | 5.57 | 1.72 | 18.01 | 5.58 | | | | |
| 163 - S Verma et al | 16 | 38 | 7 | 44 | 3.84 | 1.37 | 10.80 | 6.46 | | | | |
| 170 - B Yayla et al | 0 | 5 | 1 | 72 | 4.33 | 0.16 | 119.39 | 1.07 | | | | |
| 21 - D Bayesheva et al | 0 | 62 | 4 | 490 | 0.86 | 0.05 | 16.26 | 1.34 | | | | |
| 221 - Preston et al | 630 | 1659 | 117 | 771 | 3.42 | 2.75 | 4.27 | 13.39 | | | | |
| 28 - H Cairoli | 3 | 191 | 0 | 387 | 14.39 | 0.74 | 280.01 | 1.31 | | | | |
| 46 - M Ceano-Vivas et al | 1 | 14 | 4 | 19 | 0.29 | 0.03 | 2.92 | 2.03 | | | | |
| 75 - V Giacomet et al | 2 | 20 | 5 | 107 | 2.27 | 0.41 | 12.59 | 3.31 | | | | |
| 79 - F Gotzinger et al | 24 | 145 | 24 | 437 | 3.41 | 1.87 | 6.23 | 10.07 | | | | |
| 9 - M Alharbi et al | 5 | 24 | 1 | 41 | 10.53 | 1.15 | 96.47 | 2.19 | | | | |
| PIMS-TS/MIS-C | 41 | 54 | 40 | 88 | 1.93 | 0.41 | 8.97 | 13.08 | 0.03 | 59.89 | 0.83 | 0.40 |
| 10 - Z Almoussa et al | 4 | 4 | 5 | 6 | 2.45 | 0.08 | 76.13 | 1.00 | | | | |
| 12 - OY Antunez-Montes et al | 12 | 17 | 16 | 50 | 5.10 | 1.54 | 16.94 | 5.43 | | | | |
| 13 - AR Araujo da Silva et al | 5 | 9 | 7 | 9 | 0.36 | 0.05 | 2.77 | 2.49 | | | | |
| 41 - RL Crawford et al | 2 | 3 | 2 | 3 | 1.00 | 0.03 | 29.81 | 1.02 | | | | |
| 5 - N Abdel-Haq et al | 16 | 17 | 6 | 16 | 26.67 | 2.78 | 255.46 | 2.12 | | | | |
| 9 - M Alharbi et al | 2 | 4 | 4 | 4 | 0.11 | 0.00 | 3.35 | 1.02 | | | | |
| Death | 284 | 3045 | 312 | 10873 | 3.22 | 1.87 | 5.54 | 100.00 | 0.34 | 9.76 | 4.23 | 0.00 |
| COVID-19 | 278 | 3004 | 308 | 10739 | 4.16 | 1.97 | 8.80 | 85.40 | 0.23 | 22.11 | 3.46 | 0.00 |
| 12 - OY Antunez-Montes et al | 1 | 60 | 0 | 121 | 6.13 | 0.25 | 152.65 | 2.71 | | | | |
| 13 - AR Araujo da Silva et al | 1 | 31 | 0 | 19 | 1.92 | 0.07 | 49.50 | 2.65 | | | | |
| 151 - B Sousa et al | 261 | 1957 | 303 | 4979 | 2.37 | 1.99 | 2.83 | 54.62 | | | | |
| 155 - Kyung Sung | 0 | 5 | 0 | 96 | 0.00 | 0.00 | 0.00 | 0.00 | | | | |
| 157 - O Swann et al | 6 | 276 | 0 | 375 | 18.05 | 1.01 | 321.70 | 3.33 | | | | |

| Name | Events 1 | Total 1 | Events 2 | Total 2 | Effect Estimate | CI Start | CI End | Weight | P(Q) | I²(Q) | Z | P(Z) |
|---|---|---|---|---|---|---|---|---|---|---|---|---|
| 15 - JP Armann et al | 1 | 33 | 0 | 69 | 6.42 | 0.25 | 161.79 | 2.69 | | | | |
| 170 - B Yayla et al | 0 | 5 | 1 | 72 | 4.33 | 0.16 | 119.39 | 2.55 | | | | |
| 19 - S Bellino et al | 4 | 206 | 0 | 3630 | 161.36 | 8.66 | 3007.24 | 3.24 | | | | |
| 21 - D Bayesheva et al | 0 | 62 | 0 | 490 | 0.00 | 0.00 | 0.00 | 0.00 | | | | |
| 239 - Qian et al | 1 | 8 | 1 | 119 | 16.86 | 0.95 | 298.73 | 3.7 | | | | |
| 28 - H Cairoli | 2 | 191 | 0 | 387 | 10.22 | 0.49 | 214.03 | 3.01 | | | | |
| 79 - F Gotzinger et al | 2 | 145 | 2 | 437 | 3.04 | 0.42 | 21.79 | 6.70 | | | | |
| 92 - R Marcello et al | 0 | 9 | 1 | 23 | 0.79 | 0.03 | 21.18 | 2.59 | | | | |
| 9 - M Alharbi et al | 0 | 24 | 1 | 41 | 0.55 | 0.02 | 14.07 | 2.67 | | | | |
| PIMS-TS/MIS-C | 6 | 41 | 4 | 134 | 3.36 | 0.79 | 14.40 | 13.24 | 0.46 | 0.00 | 1.64 | 0.10 |
| 10 - Z Almoussa et al | 1 | 4 | 1 | 6 | 1.67 | 0.07 | 37.73 | 2.87 | | | | |
| 12 - OY Antunez-Montes et al | 4 | 17 | 0 | 50 | 33.67 | 1.71 | 664.69 | 3.12 | | | | |
| 13 - AR Araujo da Silva et al | 0 | 5 | 0 | 9 | 0.00 | 0.00 | 0.00 | 0.00 | | | | |
| 143 - M Riollano-Cruz et al | 0 | 5 | 1 | 10 | 0.58 | 0.02 | 16.72 | 2.48 | | | | |
| 167 - E Whittaker et al | 0 | 6 | 1 | 52 | 2.64 | 0.10 | 71.82 | 2.57 | | | | |
| 41 - RL Crawford et al | 0 | 2 | 0 | 3 | 0.00 | 0.00 | 0.00 | 0.00 | | | | |
| 9 - M Alharbi et al | 1 | 2 | 1 | 4 | 3.00 | 0.08 | 107.45 | 2.21 | | | | |
| Cardiovascular co-morbidity | | | | | | | | | | | | |
| Invasive Ventilation | 87 | 373 | 737 | 9021 | 6.20 | 2.85 | 13.49 | 100.00 | 0.04 | 48.12 | 4.61 | 0.00 |
| COVID-19 | 87 | 372 | 735 | 9008 | 6.74 | 2.97 | 15.29 | 95.76 | 0.02 | 52.83 | 4.56 | 0.00 |
| 13 - AR Araujo da Silva et al | 1 | 2 | 3 | 48 | 15.00 | 0.74 | 303.74 | 5.41 | | | | |
| 142 - S Richardson et al | 1 | 2 | 13 | 108 | 7.31 | 0.43 | 124.05 | 5.96 | | | | |
| 151 - B Sousa et al | 66 | 273 | 689 | 6663 | 2.76 | 2.07 | 3.69 | 27.56 | | | | |
| 15 - JP Armann et al | 1 | 9 | 5 | 93 | 2.20 | 0.23 | 21.20 | 8.33 | | | | |
| 174 - P Zachariah et al | 1 | 4 | 8 | 46 | 1.58 | 0.15 | 17.25 | 7.73 | | | | |
| 21 - D Bayesheva et al | 0 | 7 | 1 | 545 | 24.20 | 0.91 | 643.60 | 4.69 | | | | |
| 28 - H Cairoli | 0 | 4 | 1 | 574 | 42.48 | 1.52 | 1189.60 | 4.57 | | | | |
| 54 - A Desai et al | 14 | 41 | 14 | 252 | 8.81 | 3.80 | 20.44 | 21.44 | | | | |
| 75 - V Giacomet et al | 0 | 5 | 1 | 122 | 7.36 | 0.27 | 202.20 | 4.61 | | | | |
| 79 - F Gotzinger et al | 3 | 25 | 0 | 557 | 173.44 | 8.70 | 3459.45 | 5.45 | | | | |

| Name | Events 1 | Total 1 | Events 2 | Total 2 | Effect Estimate | CI Start | CI End | Weight | P(Q) | I²(Q) | Z | P(Z) |
|---|---|---|---|---|---|---|---|---|---|---|---|---|
| PIMS-TS/MIS-C | 0 | 1 | 2 | 13 | 1.53 | 0.05 | 49.80 | 4.24 | 1.00 | 0.00 | 0.24 | 0.81 |
| 13 - AR Araujo da Silva et al | 0 | 1 | 2 | 13 | 1.53 | 0.05 | 49.80 | 4.24 | | | | |
| Cardiovascular support | 6 | 46 | 30 | 1291 | 5.21 | 2.05 | 13.23 | 100.00 | 0.63 | 0.00 | 3.47 | 0.00 |
| COVID-19 | 6 | 46 | 25 | 1281 | 5.21 | 2.05 | 13.23 | 100.00 | 0.63 | 0.00 | 3.47 | 0.00 |
| 15 - JP Armann et al | 2 | 9 | 6 | 93 | 4.14 | 0.70 | 24.47 | 27.54 | | | | |
| 28 - H Cairoli | 0 | 4 | 1 | 574 | 42.48 | 1.52 | 1189.60 | 7.82 | | | | |
| 79 - F Gotzinger et al | 3 | 25 | 16 | 557 | 4.61 | 1.25 | 17.00 | 51.03 | | | | |
| 9 - M Alharbi et al | 1 | 8 | 2 | 57 | 3.93 | 0.31 | 49.12 | 13.61 | | | | |
| PIMS-TS/MIS-C | 0 | 0 | 5 | 10 | 0.00 | 0.00 | 0.00 | 0.00 | 0.00 | 0.00 | 0.00 | 0.00 |
| 10 - Z Almoussa et al | 0 | 0 | 5 | 10 | 0.00 | 0.00 | 0.00 | 0.00 | | | | |
| Admission to critical care | 166 | 385 | 2020 | 9708 | 2.87 | 2.32 | 3.55 | 100.00 | 0.55 | 0.00 | 9.66 | 0.00 |
| COVID-19 | 165 | 384 | 2009 | 9695 | 2.88 | 2.33 | 3.57 | 99.62 | 0.53 | 0.00 | 9.69 | 0.00 |
| 136 - G Qian et al | 2 | 3 | 5 | 124 | 47.60 | 3.67 | 616.85 | 0.70 | | | | |
| 13 - AR Araujo da Silva et al | 1 | 2 | 25 | 48 | 0.92 | 0.05 | 15.58 | 0.57 | | | | |
| 142 - S Richardson et al | 2 | 2 | 35 | 108 | 10.35 | 0.48 | 221.38 | 0.49 | | | | |
| 151 - B Sousa et al | 129 | 273 | 1738 | 6663 | 2.54 | 1.99 | 3.24 | 76.92 | | | | |
| 157 - O Swann et al | 12 | 36 | 66 | 544 | 3.62 | 1.73 | 7.58 | 8.37 | | | | |
| 15 - JP Armann et al | 4 | 9 | 11 | 93 | 5.96 | 1.39 | 25.62 | 2.15 | | | | |
| 163 - S Verma et al | 1 | 1 | 22 | 81 | 7.93 | 0.31 | 201.99 | 0.44 | | | | |
| 21 - D Bayesheva et al | 0 | 7 | 4 | 545 | 8.02 | 0.40 | 162.64 | 0.50 | | | | |
| 28 - H Cairoli | 0 | 4 | 3 | 574 | 18.14 | 0.81 | 404.74 | 0.47 | | | | |
| 32 - J Chao et al | 1 | 1 | 12 | 45 | 8.04 | 0.31 | 210.67 | 0.43 | | | | |
| 67 - G Fisler et al | 3 | 5 | 27 | 72 | 2.50 | 0.39 | 15.93 | 1.33 | | | | |
| 75 - V Giacomet et al | 1 | 5 | 7 | 122 | 4.11 | 0.40 | 41.81 | 0.85 | | | | |
| 79 - F Gotzinger et al | 5 | 25 | 43 | 557 | 2.99 | 1.07 | 8.36 | 4.33 | | | | |
| 91 - M Kainth et al | 1 | 3 | 8 | 62 | 3.38 | 0.27 | 41.64 | 0.72 | | | | |
| 9 - M Alharbi et al | 3 | 8 | 3 | 57 | 10.80 | 1.71 | 68.28 | 1.34 | | | | |
| PIMS-TS/MIS-C | 1 | 1 | 11 | 13 | 0.65 | 0.02 | 21.18 | 0.38 | 1.00 | 0.00 | 0.24 | 0.81 |
| 13 - AR Araujo da Silva et al | 1 | 1 | 11 | 13 | 0.65 | 0.02 | 21.18 | 0.38 | | | | |
| Death | 65 | 344 | 513 | 12479 | 11.33 | 2.60 | 49.33 | 100.00 | 0.00 | 69.24 | 3.23 | 0.00 |

| Name | Events 1 | Total 1 | Events 2 | Total 2 | Effect Estimate | CI Start | CI End | Weight | P(Q) | I²(Q) | Z | P(Z) |
|---|---|---|---|---|---|---|---|---|---|---|---|---|
| COVID-19 | 65 | 342 | 512 | 12436 | 12.05 | 2.37 | 61.34 | 90.58 | 0.00 | 73.55 | 3.00 | 0.00 |
| 13 - AR Araujo da Silva et al | 0 | 2 | 1 | 48 | 6.33 | 0.20 | 198.32 | 9.88 | | | | |
| 142 - S Richardson et al | 0 | 2 | 0 | 108 | 0.00 | 0.00 | 0.00 | 0.00 | | | | |
| 151 - B Sousa et al | 61 | 273 | 503 | 6663 | 3.52 | 2.61 | 4.75 | 21.34 | | | | |
| 15 - JP Armann et al | 0 | 9 | 1 | 93 | 3.25 | 0.12 | 85.36 | 10.44 | | | | |
| 19 - S Bellino et al | 3 | 17 | 1 | 3819 | 818.14 | 80.14 | 8351.99 | 14.01 | | | | |
| 21 - D Bayesheva et al | 0 | 7 | 0 | 545 | 0.00 | 0.00 | 0.00 | 0.00 | | | | |
| 28 - H Cairoli | 0 | 4 | 2 | 574 | 25.44 | 1.06 | 608.73 | 10.75 | | | | |
| 79 - F Gotzinger et al | 1 | 25 | 3 | 557 | 7.69 | 0.77 | 76.73 | 14.11 | | | | |
| 92 - R Marcello et al | 0 | 3 | 1 | 29 | 2.71 | 0.09 | 80.33 | 10.06 | | | | |
| PIMS-TS/MIS-C | 0 | 2 | 1 | 43 | 6.56 | 0.18 | 239.09 | 9.42 | 1.00 | 0.00 | 1.02 | 0.31 |
| 121 - C Moraleda et al | 0 | 1 | 1 | 30 | 6.56 | 0.18 | 239.09 | 9.42 | | | | |
| 13 - AR Araujo da Silva et al | 0 | 1 | 0 | 13 | 0.00 | 0.00 | 0.00 | 0.00 | | | | |
| Respiratory Co-morbidity | | | | | | | | | | | | |
| Invasive Ventilation | 108 | 997 | 769 | 7923 | 2.15 | 1.02 | 4.53 | 100.00 | 0.00 | 60.18 | 2.01 | 0.04 |
| COVID-19 | 105 | 987 | 738 | 7841 | 2.61 | 1.12 | 6.07 | 83.00 | 0.00 | 68.78 | 2.23 | 0.03 |
| 13 - AR Araujo da Silva et al | 1 | 8 | 3 | 42 | 1.86 | 0.17 | 20.51 | 6.30 | | | | |
| 142 - S Richardson et al | 1 | 10 | 13 | 100 | 0.74 | 0.09 | 6.36 | 7.25 | | | | |
| 151 - B Sousa et al | 70 | 692 | 685 | 6244 | 0.91 | 0.70 | 1.18 | 17.82 | | | | |
| 15 - JP Armann et al | 2 | 11 | 4 | 91 | 4.83 | 0.77 | 30.16 | 8.68 | | | | |
| 174 - P Zachariah et al | 2 | 8 | 7 | 42 | 1.67 | 0.28 | 10.03 | 8.86 | | | | |
| 28 - H Cairoli | 0 | 66 | 1 | 512 | 2.56 | 0.10 | 63.58 | 4.16 | | | | |
| 54 - A Desai et al | 24 | 162 | 4 | 131 | 5.52 | 1.86 | 16.35 | 13.14 | | | | |
| 75 - V Giacomet et al | 0 | 1 | 1 | 126 | 27.89 | 0.78 | 1003.14 | 3.50 | | | | |
| 79 - F Gotzinger et al | 5 | 29 | 20 | 553 | 5.55 | 1.92 | 16.06 | 13.30 | | | | |
| PIMS-TS/MIS-C | 3 | 10 | 31 | 82 | 0.80 | 0.11 | 5.64 | 17.00 | 0.21 | 33.12 | 0.23 | 0.82 |
| 13 - AR Araujo da Silva et al | 0 | 1 | 2 | 13 | 1.53 | 0.05 | 49.80 | 3.66 | | | | |
| 143 - M Riollano-Cruz et al | 0 | 4 | 3 | 11 | 0.27 | 0.01 | 6.46 | 4.23 | | | | |
| 167 - E Whittaker et al | 3 | 4 | 22 | 54 | 4.36 | 0.43 | 44.73 | 6.56 | | | | |
| 179 - J Pang et al | 0 | 1 | 4 | 4 | 0.04 | 0.00 | 2.82 | 2.54 | | | | |

| Name | Events 1 | Total 1 | Events 2 | Total 2 | Effect Estimate | CI Start | CI End | Weight | P(Q) | I²(Q) | Z | P(Z) |
|---|---|---|---|---|---|---|---|---|---|---|---|---|
| Cardiovascular Support | 8 | 110 | 52 | 1220 | 3.52 | 1.42 | 8.75 | 100.00 | 0.99 | 0.00 | 2.71 | 0.01 |
| COVID-19 | 5 | 106 | 23 | 1156 | 3.48 | 1.30 | 9.36 | 84.70 | 0.96 | 0.00 | 2.47 | 0.01 |
| 15 - JP Armann et al | 2 | 11 | 6 | 91 | 3.15 | 0.55 | 17.96 | 27.28 | | | | |
| 28 - H Cairoli | 0 | 66 | 1 | 512 | 2.56 | 0.10 | 63.58 | 8.03 | | | | |
| 79 - F Gotzinger et al | 3 | 29 | 16 | 553 | 3.87 | 1.06 | 14.13 | 49.39 | | | | |
| PIMS-TS/MIS-C | 3 | 4 | 29 | 64 | 3.75 | 0.37 | 38.39 | 15.30 | 1.00 | 0.00 | 1.11 | 0.27 |
| 10 - Z Almoussa et al | 0 | 0 | 5 | 10 | 0.00 | 0.00 | 0.00 | 0.00 | | | | |
| 167 - E Whittaker et al | 3 | 4 | 24 | 54 | 3.75 | 0.37 | 38.39 | 15.30 | | | | |
| Admission to Critical Care | 252 | 939 | 1982 | 8567 | 2.06 | 1.30 | 3.27 | 100.00 | 0.01 | 52.16 | 3.08 | 0.00 |
| COVID-19 | 238 | 924 | 1930 | 8477 | 1.98 | 1.23 | 3.19 | 93.09 | 0.01 | 56.05 | 2.82 | 0.00 |
| 13 - AR Araujo da Silva et al | 7 | 8 | 19 | 42 | 8.47 | 0.96 | 75.08 | 3.53 | | | | |
| 142 - S Richardson et al | 3 | 10 | 34 | 100 | 0.83 | 0.20 | 3.42 | 6.54 | | | | |
| 151 - B Sousa et al | 182 | 692 | 1685 | 6244 | 0.97 | 0.81 | 1.15 | 16.65 | | | | |
| 157 - O Swann et al | 10 | 30 | 68 | 550 | 3.54 | 1.59 | 7.89 | 11.26 | | | | |
| 15 - JP Armann et al | 3 | 11 | 12 | 91 | 2.47 | 0.57 | 10.62 | 6.29 | | | | |
| 163 - S Verma et al | 8 | 16 | 15 | 66 | 3.40 | 1.09 | 10.59 | 8.37 | | | | |
| 224 - Graff et al | 6 | 21 | 5 | 45 | 3.20 | 0.85 | 12.06 | 7.06 | | | | |
| 28 - H Cairoli | 0 | 66 | 3 | 512 | 1.09 | 0.06 | 21.42 | 2.10 | | | | |
| 32 - J Chao et al | 3 | 11 | 10 | 35 | 0.94 | 0.21 | 4.27 | 5.99 | | | | |
| 67 - G Fisler et al | 8 | 16 | 22 | 61 | 1.77 | 0.58 | 5.38 | 8.56 | | | | |
| 75 - V Giacomet et al | 0 | 1 | 8 | 126 | 4.65 | 0.18 | 122.93 | 1.77 | | | | |
| 79 - F Gotzinger et al | 6 | 29 | 42 | 553 | 3.17 | 1.23 | 8.22 | 9.87 | | | | |
| 91 - M Kainth et al | 2 | 13 | 7 | 52 | 1.17 | 0.21 | 6.42 | 5.11 | | | | |
| PIMS-TS/MIS-C | 14 | 15 | 52 | 90 | 3.50 | 0.58 | 21.00 | 6.91 | 0.30 | 16.37 | 1.37 | 0.17 |
| 13 - AR Araujo da Silva et al | 1 | 1 | 11 | 13 | 0.65 | 0.02 | 21.18 | 1.59 | | | | |
| 167 - E Whittaker et al | 3 | 4 | 29 | 54 | 2.59 | 0.25 | 26.46 | 3.19 | | | | |
| 5 - N Abdel-Haq et al | 10 | 10 | 12 | 23 | 19.32 | 1.01 | 368.35 | 2.13 | | | | |
| Death | 41 | 865 | 540 | 8263 | 1.49 | 0.64 | 3.52 | 100.00 | 0.20 | 27.57 | 0.92 | 0.36 |
| COVID-19 | 41 | 856 | 538 | 8185 | 1.70 | 0.59 | 4.90 | 89.44 | 0.10 | 43.07 | 0.98 | 0.33 |
| 13 - AR Araujo da Silva et al | 0 | 8 | 1 | 42 | 1.63 | 0.06 | 43.45 | 5.90 | | | | |

| Name | Events 1 | Total 1 | Events 2 | Total 2 | Effect Estimate | CI Start | CI End | Weight | P(Q) | I²(Q) | Z | P(Z) |
|---|---|---|---|---|---|---|---|---|---|---|---|---|
| 142 - S Richardson et al | 0 | 10 | 0 | 100 | 0.00 | 0.00 | 0.00 | 0.00 | | | | |
| 151 - B Sousa et al | 39 | 692 | 525 | 6244 | 0.65 | 0.47 | 0.91 | 42.16 | | | | |
| 157 - O Swann et al | 2 | 35 | 4 | 616 | 9.27 | 1.64 | 52.47 | 15.82 | | | | |
| 15 - JP Armann et al | 0 | 11 | 1 | 91 | 2.62 | 0.10 | 68.26 | 5.98 | | | | |
| 28 - H Cairoli | 0 | 66 | 2 | 512 | 1.54 | 0.07 | 32.33 | 6.71 | | | | |
| 79 - F Gotzinger et al | 0 | 29 | 4 | 553 | 2.07 | 0.11 | 39.35 | 7.11 | | | | |
| 92 - R Marcello et al | 0 | 5 | 1 | 27 | 1.61 | 0.06 | 44.86 | 5.76 | | | | |
| PIMS-TS/MIS-C | 0 | 9 | 2 | 78 | 1.41 | 0.12 | 16.70 | 10.56 | 0.61 | 0.00 | 0.27 | 0.79 |
| 13 - AR Araujo da Silva et al | 0 | 1 | 1 | 13 | 2.78 | 0.07 | 103.83 | 4.97 | | | | |
| 143 - M Riollano-Cruz et al | 0 | 4 | 1 | 11 | 0.78 | 0.03 | 22.98 | 5.59 | | | | |
| 167 - E Whittaker et al | 0 | 4 | 0 | 54 | 0.00 | 0.00 | 0.00 | 0.00 | | | | |
| Asthma | | | | | | | | | | | | |
| Invasive Ventilation | 52 | 649 | 795 | 8430 | 1.45 | 0.73 | 2.92 | 100.00 | 0.11 | 35.48 | 1.06 | 0.29 |
| COVID-19 | 49 | 641 | 770 | 8365 | 1.46 | 0.69 | 3.08 | 88.76 | 0.11 | 38.59 | 0.99 | 0.32 |
| 13 - AR Araujo da Silva et al | 0 | 7 | 4 | 43 | 0.59 | 0.03 | 12.04 | 4.57 | | | | |
| 142 - S Richardson et al | 1 | 10 | 13 | 100 | 0.74 | 0.09 | 6.36 | 7.98 | | | | |
| 151 - B Sousa et al | 41 | 528 | 714 | 6408 | 0.67 | 0.48 | 0.93 | 30.93 | | | | |
| 155 - Kyung Sung | 0 | 1 | 0 | 100 | 0.00 | 0.00 | 0.00 | 0.00 | | | | |
| 15 - JP Armann et al | 1 | 5 | 5 | 97 | 4.60 | 0.43 | 49.16 | 6.85 | | | | |
| 170 - B Yayla et al | 0 | 1 | 1 | 76 | 16.78 | 0.47 | 605.19 | 3.38 | | | | |
| 174 - P Zachariah et al | 2 | 6 | 7 | 44 | 2.64 | 0.40 | 17.32 | 9.70 | | | | |
| 28 - H Cairoli | 0 | 39 | 1 | 539 | 4.54 | 0.18 | 113.38 | 4.10 | | | | |
| 54 - A Desai et al | 4 | 37 | 24 | 256 | 1.17 | 0.38 | 3.59 | 17.86 | | | | |
| 75 - V Giacomet et al | 0 | 1 | 1 | 126 | 27.89 | 0.78 | 1003.14 | 3.39 | | | | |
| 79 - F Gotzinger et al | 0 | 6 | 0 | 576 | 0.00 | 0.00 | 0.00 | 0.00 | | | | |
| PIMS-TS/MIS-C | 3 | 8 | 25 | 65 | 1.35 | 0.09 | 20.17 | 11.24 | 0.16 | 48.35 | 0.22 | 0.83 |
| 143 - M Riollano-Cruz et al | 0 | 4 | 3 | 11 | 0.27 | 0.01 | 6.46 | 4.19 | | | | |
| 167 - E Whittaker et al | 3 | 4 | 22 | 54 | 4.36 | 0.43 | 44.73 | 7.04 | | | | |
| Cardiovascular Support | 4 | 55 | 51 | 1342 | 3.31 | 0.90 | 12.24 | 100.00 | 0.99 | 0.00 | 1.79 | 0.07 |
| COVID-19 | 1 | 51 | 27 | 1288 | 3.12 | 0.64 | 15.19 | 68.39 | 0.95 | 0.00 | 1.41 | 0.16 |

| Name | Events 1 | Total 1 | Events 2 | Total 2 | Effect Estimate | CI Start | CI End | Weight | P(Q) | I²(Q) | Z | P(Z) |
|---|---|---|---|---|---|---|---|---|---|---|---|---|
| 15 - JP Armann et al | 1 | 5 | 7 | 97 | 3.21 | 0.32 | 32.78 | 31.70 | | | | |
| 170 - B Yayla et al | 0 | 1 | 0 | 76 | 0.00 | 0.00 | 0.00 | 0.00 | | | | |
| 28 - H Cairoli | 0 | 39 | 1 | 539 | 4.54 | 0.18 | 113.38 | 16.52 | | | | |
| 79 - F Gotzinger et al | 0 | 6 | 19 | 576 | 2.20 | 0.12 | 40.44 | 20.17 | | | | |
| PIMS-TS/MIS-C | 3 | 4 | 24 | 54 | 3.75 | 0.37 | 38.39 | 31.61 | 1.00 | 0.00 | 1.11 | 0.27 |
| 167 - E Whittaker et al | 3 | 4 | 24 | 54 | 3.75 | 0.37 | 38.39 | 31.61 | | | | |
| Admission to Critical Care | 160 | 661 | 1955 | 8352 | 1.72 | 0.98 | 3.03 | 100.00 | 0.07 | 39.18 | 1.89 | 0.06 |
| COVID-19 | 147 | 647 | 1914 | 8275 | 1.48 | 0.86 | 2.55 | 92.08 | 0.12 | 33.33 | 1.42 | 0.15 |
| 13 - AR Araujo da Silva et al | 6 | 7 | 20 | 43 | 6.90 | 0.76 | 62.28 | 5.18 | | | | |
| 142 - S Richardson et al | 3 | 10 | 34 | 100 | 0.83 | 0.20 | 3.42 | 9.66 | | | | |
| 151 - B Sousa et al | 123 | 528 | 1744 | 6408 | 0.81 | 0.66 | 1.00 | 23.87 | | | | |
| 155 - Kyung Sung | 0 | 1 | 0 | 100 | 0.00 | 0.00 | 0.00 | 0.00 | | | | |
| 15 - JP Armann et al | 1 | 5 | 14 | 97 | 1.48 | 0.15 | 14.25 | 4.95 | | | | |
| 163 - S Verma et al | 5 | 12 | 18 | 70 | 2.06 | 0.58 | 7.32 | 10.98 | | | | |
| 170 - B Yayla et al | 0 | 1 | 1 | 76 | 16.78 | 0.47 | 605.19 | 2.24 | | | | |
| 224 - Graff et al | 4 | 16 | 7 | 50 | 2.05 | 0.51 | 8.18 | 9.90 | | | | |
| 28 - H Cairoli | 0 | 39 | 3 | 539 | 1.94 | 0.10 | 38.23 | 3.12 | | | | |
| 32 - J Chao et al | 3 | 11 | 10 | 35 | 0.94 | 0.21 | 4.27 | 8.85 | | | | |
| 75 - V Giacomet et al | 0 | 1 | 8 | 126 | 4.65 | 0.18 | 122.93 | 2.64 | | | | |
| 79 - F Gotzinger et al | 2 | 6 | 46 | 576 | 5.76 | 1.03 | 32.30 | 7.45 | | | | |
| 91 - M Kainth et al | 0 | 10 | 9 | 55 | 0.23 | 0.01 | 4.33 | 3.23 | | | | |
| PIMS-TS/MIS-C | 13 | 14 | 41 | 77 | 5.78 | 0.80 | 41.84 | 7.92 | 0.28 | 13.73 | 1.74 | 0.08 |
| 167 - E Whittaker et al | 3 | 4 | 29 | 54 | 2.59 | 0.25 | 26.46 | 4.74 | | | | |
| 5 - N Abdel-Haq et al | 10 | 10 | 12 | 23 | 19.32 | 1.01 | 368.35 | 3.18 | | | | |
| Death | 19 | 614 | 558 | 8058 | 1.83 | 0.62 | 5.41 | 100.00 | 0.09 | 40.19 | 1.09 | 0.28 |
| COVID-19 | 19 | 602 | 555 | 7966 | 2.16 | 0.52 | 8.90 | 76.80 | 0.04 | 53.62 | 1.06 | 0.29 |
| 13 - AR Araujo da Silva et al | 0 | 7 | 1 | 43 | 1.89 | 0.07 | 50.88 | 7.97 | | | | |
| 142 - S Richardson et al | 0 | 10 | 0 | 100 | 0.00 | 0.00 | 0.00 | 0.00 | | | | |
| 151 - B Sousa et al | 19 | 528 | 545 | 6408 | 0.40 | 0.25 | 0.64 | 28.21 | | | | |
| 155 - Kyung Sung | 0 | 1 | 0 | 100 | 0.00 | 0.00 | 0.00 | 0.00 | | | | |

| Name | Events 1 | Total 1 | Events 2 | Total 2 | Effect Estimate | CI Start | CI End | Weight | P(Q) | I²(Q) | Z | P(Z) |
|---|---|---|---|---|---|---|---|---|---|---|---|---|
| 15 - JP Armann et al | 0 | 5 | 1 | 97 | 5.85 | 0.21 | 160.79 | 7.89 | | | | |
| 170 - B Yayla et al | 0 | 1 | 1 | 76 | 16.78 | 0.47 | 605.19 | 7.01 | | | | |
| 28 - H Cairoli | 0 | 39 | 2 | 539 | 2.72 | 0.13 | 57.67 | 8.88 | | | | |
| 79 - F Gotzinger et al | 0 | 6 | 4 | 576 | 9.79 | 0.48 | 201.00 | 9.01 | | | | |
| 92 - R Marcello et al | 0 | 5 | 1 | 27 | 1.61 | 0.06 | 44.86 | 7.84 | | | | |
| PIMS-TS/MIS-C | 0 | 12 | 3 | 92 | 1.84 | 0.26 | 12.77 | 23.20 | 0.79 | 0.00 | 0.61 | 0.54 |
| 121 - C Moraleda et al | 0 | 4 | 1 | 27 | 1.96 | 0.07 | 56.12 | 7.76 | | | | |
| 143 - M Riollano-Cruz et al | 0 | 4 | 1 | 11 | 0.78 | 0.03 | 22.98 | 7.65 | | | | |
| 167 - E Whittaker et al | 0 | 4 | 1 | 54 | 3.96 | 0.14 | 112.06 | 7.80 | | | | |
| Cystic Fibrosis | | | | | | | | | | | | |
| Invasive Ventilation | 0 | 2 | 25 | 580 | 4.36 | 0.20 | 93.13 | 100.00 | 1.00 | 0.00 | 0.94 | 0.35 |
| COVID-19 | 0 | 2 | 25 | 580 | 4.36 | 0.20 | 93.13 | 100.00 | 1.00 | 0.00 | 0.94 | 0.35 |
| 79 - F Gotzinger et al | 0 | 2 | 25 | 580 | 4.36 | 0.20 | 93.13 | 100.00 | | | | |
| PIMS-TS/MIS-C | 0 | 0 | 0 | 0 | 0.00 | 0.00 | 0.00 | 0.00 | 0.00 | 0.00 | 0.00 | 0.00 |
| Cardiovascular Support | 0 | 2 | 27 | 682 | 5.76 | 0.27 | 124.03 | 100.00 | 1.00 | 0.00 | 1.12 | 0.26 |
| COVID-19 | 0 | 2 | 27 | 682 | 5.76 | 0.27 | 124.03 | 100.00 | 1.00 | 0.00 | 1.12 | 0.26 |
| 15 - JP Armann et al | 0 | 0 | 8 | 102 | 0.00 | 0.00 | 0.00 | 0.00 | | | | |
| 79 - F Gotzinger et al | 0 | 2 | 19 | 580 | 5.76 | 0.27 | 124.03 | 100.00 | | | | |
| PIMS-TS/MIS-C | 0 | 0 | 0 | 0 | 0.00 | 0.00 | 0.00 | 0.00 | 0.00 | 0.00 | 0.00 | 0.00 |
| Admission to Critical Care | 0 | 0 | 0 | 0 | 0.00 | 0.00 | 0.00 | 0.00 | 0.00 | 0.00 | 0.00 | 0.00 |
| COVID-19 | 0 | 0 | 0 | 0 | 0.00 | 0.00 | 0.00 | 0.00 | 0.00 | 0.00 | 0.00 | 0.00 |
| PIMS-TS/MIS-C | 0 | 0 | 0 | 0 | 0.00 | 0.00 | 0.00 | 0.00 | 0.00 | 0.00 | 0.00 | 0.00 |
| Death | 0 | 2 | 4 | 580 | 25.62 | 1.07 | 612.98 | 100.00 | 1.00 | 0.00 | 2.00 | 0.05 |
| COVID-19 | 0 | 2 | 4 | 580 | 25.62 | 1.07 | 612.98 | 100.00 | 1.00 | 0.00 | 2.00 | 0.05 |
| 79 - F Gotzinger et al | 0 | 2 | 4 | 580 | 25.62 | 1.07 | 612.98 | 100.00 | | | | |
| PIMS-TS/MIS-C | 0 | 0 | 0 | 0 | 0.00 | 0.00 | 0.00 | 0.00 | 0.00 | 0.00 | 0.00 | 0.00 |
| GI co-morbidity | | | | | | | | | | | | |
| Invasive Ventilation | 8 | 55 | 782 | 8785 | 4.93 | 1.74 | 14.01 | 100.00 | 0.28 | 19.46 | 2.99 | 0.00 |
| COVID-19 | 7 | 54 | 780 | 8771 | 4.86 | 1.51 | 15.67 | 92.02 | 0.24 | 25.90 | 2.64 | 0.01 |
| 13 - AR Araujo da Silva et al | 0 | 1 | 4 | 49 | 3.37 | 0.12 | 95.48 | 8.55 | | | | |

| Name | Events 1 | Total 1 | Events 2 | Total 2 | Effect Estimate | CI Start | CI End | Weight | P(Q) | I²(Q) | Z | P(Z) |
|---|---|---|---|---|---|---|---|---|---|---|---|---|
| 151 - B Sousa et al | 7 | 35 | 748 | 6901 | 2.06 | 0.90 | 4.72 | 48.00 | | | | |
| 21 - D Bayesheva et al | 0 | 1 | 1 | 551 | 122.33 | 3.41 | 4385.74 | 7.58 | | | | |
| 28 - H Cairoli | 0 | 10 | 1 | 568 | 18.02 | 0.69 | 468.51 | 8.94 | | | | |
| 75 - V Giacomet et al | 0 | 4 | 1 | 123 | 9.07 | 0.32 | 255.04 | 8.58 | | | | |
| 79 - F Gotzinger et al | 0 | 3 | 25 | 579 | 3.11 | 0.16 | 61.75 | 10.37 | | | | |
| PIMS-TS/MIS-C | 1 | 1 | 2 | 14 | 15.00 | 0.46 | 485.32 | 7.98 | 1.00 | 0.00 | 1.53 | 0.13 |
| 143 - M Riollano-Cruz et al | 1 | 1 | 2 | 14 | 15.00 | 0.46 | 485.32 | 7.98 | | | | |
| Cardiovascular Support | 0 | 13 | 20 | 1147 | 8.09 | 0.89 | 73.42 | 100.00 | 0.50 | 0.00 | 1.86 | 0.06 |
| COVID-19 | 0 | 13 | 20 | 1147 | 8.09 | 0.89 | 73.42 | 100.00 | 0.50 | 0.00 | 1.86 | 0.06 |
| 28 - H Cairoli | 0 | 10 | 1 | 568 | 18.02 | 0.69 | 468.51 | 45.84 | | | | |
| 79 - F Gotzinger et al | 0 | 3 | 19 | 579 | 4.11 | 0.20 | 82.26 | 54.16 | | | | |
| PIMS-TS/MIS-C | 0 | 0 | 0 | 0 | 0.00 | 0.00 | 0.00 | 0.00 | 0.00 | 0.00 | 0.00 | 0.00 |
| Admission to Critical Care | 29 | 86 | 2022 | 9450 | 3.08 | 1.92 | 4.96 | 100.00 | 0.53 | 0.00 | 4.64 | 0.00 |
| COVID-19 | 29 | 86 | 2022 | 9450 | 3.08 | 1.92 | 4.96 | 100.00 | 0.53 | 0.00 | 4.64 | 0.00 |
| 13 - AR Araujo da Silva et al | 1 | 1 | 25 | 49 | 2.88 | 0.11 | 74.21 | 2.14 | | | | |
| 151 - B Sousa et al | 15 | 35 | 1852 | 6901 | 2.04 | 1.04 | 4.00 | 50.08 | | | | |
| 157 - O Swann et al | 7 | 18 | 71 | 562 | 4.40 | 1.65 | 11.72 | 23.53 | | | | |
| 21 - D Bayesheva et al | 0 | 1 | 4 | 551 | 40.56 | 1.45 | 1135.72 | 2.03 | | | | |
| 224 - Graff et al | 4 | 13 | 7 | 53 | 2.92 | 0.71 | 12.10 | 11.18 | | | | |
| 28 - H Cairoli | 0 | 10 | 3 | 568 | 7.69 | 0.37 | 158.51 | 2.47 | | | | |
| 75 - V Giacomet et al | 1 | 4 | 7 | 123 | 5.52 | 0.51 | 60.18 | 3.96 | | | | |
| 79 - F Gotzinger et al | 0 | 3 | 48 | 579 | 1.57 | 0.08 | 30.75 | 2.55 | | | | |
| 9 - M Alharbi et al | 1 | 1 | 5 | 64 | 32.45 | 1.18 | 895.61 | 2.05 | | | | |
| PIMS-TS/MIS-C | 0 | 0 | 0 | 0 | 0.00 | 0.00 | 0.00 | 0.00 | 0.00 | 0.00 | 0.00 | 0.00 |
| Death | 7 | 51 | 565 | 8662 | 3.63 | 1.72 | 7.65 | 100.00 | 0.69 | 0.00 | 3.40 | 0.00 |
| COVID-19 | 7 | 50 | 564 | 8648 | 3.66 | 1.71 | 7.84 | 95.77 | 0.52 | 0.00 | 3.34 | 0.00 |
| 13 - AR Araujo da Silva et al | 0 | 1 | 1 | 49 | 10.78 | 0.30 | 390.30 | 4.30 | | | | |
| 151 - B Sousa et al | 7 | 35 | 557 | 6901 | 2.85 | 1.24 | 6.55 | 79.93 | | | | |
| 21 - D Bayesheva et al | 0 | 1 | 0 | 551 | 0.00 | 0.00 | 0.00 | 0.00 | | | | |
| 28 - H Cairoli | 0 | 10 | 2 | 568 | 10.79 | 0.49 | 238.84 | 5.78 | | | | |

| Name | Events 1 | Total 1 | Events 2 | Total 2 | Effect Estimate | CI Start | CI End | Weight | P(Q) | I²(Q) | Z | P(Z) |
|---|---|---|---|---|---|---|---|---|---|---|---|---|
| 79 - F Gotzinger et al | 0 | 3 | 4 | 579 | 18.27 | 0.82 | 407.57 | 5.75 | | | | |
| PIMS-TS/MIS-C | 0 | 1 | 1 | 14 | 3.00 | 0.08 | 111.78 | 4.23 | 1.00 | 0.00 | 0.60 | 0.55 |
| 143 - M Riollano-Cruz et al | 0 | 1 | 1 | 14 | 3.00 | 0.08 | 111.78 | 4.23 | | | | |
| Neurological/Neuromuscular Co-morbidity | | | | | | | | | | | | |
| Invasive Ventilation | 145 | 562 | 749 | 9250 | 4.30 | 2.75 | 6.73 | 100.00 | 0.28 | 14.82 | 6.38 | 0.00 |
| COVID-19 | 141 | 553 | 694 | 9109 | 4.24 | 3.41 | 5.27 | 91.31 | 0.45 | 0.00 | 13.00 | 0.00 |
| 12 - OY Antunez-Montes et al | 0 | 9 | 3 | 172 | 2.55 | 0.12 | 52.99 | 2.08 | | | | |
| 13 - AR Araujo da Silva et al | 2 | 14 | 2 | 36 | 2.83 | 0.36 | 22.40 | 4.27 | | | | |
| 151 - B Sousa et al | 112 | 372 | 643 | 6564 | 3.97 | 3.13 | 5.02 | 41.54 | | | | |
| 155 - Kyung Sung | 0 | 1 | 0 | 100 | 0.00 | 0.00 | 0.00 | 0.00 | | | | |
| 15 - JP Armann et al | 2 | 6 | 4 | 96 | 11.50 | 1.60 | 82.51 | 4.65 | | | | |
| 170 - B Yayla et al | 0 | 2 | 1 | 75 | 9.93 | 0.32 | 309.72 | 1.64 | | | | |
| 174 - P Zachariah et al | 1 | 7 | 8 | 43 | 0.73 | 0.08 | 6.93 | 3.65 | | | | |
| 21 - D Bayesheva et al | 0 | 9 | 1 | 543 | 19.04 | 0.73 | 497.97 | 1.81 | | | | |
| 28 - H Cairoli | 0 | 26 | 1 | 552 | 6.94 | 0.28 | 174.36 | 1.85 | | | | |
| 46 - M Ceano-Vivas et al | 1 | 2 | 0 | 31 | 63.00 | 1.73 | 2295.08 | 1.50 | | | | |
| 54 - A Desai et al | 20 | 77 | 8 | 216 | 9.12 | 3.82 | 21.79 | 16.92 | | | | |
| 75 - V Giacomet et al | 0 | 2 | 1 | 125 | 16.60 | 0.53 | 516.03 | 1.64 | | | | |
| 79 - F Gotzinger et al | 3 | 26 | 22 | 556 | 3.17 | 0.88 | 11.35 | 9.75 | | | | |
| PIMS-TS/MIS-C | 4 | 9 | 55 | 141 | 0.99 | 0.22 | 4.48 | 8.69 | 0.39 | 2.80 | 0.02 | 0.99 |
| 12 - OY Antunez-Montes et al | 2 | 2 | 22 | 65 | 9.67 | 0.44 | 210.06 | 2.03 | | | | |
| 13 - AR Araujo da Silva et al | 0 | 2 | 2 | 12 | 0.84 | 0.03 | 23.58 | 1.74 | | | | |
| 167 - E Whittaker et al | 0 | 2 | 25 | 56 | 0.25 | 0.01 | 5.38 | 2.02 | | | | |
| 179 - J Pang et al | 1 | 2 | 3 | 3 | 0.14 | 0.00 | 5.95 | 1.40 | | | | |
| 9 - M Alharbi et al | 1 | 1 | 3 | 5 | 2.14 | 0.06 | 77.54 | 1.51 | | | | |
| Cardiovascular Support | 7 | 81 | 81 | 1668 | 3.61 | 1.35 | 9.64 | 100.00 | 0.58 | 0.00 | 2.56 | 0.01 |
| COVID-19 | 3 | 76 | 30 | 1542 | 4.31 | 1.32 | 14.13 | 70.62 | 0.40 | 2.63 | 2.41 | 0.02 |
| 12 - OY Antunez-Montes et al | 0 | 9 | 1 | 172 | 6.02 | 0.23 | 157.79 | 9.05 | | | | |
| 15 - JP Armann et al | 1 | 6 | 7 | 96 | 2.54 | 0.26 | 24.88 | 18.56 | | | | |
| 170 - B Yayla et al | 0 | 2 | 0 | 75 | 0.00 | 0.00 | 0.00 | 0.00 | | | | |

| Name | Events 1 | Total 1 | Events 2 | Total 2 | Effect Estimate | CI Start | CI End | Weight | P(Q) | I²(Q) | Z | P(Z) |
|---|---|---|---|---|---|---|---|---|---|---|---|---|
| 28 - H Cairoli | 0 | 26 | 1 | 552 | 6.94 | 0.28 | 174.36 | 9.29 | | | | |
| 46 - M Ceano-Vivas et al | 1 | 2 | 0 | 31 | 63.00 | 1.73 | 2295.08 | 7.47 | | | | |
| 79 - F Gotzinger et al | 0 | 26 | 19 | 556 | 0.52 | 0.03 | 8.85 | 12.02 | | | | |
| 9 - M Alharbi et al | 1 | 5 | 2 | 60 | 7.25 | 0.54 | 98.15 | 14.22 | | | | |
| PIMS-TS/MIS-C | 4 | 5 | 51 | 126 | 2.38 | 0.39 | 14.61 | 29.38 | 0.51 | 0.00 | 0.94 | 0.35 |
| 12 - OY Antunez-Montes et al | 2 | 2 | 21 | 65 | 10.35 | 0.48 | 225.10 | 10.18 | | | | |
| 167 - E Whittaker et al | 1 | 2 | 26 | 56 | 1.15 | 0.07 | 19.38 | 12.13 | | | | |
| 9 - M Alharbi et al | 1 | 1 | 4 | 5 | 1.00 | 0.02 | 40.28 | 7.07 | | | | |
| Admission to Critical Care | 223 | 563 | 1967 | 9850 | 2.61 | 2.17 | 3.14 | 100.00 | 0.92 | 0.00 | 10.14 | 0.00 |
| COVID-19 | 216 | 556 | 1896 | 9712 | 2.60 | 2.16 | 3.13 | 98.97 | 0.84 | 0.00 | 10.05 | 0.00 |
| 12 - OY Antunez-Montes et al | 0 | 9 | 11 | 172 | 0.74 | 0.04 | 13.51 | 0.41 | | | | |
| 136 - G Qian et al | 0 | 1 | 7 | 126 | 5.31 | 0.20 | 141.79 | 0.32 | | | | |
| 13 - AR Araujo da Silva et al | 8 | 14 | 18 | 36 | 1.33 | 0.38 | 4.63 | 2.22 | | | | |
| 151 - B Sousa et al | 171 | 372 | 1696 | 6564 | 2.44 | 1.98 | 3.02 | 76.96 | | | | |
| 155 - Kyung Sung | 0 | 1 | 0 | 100 | 0.00 | 0.00 | 0.00 | 0.00 | | | | |
| 157 - O Swann et al | 17 | 63 | 61 | 517 | 2.76 | 1.49 | 5.12 | 9.02 | | | | |
| 15 - JP Armann et al | 3 | 6 | 12 | 96 | 7.00 | 1.27 | 38.73 | 1.17 | | | | |
| 170 - B Yayla et al | 0 | 2 | 1 | 75 | 9.93 | 0.32 | 309.72 | 0.29 | | | | |
| 21 - D Bayesheva et al | 0 | 9 | 4 | 543 | 6.31 | 0.32 | 125.64 | 0.38 | | | | |
| 224 - Graff et al | 3 | 8 | 8 | 58 | 3.75 | 0.75 | 18.84 | 1.32 | | | | |
| 26 - CE Bolaños-Almeida et al | 0 | 26 | 3 | 552 | 2.96 | 0.15 | 58.83 | 0.38 | | | | |
| 32 - J Chao et al | 3 | 4 | 10 | 42 | 9.60 | 0.90 | 102.90 | 0.61 | | | | |
| 46 - M Ceano-Vivas et al | 1 | 2 | 4 | 31 | 6.75 | 0.35 | 130.79 | 0.39 | | | | |
| 75 - V Giacomet et al | 1 | 2 | 7 | 125 | 16.86 | 0.95 | 298.73 | 0.42 | | | | |
| 79 - F Gotzinger et al | 5 | 26 | 43 | 556 | 2.84 | 1.02 | 7.91 | 3.28 | | | | |
| 91 - M Kainth et al | 2 | 6 | 7 | 59 | 3.71 | 0.57 | 24.14 | 0.98 | | | | |
| 9 - M Alharbi et al | 2 | 5 | 4 | 60 | 9.33 | 1.19 | 72.99 | 0.81 | | | | |
| PIMS-TS/MIS-C | 7 | 7 | 71 | 138 | 3.57 | 0.58 | 22.08 | 1.03 | 0.72 | 0.00 | 1.37 | 0.17 |
| 12 - OY Antunez-Montes et al | 2 | 2 | 26 | 65 | 7.45 | 0.34 | 161.51 | 0.36 | | | | |
| 13 - AR Araujo da Silva et al | 2 | 2 | 10 | 12 | 1.19 | 0.04 | 33.43 | 0.31 | | | | |

| Name | Events 1 | Total 1 | Events 2 | Total 2 | Effect Estimate | CI Start | CI End | Weight | P(Q) | I²(Q) | Z | P(Z) |
|---|---|---|---|---|---|---|---|---|---|---|---|---|
| 167 - E Whittaker et al | 2 | 2 | 30 | 56 | 4.34 | 0.20 | 94.59 | 0.36 | | | | |
| 9 - M Alharbi et al | 1 | 1 | 5 | 5 | 0.00 | 0.00 | 0.00 | 0.00 | | | | |
| Death | 63 | 531 | 524 | 9403 | 2.47 | 1.81 | 3.37 | 100.00 | 0.44 | 0.48 | 5.72 | 0.00 |
| COVID-19 | 62 | 524 | 518 | 9265 | 4.34 | 2.00 | 9.43 | 97.55 | 0.24 | 23.35 | 3.71 | 0.00 |
| 136 - G Qian et al | 0 | 1 | 0 | 126 | 0.00 | 0.00 | 0.00 | 0.00 | | | | |
| 13 - AR Araujo da Silva et al | 1 | 14 | 0 | 36 | 8.11 | 0.31 | 211.49 | 0.90 | | | | |
| 151 - B Sousa et al | 56 | 372 | 508 | 6564 | 2.11 | 1.57 | 2.85 | 87.95 | | | | |
| 155 - Kyung Sung | 0 | 1 | 0 | 100 | 0.00 | 0.00 | 0.00 | 0.00 | | | | |
| 157 - O Swann et al | 2 | 65 | 4 | 586 | 4.62 | 0.83 | 25.72 | 3.23 | | | | |
| 15 - JP Armann et al | 0 | 6 | 1 | 96 | 4.90 | 0.18 | 132.48 | 0.88 | | | | |
| 170 - B Yayla et al | 0 | 2 | 1 | 75 | 9.93 | 0.32 | 309.72 | 0.81 | | | | |
| 21 - D Bayesheva et al | 0 | 9 | 0 | 543 | 0.00 | 0.00 | 0.00 | 0.00 | | | | |
| 28 - H Cairoli | 1 | 26 | 1 | 552 | 22.04 | 1.34 | 362.67 | 1.22 | | | | |
| 46 - M Ceano-Vivas et al | 1 | 2 | 0 | 31 | 63.00 | 1.73 | 2295.08 | 0.74 | | | | |
| 79 - F Gotzinger et al | 1 | 26 | 3 | 556 | 7.37 | 0.74 | 73.42 | 1.81 | | | | |
| PIMS-TS/MIS-C | 1 | 7 | 6 | 138 | 5.33 | 0.74 | 38.45 | 2.45 | 0.87 | 0.00 | 1.66 | 0.10 |
| 12 - OY Antunez-Montes et al | 0 | 2 | 4 | 65 | 2.73 | 0.11 | 65.97 | 0.94 | | | | |
| 13 - AR Araujo da Silva et al | 0 | 2 | 0 | 12 | 0.00 | 0.00 | 0.00 | 0.00 | | | | |
| 167 - E Whittaker et al | 0 | 2 | 1 | 56 | 7.40 | 0.24 | 231.32 | 0.81 | | | | |
| 9 - M Alharbi et al | 1 | 1 | 1 | 5 | 9.00 | 0.22 | 362.48 | 0.70 | | | | |
| Oncological co-morbidity | | | | | | | | | | | | |
| Invasive Ventilation | 9 | 86 | 58 | 1801 | 2.73 | 1.35 | 5.54 | 100.00 | 0.82 | 0.00 | 2.79 | 0.01 |
| COVID-19 | 9 | 86 | 58 | 1801 | 2.73 | 1.35 | 5.54 | 100.00 | 0.82 | 0.00 | 2.79 | 0.01 |
| 12 - OY Antunez-Montes et al | 0 | 7 | 3 | 174 | 3.27 | 0.15 | 69.12 | 5.35 | | | | |
| 13 - AR Araujo da Silva et al | 0 | 5 | 4 | 45 | 0.84 | 0.04 | 17.78 | 5.34 | | | | |
| 155 - Kyung Sung | 0 | 3 | 0 | 98 | 0.00 | 0.00 | 0.00 | 0.00 | | | | |
| 15 - JP Armann et al | 1 | 7 | 5 | 95 | 3.00 | 0.30 | 29.94 | 9.41 | | | | |
| 28 - H Cairoli | 0 | 9 | 1 | 569 | 19.95 | 0.76 | 521.81 | 4.67 | | | | |
| 54 - A Desai et al | 5 | 28 | 23 | 265 | 2.29 | 0.79 | 6.59 | 44.54 | | | | |
| 79 - F Gotzinger et al | 3 | 27 | 22 | 555 | 3.03 | 0.85 | 10.82 | 30.70 | | | | |

| Name | Events 1 | Total 1 | Events 2 | Total 2 | Effect Estimate | CI Start | CI End | Weight | P(Q) | I²(Q) | Z | P(Z) |
|---|---|---|---|---|---|---|---|---|---|---|---|---|
| PIMS-TS/MIS-C | 0 | 0 | 0 | 0 | 0.00 | 0.00 | 0.00 | 0.00 | 0.00 | 0.00 | 0.00 | 0.00 |
| Cardiovascular Support | 3 | 50 | 26 | 1393 | 3.48 | 1.15 | 10.52 | 100.00 | 0.62 | 0.00 | 2.21 | 0.03 |
| COVID-19 | 3 | 50 | 26 | 1393 | 3.48 | 1.15 | 10.52 | 100.00 | 0.62 | 0.00 | 2.21 | 0.03 |
| 12 - OY Antunez-Montes et al | 0 | 7 | 1 | 174 | 7.71 | 0.29 | 205.55 | 11.36 | | | | |
| 15 - JP Armann et al | 1 | 7 | 7 | 95 | 2.10 | 0.22 | 19.93 | 24.12 | | | | |
| 28 - H Cairoli | 0 | 9 | 1 | 569 | 19.95 | 0.76 | 521.81 | 11.49 | | | | |
| 79 - F Gotzinger et al | 2 | 27 | 17 | 555 | 2.53 | 0.55 | 11.56 | 53.04 | | | | |
| PIMS-TS/MIS-C | 0 | 0 | 0 | 0 | 0.00 | 0.00 | 0.00 | 0.00 | 0.00 | 0.00 | 0.00 | 0.00 |
| Admission to Critical Care | 10 | 72 | 152 | 1837 | 1.77 | 0.51 | 6.11 | 100.00 | 0.01 | 58.59 | 0.91 | 0.36 |
| COVID-19 | 10 | 72 | 152 | 1837 | 1.77 | 0.51 | 6.11 | 100.00 | 0.01 | 58.59 | 0.91 | 0.36 |
| 12 - OY Antunez-Montes et al | 0 | 7 | 11 | 174 | 0.95 | 0.05 | 17.65 | 9.59 | | | | |
| 136 - G Qian et al | 2 | 3 | 5 | 124 | 47.60 | 3.67 | 616.85 | 10.96 | | | | |
| 13 - AR Araujo da Silva et al | 0 | 5 | 26 | 45 | 0.07 | 0.00 | 1.28 | 9.49 | | | | |
| 155 - Kyung Sung | 0 | 3 | 0 | 98 | 0.00 | 0.00 | 0.00 | 0.00 | | | | |
| 15 - JP Armann et al | 1 | 7 | 14 | 95 | 0.96 | 0.11 | 8.63 | 12.54 | | | | |
| 28 - H Cairoli | 0 | 9 | 3 | 569 | 8.52 | 0.41 | 176.63 | 9.22 | | | | |
| 32 - J Chao et al | 1 | 1 | 12 | 45 | 8.04 | 0.31 | 210.67 | 8.47 | | | | |
| 67 - G Fisler et al | 0 | 5 | 30 | 72 | 0.13 | 0.01 | 2.38 | 9.57 | | | | |
| 79 - F Gotzinger et al | 5 | 27 | 43 | 555 | 2.71 | 0.98 | 7.50 | 18.14 | | | | |
| 91 - M Kainth et al | 1 | 5 | 8 | 60 | 1.63 | 0.16 | 16.44 | 12.00 | | | | |
| PIMS-TS/MIS-C | 0 | 0 | 0 | 0 | 0.00 | 0.00 | 0.00 | 0.00 | 0.00 | 0.00 | 0.00 | 0.00 |
| Death | 5 | 119 | 16 | 6025 | 12.83 | 4.02 | 40.99 | 100.00 | 0.18 | 30.42 | 4.31 | 0.00 |
| COVID-19 | 4 | 118 | 16 | 5995 | 10.76 | 3.27 | 35.41 | 93.77 | 0.18 | 30.99 | 3.91 | 0.00 |
| 12 - OY Antunez-Montes et al | 0 | 7 | 1 | 174 | 7.71 | 0.29 | 205.55 | 9.38 | | | | |
| 13 - AR Araujo da Silva et al | 0 | 5 | 1 | 45 | 2.70 | 0.10 | 74.67 | 9.22 | | | | |
| 155 - Kyung Sung | 0 | 3 | 0 | 98 | 0.00 | 0.00 | 0.00 | 0.00 | | | | |
| 157 - O Swann et al | 1 | 48 | 5 | 603 | 2.54 | 0.29 | 22.23 | 16.24 | | | | |
| 15 - JP Armann et al | 0 | 7 | 1 | 95 | 4.20 | 0.16 | 112.27 | 9.36 | | | | |
| 19 - S Bellino et al | 1 | 11 | 3 | 3825 | 127.40 | 12.19 | 1331.49 | 14.80 | | | | |
| 28 - H Cairoli | 1 | 9 | 1 | 569 | 71.00 | 4.07 | 1237.66 | 11.45 | | | | |

| Name | Events 1 | Total 1 | Events 2 | Total 2 | Effect Estimate | CI Start | CI End | Weight | P(Q) | I²(Q) | Z | P(Z) |
|---|---|---|---|---|---|---|---|---|---|---|---|---|
| 79 - F Gotzinger et al | 1 | 27 | 3 | 555 | 7.08 | 0.71 | 70.38 | 15.18 | | | | |
| 92 - R Marcello et al | 0 | 1 | 1 | 31 | 6.78 | 0.19 | 247.05 | 8.16 | | | | |
| PIMS-TS/MIS-C | 1 | 1 | 0 | 30 | 183.00 | 2.61 | 12814.59 | 6.23 | 1.00 | 0.00 | 2.40 | 0.02 |
| 121 - C Moraleda et al | 1 | 1 | 0 | 30 | 183.00 | 2.61 | 12814.59 | 6.23 | | | | |
| Non-malig haematological disease | | | | | | | | | | | | |
| Invasive Ventilation | 25 | 232 | 773 | 8695 | 2.11 | 0.99 | 4.52 | 100.00 | 0.34 | 11.29 | 1.93 | 0.05 |
| COVID-19 | 25 | 232 | 773 | 8695 | 2.11 | 0.99 | 4.52 | 100.00 | 0.34 | 11.29 | 1.93 | 0.05 |
| 151 - B Sousa et al | 24 | 178 | 731 | 6758 | 1.28 | 0.83 | 1.99 | 67.16 | | | | |
| 15 - JP Armann et al | 0 | 2 | 6 | 100 | 2.91 | 0.13 | 67.09 | 5.49 | | | | |
| 174 - P Zachariah et al | 1 | 2 | 8 | 48 | 5.00 | 0.28 | 88.53 | 6.47 | | | | |
| 21 - D Bayesheva et al | 0 | 34 | 1 | 518 | 5.00 | 0.20 | 125.03 | 5.23 | | | | |
| 28 - H Cairoli | 0 | 10 | 1 | 568 | 18.02 | 0.69 | 468.51 | 5.11 | | | | |
| 75 - V Giacomet et al | 0 | 1 | 1 | 126 | 27.89 | 0.78 | 1003.14 | 4.27 | | | | |
| 79 - F Gotzinger et al | 0 | 5 | 25 | 577 | 1.97 | 0.11 | 36.60 | 6.27 | | | | |
| PIMS-TS/MIS-C | 0 | 0 | 0 | 0 | 0.00 | 0.00 | 0.00 | 0.00 | 0.00 | 0.00 | 0.00 | 0.00 |
| Cardiovascular Support | 1 | 17 | 27 | 1245 | 6.82 | 1.39 | 33.37 | 100.00 | 0.62 | 0.00 | 2.37 | 0.02 |
| COVID-19 | 1 | 17 | 27 | 1245 | 6.82 | 1.39 | 33.37 | 100.00 | 0.62 | 0.00 | 2.37 | 0.02 |
| 15 - JP Armann et al | 0 | 2 | 8 | 100 | 2.18 | 0.10 | 49.12 | 25.98 | | | | |
| 28 - H Cairoli | 0 | 10 | 1 | 568 | 18.02 | 0.69 | 468.51 | 23.77 | | | | |
| 79 - F Gotzinger et al | 1 | 5 | 18 | 577 | 7.76 | 0.83 | 73.00 | 50.25 | | | | |
| PIMS-TS/MIS-C | 0 | 0 | 0 | 0 | 0.00 | 0.00 | 0.00 | 0.00 | 0.00 | 0.00 | 0.00 | 0.00 |
| Admission to Critical Care | 49 | 234 | 1905 | 8708 | 1.07 | 0.77 | 1.47 | 100.00 | 0.74 | 0.00 | 0.39 | 0.70 |
| COVID-19 | 49 | 234 | 1905 | 8708 | 1.07 | 0.77 | 1.47 | 100.00 | 0.74 | 0.00 | 0.39 | 0.70 |
| 151 - B Sousa et al | 48 | 178 | 1819 | 6758 | 1.00 | 0.72 | 1.40 | 92.33 | | | | |
| 15 - JP Armann et al | 0 | 2 | 15 | 100 | 1.10 | 0.05 | 24.11 | 1.09 | | | | |
| 21 - D Bayesheva et al | 0 | 34 | 4 | 518 | 1.66 | 0.09 | 31.41 | 1.20 | | | | |
| 28 - H Cairoli | 0 | 10 | 3 | 568 | 7.69 | 0.37 | 158.51 | 1.13 | | | | |
| 75 - V Giacomet et al | 0 | 1 | 8 | 126 | 4.65 | 0.18 | 122.93 | 0.97 | | | | |
| 79 - F Gotzinger et al | 1 | 5 | 47 | 577 | 2.82 | 0.31 | 25.74 | 2.12 | | | | |
| 91 - M Kainth et al | 0 | 4 | 9 | 61 | 0.61 | 0.03 | 12.36 | 1.15 | | | | |

| Name | Events 1 | Total 1 | Events 2 | Total 2 | Effect Estimate | CI Start | CI End | Weight | P(Q) | I²(Q) | Z | P(Z) |
|---|---|---|---|---|---|---|---|---|---|---|---|---|
| PIMS-TS/MIS-C | 0 | 0 | 0 | 0 | 0.00 | 0.00 | 0.00 | 0.00 | 0.00 | 0.00 | 0.00 | 0.00 |
| Death | 26 | 230 | 546 | 8551 | 2.20 | 1.46 | 3.32 | 100.00 | 0.43 | 0.00 | 3.75 | 0.00 |
| COVID-19 | 26 | 229 | 545 | 8521 | 2.92 | 1.20 | 7.13 | 98.69 | 0.32 | 13.84 | 2.35 | 0.02 |
| 151 - B Sousa et al | 26 | 178 | 538 | 6758 | 1.98 | 1.29 | 3.03 | 93.66 | | | | |
| 15 - JP Armann et al | 0 | 2 | 1 | 100 | 13.27 | 0.43 | 412.87 | 1.43 | | | | |
| 21 - D Bayesheva et al | 0 | 34 | 0 | 518 | 0.00 | 0.00 | 0.00 | 0.00 | | | | |
| 28 - H Cairoli | 0 | 10 | 2 | 568 | 10.79 | 0.49 | 238.84 | 1.77 | | | | |
| 79 - F Gotzinger et al | 0 | 5 | 4 | 577 | 11.59 | 0.55 | 242.21 | 1.83 | | | | |
| PIMS-TS/MIS-C | 0 | 1 | 1 | 30 | 6.56 | 0.18 | 239.09 | 1.31 | 1.00 | 0.00 | 1.02 | 0.31 |
| 121 - C Moraleda et al | 0 | 1 | 1 | 30 | 6.56 | 0.18 | 239.09 | 1.31 | | | | |
| Transplant (BM or solid organ) | | | | | | | | | | | | |
| Invasive Ventilation | 2 | 3 | 12 | 176 | 17.08 | 1.62 | 180.17 | 100.00 | 0.30 | 6.76 | 2.36 | 0.02 |
| COVID-19 | 2 | 3 | 12 | 176 | 17.08 | 1.62 | 180.17 | 100.00 | 0.30 | 6.76 | 2.36 | 0.02 |
| 15 - JP Armann et al | 0 | 1 | 6 | 101 | 4.90 | 0.18 | 132.48 | 47.73 | | | | |
| 67 - G Fisler et al | 2 | 2 | 6 | 75 | 53.46 | 2.31 | 1236.38 | 52.27 | | | | |
| PIMS-TS/MIS-C | 0 | 0 | 0 | 0 | 0.00 | 0.00 | 0.00 | 0.00 | 0.00 | 0.00 | 0.00 | 0.00 |
| Cardiovascular Support | 0 | 1 | 8 | 101 | 3.67 | 0.14 | 97.13 | 100.00 | 1.00 | 0.00 | 0.78 | 0.44 |
| COVID-19 | 0 | 1 | 8 | 101 | 3.67 | 0.14 | 97.13 | 100.00 | 1.00 | 0.00 | 0.78 | 0.44 |
| 15 - JP Armann et al | 0 | 1 | 8 | 101 | 3.67 | 0.14 | 97.13 | 100.00 | | | | |
| PIMS-TS/MIS-C | 0 | 0 | 0 | 0 | 0.00 | 0.00 | 0.00 | 0.00 | 0.00 | 0.00 | 0.00 | 0.00 |
| Admission to Critical Care | 1 | 3 | 44 | 176 | 1.70 | 0.20 | 14.22 | 100.00 | 0.94 | 0.00 | 0.49 | 0.63 |
| COVID-19 | 1 | 3 | 44 | 176 | 1.70 | 0.20 | 14.22 | 100.00 | 0.94 | 0.00 | 0.49 | 0.63 |
| 15 - JP Armann et al | 0 | 1 | 15 | 101 | 1.86 | 0.07 | 47.78 | 42.85 | | | | |
| 67 - G Fisler et al | 1 | 2 | 29 | 75 | 1.59 | 0.10 | 26.36 | 57.15 | | | | |
| PIMS-TS/MIS-C | 0 | 0 | 0 | 0 | 0.00 | 0.00 | 0.00 | 0.00 | 0.00 | 0.00 | 0.00 | 0.00 |
| Death | 0 | 1 | 1 | 101 | 22.33 | 0.62 | 804.17 | 100.00 | 1.00 | 0.00 | 1.70 | 0.09 |
| COVID-19 | 0 | 1 | 1 | 101 | 22.33 | 0.62 | 804.17 | 100.00 | 1.00 | 0.00 | 1.70 | 0.09 |
| 15 - JP Armann et al | 0 | 1 | 1 | 101 | 22.33 | 0.62 | 804.17 | 100.00 | | | | |
| PIMS-TS/MIS-C | 0 | 0 | 0 | 0 | 0.00 | 0.00 | 0.00 | 0.00 | 0.00 | 0.00 | 0.00 | 0.00 |
| Immunosuppression (other) | | | | | | | | | | | | |

| Name | Events 1 | Total 1 | Events 2 | Total 2 | Effect Estimate | CI Start | CI End | Weight | P(Q) | I²(Q) | Z | P(Z) |
|---|---|---|---|---|---|---|---|---|---|---|---|---|
| Invasive Ventilation | 67 | 417 | 757 | 7628 | 3.01 | 1.04 | 8.70 | 100.00 | 0.05 | 51.77 | 2.04 | 0.04 |
| COVID-19 | 66 | 416 | 734 | 7562 | 2.92 | 0.91 | 9.38 | 91.91 | 0.04 | 58.24 | 1.80 | 0.07 |
| 12 - OY Antunez-Montes et al | 0 | 2 | 3 | 179 | 10.09 | 0.40 | 251.27 | 8.18 | | | | |
| 151 - B Sousa et al | 61 | 369 | 694 | 6567 | 1.68 | 1.26 | 2.23 | 32.12 | | | | |
| 15 - JP Armann et al | 3 | 6 | 3 | 96 | 31.00 | 4.32 | 222.40 | 15.41 | | | | |
| 174 - P Zachariah et al | 1 | 8 | 8 | 42 | 0.61 | 0.07 | 5.66 | 13.39 | | | | |
| 75 - V Giacomet et al | 0 | 2 | 2 | 125 | 9.88 | 0.37 | 263.72 | 7.92 | | | | |
| 79 - F Gotzinger et al | 1 | 29 | 24 | 553 | 0.79 | 0.10 | 6.03 | 14.88 | | | | |
| PIMS-TS/MIS-C | 1 | 1 | 23 | 66 | 5.55 | 0.22 | 141.75 | 8.09 | 1.00 | 0.00 | 1.04 | 0.30 |
| 12 - OY Antunez-Montes et al | 1 | 1 | 23 | 66 | 5.55 | 0.22 | 141.75 | 8.09 | | | | |
| Cardiovascular Support | 2 | 38 | 49 | 894 | 2.95 | 0.61 | 14.27 | 100.00 | 0.31 | 17.06 | 1.35 | 0.18 |
| COVID-19 | 1 | 37 | 27 | 828 | 2.59 | 0.30 | 22.27 | 79.69 | 0.17 | 43.25 | 0.87 | 0.38 |
| 12 - OY Antunez-Montes et al | 0 | 2 | 1 | 179 | 23.80 | 0.77 | 738.85 | 18.35 | | | | |
| 15 - JP Armann et al | 1 | 6 | 7 | 96 | 2.54 | 0.26 | 24.88 | 35.85 | | | | |
| 79 - F Gotzinger et al | 0 | 29 | 19 | 553 | 0.46 | 0.03 | 7.88 | 25.49 | | | | |
| PIMS-TS/MIS-C | 1 | 1 | 22 | 66 | 5.93 | 0.23 | 151.58 | 20.31 | 1.00 | 0.00 | 1.08 | 0.28 |
| 12 - OY Antunez-Montes et al | 1 | 1 | 22 | 66 | 5.93 | 0.23 | 151.58 | 20.31 | | | | |
| Admission to Critical Care | 136 | 492 | 1982 | 8337 | 1.29 | 0.88 | 1.90 | 100.00 | 0.32 | 12.54 | 1.30 | 0.19 |
| COVID-19 | 135 | 491 | 1955 | 8271 | 1.27 | 0.84 | 1.94 | 98.61 | 0.28 | 17.31 | 1.13 | 0.26 |
| 12 - OY Antunez-Montes et al | 0 | 2 | 11 | 179 | 2.93 | 0.13 | 64.70 | 1.52 | | | | |
| 151 - B Sousa et al | 116 | 369 | 1751 | 6567 | 1.26 | 1.01 | 1.58 | 54.16 | | | | |
| 157 - O Swann et al | 6 | 48 | 72 | 532 | 0.91 | 0.37 | 2.22 | 14.62 | | | | |
| 15 - JP Armann et al | 4 | 6 | 11 | 96 | 15.45 | 2.53 | 94.42 | 4.25 | | | | |
| 224 - Graff et al | 1 | 12 | 10 | 54 | 0.40 | 0.05 | 3.47 | 3.04 | | | | |
| 32 - J Chao et al | 1 | 2 | 12 | 44 | 2.67 | 0.15 | 46.11 | 1.78 | | | | |
| 67 - G Fisler et al | 3 | 9 | 27 | 68 | 0.76 | 0.17 | 3.30 | 6.24 | | | | |
| 75 - V Giacomet et al | 0 | 2 | 8 | 125 | 2.76 | 0.12 | 62.31 | 1.50 | | | | |
| 79 - F Gotzinger et al | 3 | 29 | 45 | 553 | 1.30 | 0.38 | 4.47 | 8.52 | | | | |
| 91 - M Kainth et al | 1 | 12 | 8 | 53 | 0.51 | 0.06 | 4.53 | 2.98 | | | | |
| PIMS-TS/MIS-C | 1 | 1 | 27 | 66 | 4.31 | 0.17 | 109.74 | 1.39 | 1.00 | 0.00 | 0.88 | 0.38 |

| Name | Events 1 | Total 1 | Events 2 | Total 2 | Effect Estimate | CI Start | CI End | Weight | P(Q) | I²(Q) | Z | P(Z) |
|---|---|---|---|---|---|---|---|---|---|---|---|---|
| 12 - OY Antunez-Montes et al | 1 | 1 | 27 | 66 | 4.31 | 0.17 | 109.74 | 1.39 | | | | |
| Death | 77 | 407 | 497 | 7462 | 3.50 | 2.23 | 5.50 | 100.00 | 0.39 | 2.69 | 5.45 | 0.00 |
| COVID-19 | 77 | 406 | 493 | 7396 | 4.93 | 1.57 | 15.50 | 98.20 | 0.25 | 26.34 | 2.73 | 0.01 |
| 12 - OY Antunez-Montes et al | 0 | 2 | 1 | 179 | 23.80 | 0.77 | 738.85 | 1.70 | | | | |
| 151 - B Sousa et al | 76 | 369 | 488 | 6567 | 3.23 | 2.47 | 4.23 | 92.37 | | | | |
| 15 - JP Armann et al | 1 | 6 | 0 | 96 | 52.64 | 1.91 | 1447.13 | 1.83 | | | | |
| 79 - F Gotzinger et al | 0 | 29 | 4 | 554 | 2.07 | 0.11 | 39.42 | 2.30 | | | | |
| PIMS-TS/MIS-C | 0 | 1 | 4 | 66 | 4.63 | 0.16 | 130.71 | 1.80 | 1.00 | 0.00 | 0.90 | 0.37 |
| 12 - OY Antunez-Montes et al | 0 | 1 | 4 | 66 | 4.63 | 0.16 | 130.71 | 1.80 | | | | |
| T21 | | | | | | | | | | | | |
| Invasive Ventilation | 20 | 93 | 761 | 8003 | 5.97 | 1.23 | 28.86 | 100.00 | 0.06 | 65.23 | 2.22 | 0.03 |
| COVID-19 | 20 | 93 | 761 | 8003 | 5.97 | 1.23 | 28.86 | 100.00 | 0.06 | 65.23 | 2.22 | 0.03 |
| 151 - B Sousa et al | 18 | 83 | 737 | 6853 | 2.30 | 1.36 | 3.89 | 50.95 | | | | |
| 26 - CE Bolaños-Almeida et al | 0 | 2 | 1 | 576 | 76.73 | 2.48 | 2376.96 | 15.17 | | | | |
| 79 - F Gotzinger et al | 2 | 8 | 23 | 574 | 7.99 | 1.53 | 41.74 | 33.88 | | | | |
| PIMS-TS/MIS-C | 0 | 0 | 0 | 0 | 0.00 | 0.00 | 0.00 | 0.00 | 0.00 | 0.00 | 0.00 | 0.00 |
| Cardiovascular Support | 2 | 10 | 18 | 1150 | 16.38 | 3.33 | 80.55 | 100.00 | 0.30 | 5.31 | 3.44 | 0.00 |
| COVID-19 | 2 | 10 | 18 | 1150 | 16.38 | 3.33 | 80.55 | 100.00 | 0.30 | 5.31 | 3.44 | 0.00 |
| 26 - CE Bolaños-Almeida et al | 0 | 2 | 1 | 576 | 76.73 | 2.48 | 2376.96 | 20.80 | | | | |
| 79 - F Gotzinger et al | 2 | 8 | 17 | 574 | 10.92 | 2.05 | 58.10 | 79.20 | | | | |
| PIMS-TS/MIS-C | 0 | 0 | 0 | 0 | 0.00 | 0.00 | 0.00 | 0.00 | 0.00 | 0.00 | 0.00 | 0.00 |
| Admission to Critical Care | 36 | 93 | 1882 | 8003 | 3.08 | 1.02 | 9.32 | 100.00 | 0.17 | 43.95 | 1.99 | 0.05 |
| COVID-19 | 36 | 93 | 1882 | 8003 | 3.08 | 1.02 | 9.32 | 100.00 | 0.17 | 43.95 | 1.99 | 0.05 |
| 151 - B Sousa et al | 34 | 83 | 1833 | 6853 | 1.90 | 1.22 | 2.95 | 62.17 | | | | |
| 26 - CE Bolaños-Almeida et al | 0 | 2 | 3 | 576 | 32.77 | 1.32 | 814.54 | 10.14 | | | | |
| 79 - F Gotzinger et al | 2 | 8 | 46 | 574 | 3.83 | 0.75 | 19.50 | 27.69 | | | | |
| PIMS-TS/MIS-C | 0 | 0 | 0 | 0 | 0.00 | 0.00 | 0.00 | 0.00 | 0.00 | 0.00 | 0.00 | 0.00 |
| Death | 15 | 93 | 555 | 8003 | 5.12 | 1.08 | 24.40 | 100.00 | 0.19 | 40.64 | 2.05 | 0.04 |
| COVID-19 | 15 | 93 | 555 | 8003 | 5.12 | 1.08 | 24.40 | 100.00 | 0.19 | 40.64 | 2.05 | 0.04 |
| 151 - B Sousa et al | 15 | 83 | 549 | 6853 | 2.53 | 1.44 | 4.46 | 63.47 | | | | |

| Name | Events 1 | Total 1 | Events 2 | Total 2 | Effect Estimate | CI Start | CI End | Weight | P(Q) | I²(Q) | Z | P(Z) |
|---|---|---|---|---|---|---|---|---|---|---|---|---|
| 26 - CE Bolaños-Almeida et al | 0 | 2 | 2 | 576 | 45.96 | 1.73 | 1222.24 | 17.06 | | | | |
| 79 - F Gotzinger et al | 0 | 8 | 4 | 574 | 7.46 | 0.37 | 149.67 | 19.47 | | | | |
| PIMS-TS/MIS-C | 0 | 0 | 0 | 0 | 0.00 | 0.00 | 0.00 | 0.00 | 0.00 | 0.00 | 0.00 | 0.00 |
| Metabolic including obesity | | | | | | | | | | | | |
| Invasive Ventilation | 30 | 148 | 804 | 9212 | 5.12 | 2.25 | 11.64 | 100.00 | 0.10 | 35.15 | 3.89 | 0.00 |
| COVID-19 | 29 | 143 | 798 | 9187 | 7.25 | 2.71 | 19.43 | 86.21 | 0.04 | 48.86 | 3.94 | 0.00 |
| 151 - B Sousa et al | 20 | 104 | 735 | 6832 | 1.98 | 1.21 | 3.24 | 25.56 | | | | |
| 15 - JP Armann et al | 0 | 1 | 6 | 101 | 4.90 | 0.18 | 132.48 | 5.09 | | | | |
| 170 - B Yayla et al | 0 | 1 | 1 | 76 | 16.78 | 0.47 | 605.19 | 4.43 | | | | |
| 174 - P Zachariah et al | 6 | 11 | 3 | 39 | 14.40 | 2.71 | 76.65 | 13.00 | | | | |
| 21 - D Bayesheva et al | 0 | 1 | 1 | 551 | 122.33 | 3.41 | 4385.74 | 4.44 | | | | |
| 28 - H Cairoli | 0 | 5 | 1 | 573 | 34.70 | 1.27 | 949.17 | 5.06 | | | | |
| 46 - M Ceano-Vivas et al | 1 | 2 | 0 | 31 | 63.00 | 1.73 | 2295.08 | 4.41 | | | | |
| 54 - A Desai et al | 2 | 14 | 25 | 279 | 1.69 | 0.36 | 8.00 | 14.05 | | | | |
| 75 - V Giacomet et al | 0 | 3 | 1 | 124 | 11.76 | 0.40 | 342.81 | 4.90 | | | | |
| 79 - F Gotzinger et al | 0 | 1 | 25 | 581 | 7.27 | 0.29 | 183.00 | 5.28 | | | | |
| PIMS-TS/MIS-C | 1 | 5 | 6 | 25 | 1.25 | 0.16 | 9.46 | 13.79 | 0.88 | 0.00 | 0.22 | 0.83 |
| 10 - Z Almoussa et al | 0 | 3 | 1 | 7 | 0.62 | 0.02 | 19.58 | 4.71 | | | | |
| 13 - AR Araujo da Silva et al | 0 | 1 | 2 | 13 | 1.53 | 0.05 | 49.80 | 4.65 | | | | |
| 9 - M Alharbi et al | 1 | 1 | 3 | 5 | 2.14 | 0.06 | 77.54 | 4.42 | | | | |
| Cardiovascular Support | 4 | 15 | 35 | 1372 | 6.78 | 0.95 | 48.48 | 100.00 | 0.12 | 45.98 | 1.91 | 0.06 |
| COVID-19 | 1 | 10 | 28 | 1362 | 15.59 | 2.93 | 83.02 | 77.06 | 0.64 | 0.00 | 3.22 | 0.00 |
| 15 - JP Armann et al | 0 | 1 | 8 | 101 | 3.67 | 0.14 | 97.13 | 19.74 | | | | |
| 170 - B Yayla et al | 0 | 1 | 0 | 76 | 0.00 | 0.00 | 0.00 | 0.00 | | | | |
| 28 - H Cairoli | 0 | 5 | 1 | 573 | 34.70 | 1.27 | 949.17 | 19.53 | | | | |
| 46 - M Ceano-Vivas et al | 1 | 2 | 0 | 31 | 63.00 | 1.73 | 2295.08 | 17.76 | | | | |
| 79 - F Gotzinger et al | 0 | 1 | 19 | 581 | 9.62 | 0.38 | 243.63 | 20.04 | | | | |
| PIMS-TS/MIS-C | 3 | 5 | 7 | 10 | 0.38 | 0.02 | 6.35 | 22.94 | 0.00 | 100.00 | 0.68 | 0.50 |
| 10 - Z Almoussa et al | 1 | 3 | 4 | 7 | 0.38 | 0.02 | 6.35 | 22.94 | | | | |
| 41 - RL Crawford et al | 2 | 2 | 3 | 3 | 0.00 | 0.00 | 0.00 | 0.00 | | | | |

| Name | Events 1 | Total 1 | Events 2 | Total 2 | Effect Estimate | CI Start | CI End | Weight | P(Q) | I²(Q) | Z | P(Z) |
|---|---|---|---|---|---|---|---|---|---|---|---|---|
| Admission to Critical Care | 279 | 494 | 2528 | 10393 | 2.08 | 1.50 | 2.88 | 100.00 | 0.24 | 17.47 | 4.40 | 0.00 |
| COVID-19 | 75 | 192 | 2031 | 9547 | 2.48 | 1.66 | 3.70 | 62.74 | 0.35 | 9.54 | 4.44 | 0.00 |
| 151 - B Sousa et al | 47 | 104 | 1820 | 6832 | 2.27 | 1.54 | 3.35 | 26.75 | | | | |
| 157 - O Swann et al | 4 | 14 | 74 | 566 | 2.66 | 0.81 | 8.70 | 6.43 | | | | |
| 15 - JP Armann et al | 1 | 1 | 14 | 101 | 18.10 | 0.70 | 466.19 | 0.98 | | | | |
| 163 - S Verma et al | 10 | 19 | 13 | 63 | 4.27 | 1.44 | 12.68 | 7.42 | | | | |
| 170 - B Yayla et al | 0 | 1 | 1 | 76 | 16.78 | 0.47 | 605.19 | 0.81 | | | | |
| 21 - D Bayesheva et al | 0 | 1 | 4 | 551 | 40.56 | 1.45 | 1135.72 | 0.93 | | | | |
| 224 - Graff et al | 4 | 15 | 7 | 51 | 2.29 | 0.57 | 9.22 | 4.84 | | | | |
| 28 - H Cairoli | 0 | 5 | 3 | 573 | 14.82 | 0.68 | 322.38 | 1.09 | | | | |
| 32 - J Chao et al | 3 | 14 | 10 | 32 | 0.60 | 0.14 | 2.63 | 4.35 | | | | |
| 46 - M Ceano-Vivas et al | 1 | 2 | 4 | 31 | 6.75 | 0.35 | 130.79 | 1.17 | | | | |
| 67 - G Fisler et al | 5 | 12 | 25 | 65 | 1.14 | 0.33 | 4.00 | 5.85 | | | | |
| 75 - V Giacomet et al | 0 | 3 | 8 | 125 | 1.97 | 0.09 | 41.44 | 1.11 | | | | |
| 79 - F Gotzinger et al | 0 | 1 | 48 | 481 | 2.98 | 0.12 | 74.15 | 1.00 | | | | |
| PIMS-TS/MIS-C | 204 | 302 | 497 | 846 | 1.45 | 1.10 | 1.92 | 37.26 | 0.77 | 0.00 | 2.64 | 0.01 |
| 10 - Z Almoussa et al | 3 | 3 | 6 | 7 | 1.62 | 0.05 | 51.11 | 0.87 | | | | |
| 13 - AR Araujo da Silva et al | 1 | 1 | 11 | 13 | 0.65 | 0.02 | 21.18 | 0.86 | | | | |
| 226 - Abrams et al | 189 | 286 | 459 | 794 | 1.42 | 1.07 | 1.89 | 32.72 | | | | |
| 41 - RL Crawford et al | 2 | 2 | 2 | 3 | 3.00 | 0.08 | 115.34 | 0.78 | | | | |
| 5 - N Abdel-Haq et al | 8 | 9 | 14 | 24 | 5.71 | 0.61 | 53.23 | 2.03 | | | | |
| 9 - M Alharbi et al | 1 | 1 | 5 | 5 | 0.00 | 0.00 | 0.00 | 0.00 | | | | |
| Death | 18 | 121 | 557 | 8735 | 8.68 | 2.32 | 32.41 | 100.00 | 0.11 | 42.47 | 3.21 | 0.00 |
| COVID-19 | 17 | 115 | 556 | 8712 | 11.65 | 2.43 | 55.85 | 88.30 | 0.06 | 52.07 | 3.07 | 0.00 |
| 151 - B Sousa et al | 16 | 104 | 548 | 6832 | 2.08 | 1.22 | 3.58 | 35.49 | | | | |
| 15 - JP Armann et al | 0 | 1 | 1 | 101 | 22.33 | 0.62 | 804.17 | 9.95 | | | | |
| 170 - B Yayla et al | 0 | 1 | 1 | 76 | 16.78 | 0.47 | 605.19 | 9.95 | | | | |
| 21 - D Bayesheva et al | 0 | 1 | 0 | 551 | 0.00 | 0.00 | 0.00 | 0.00 | | | | |
| 28 - H Cairoli | 0 | 5 | 2 | 573 | 20.78 | 0.89 | 485.13 | 11.95 | | | | |
| 46 - M Ceano-Vivas et al | 1 | 2 | 0 | 31 | 63.00 | 1.73 | 2295.08 | 9.91 | | | | |

| Name | Events 1 | Total 1 | Events 2 | Total 2 | Effect Estimate | CI Start | CI End | Weight | P(Q) | I²(Q) | Z | P(Z) |
|---|---|---|---|---|---|---|---|---|---|---|---|---|
| 79 - F Gotzinger et al | 0 | 1 | 4 | 548 | 40.33 | 1.44 | 1129.51 | 11.06 | | | | |
| PIMS-TS/MIS-C | 1 | 6 | 1 | 23 | 3.00 | 0.12 | 73.64 | 11.70 | 1.00 | 0.00 | 0.67 | 0.50 |
| 10 - Z Almoussa et al | 1 | 3 | 1 | 7 | 3.00 | 0.12 | 73.64 | 11.70 | | | | |
| 13 - AR Araujo da Silva et al | 0 | 1 | 0 | 13 | 0.00 | 0.00 | 0.00 | 0.00 | | | | |
| 41 - RL Crawford et al | 0 | 2 | 0 | 3 | 0.00 | 0.00 | 0.00 | 0.00 | | | | |
| Endocrine including DM | | | | | | | | | | | | |
| Invasive Ventilation | 4 | 24 | 43 | 1942 | 10.73 | 3.53 | 32.61 | 100.00 | 0.70 | 0.00 | 4.19 | 0.00 |
| COVID-19 | 3 | 22 | 40 | 1919 | 12.58 | 3.64 | 43.41 | 80.47 | 0.58 | 0.00 | 4.01 | 0.00 |
| 15 - JP Armann et al | 1 | 2 | 5 | 100 | 19.00 | 1.03 | 350.19 | 14.54 | | | | |
| 170 - B Yayla et al | 0 | 1 | 1 | 76 | 16.78 | 0.47 | 605.19 | 9.60 | | | | |
| 174 - P Zachariah et al | 2 | 3 | 7 | 47 | 11.43 | 0.91 | 143.64 | 19.27 | | | | |
| 21 - D Bayesheva et al | 0 | 3 | 1 | 549 | 52.24 | 1.80 | 1517.10 | 10.88 | | | | |
| 28 - H Cairoli | 0 | 6 | 1 | 572 | 29.31 | 1.09 | 788.80 | 11.39 | | | | |
| 79 - F Gotzinger et al | 0 | 7 | 25 | 575 | 1.44 | 0.08 | 25.90 | 14.78 | | | | |
| PIMS-TS/MIS-C | 1 | 2 | 3 | 23 | 5.58 | 0.45 | 69.01 | 19.53 | 0.42 | 0.00 | 1.34 | 0.18 |
| 10 - Z Almoussa et al | 0 | 1 | 1 | 9 | 1.89 | 0.05 | 72.02 | 9.31 | | | | |
| 143 - M Riollano-Cruz et al | 1 | 1 | 2 | 14 | 15.00 | 0.46 | 485.32 | 10.22 | | | | |
| Cardiovascular Support | 1 | 17 | 32 | 1332 | 4.26 | 0.87 | 20.75 | 100.00 | 0.59 | 0.00 | 1.79 | 0.07 |
| COVID-19 | 0 | 16 | 28 | 1323 | 4.44 | 0.75 | 26.41 | 78.75 | 0.37 | 0.00 | 1.64 | 0.10 |
| 15 - JP Armann et al | 0 | 2 | 8 | 100 | 2.18 | 0.10 | 49.12 | 25.80 | | | | |
| 170 - B Yayla et al | 0 | 1 | 0 | 76 | 0.00 | 0.00 | 0.00 | 0.00 | | | | |
| 28 - H Cairoli | 0 | 6 | 1 | 572 | 29.31 | 1.09 | 788.80 | 23.12 | | | | |
| 79 - F Gotzinger et al | 0 | 7 | 19 | 575 | 1.90 | 0.10 | 34.51 | 29.84 | | | | |
| PIMS-TS/MIS-C | 1 | 1 | 4 | 9 | 3.67 | 0.12 | 113.73 | 21.25 | 1.00 | 0.00 | 0.74 | 0.46 |
| 10 - Z Almoussa et al | 1 | 1 | 4 | 9 | 3.67 | 0.12 | 113.73 | 21.25 | | | | |
| Admission to Critical Care | 11 | 57 | 197 | 2568 | 2.53 | 1.22 | 5.25 | 100.00 | 0.41 | 3.07 | 2.50 | 0.01 |
| COVID-19 | 9 | 55 | 168 | 2527 | 3.06 | 1.26 | 7.41 | 91.19 | 0.29 | 17.65 | 2.47 | 0.01 |
| 157 - O Swann et al | 4 | 24 | 74 | 556 | 1.30 | 0.43 | 3.92 | 38.13 | | | | |
| 163 - S Verma et al | 1 | 1 | 22 | 81 | 7.93 | 0.31 | 201.99 | 4.99 | | | | |
| 170 - B Yayla et al | 0 | 1 | 1 | 76 | 16.78 | 0.47 | 605.19 | 4.08 | | | | |

| Name | Events 1 | Total 1 | Events 2 | Total 2 | Effect Estimate | CI Start | CI End | Weight | P(Q) | I²(Q) | Z | P(Z) |
|---|---|---|---|---|---|---|---|---|---|---|---|---|
| 21 - D Bayesheva et al | 0 | 3 | 4 | 549 | 17.32 | 0.78 | 386.35 | 5.41 | | | | |
| 224 - Graff et al | 4 | 9 | 7 | 57 | 5.71 | 1.23 | 26.50 | 20.96 | | | | |
| 28 - H Cairoli | 0 | 6 | 3 | 572 | 12.52 | 0.59 | 267.58 | 5.56 | | | | |
| 79 - F Gotzinger et al | 0 | 7 | 48 | 575 | 0.73 | 0.04 | 12.89 | 6.28 | | | | |
| 91 - M Kainth et al | 0 | 4 | 9 | 61 | 0.61 | 0.03 | 12.36 | 5.78 | | | | |
| PIMS-TS/MIS-C | 2 | 2 | 29 | 41 | 0.98 | 0.09 | 11.17 | 8.81 | 0.66 | 0.00 | 0.02 | 0.98 |
| 10 - Z Almoussa et al | 1 | 1 | 8 | 9 | 0.53 | 0.01 | 20.19 | 3.96 | | | | |
| 5 - N Abdel-Haq et al | 1 | 1 | 21 | 32 | 1.60 | 0.06 | 42.63 | 4.86 | | | | |
| Death | 0 | 20 | 9 | 1886 | 10.25 | 2.31 | 45.43 | 100.00 | 0.95 | 0.00 | 3.06 | 0.00 |
| COVID-19 | 0 | 19 | 8 | 1872 | 13.17 | 2.57 | 67.45 | 83.06 | 0.99 | 0.00 | 3.09 | 0.00 |
| 15 - JP Armann et al | 0 | 2 | 1 | 100 | 13.27 | 0.43 | 412.87 | 18.76 | | | | |
| 170 - B Yayla et al | 0 | 1 | 1 | 76 | 16.78 | 0.47 | 605.19 | 17.24 | | | | |
| 21 - D Bayesheva et al | 0 | 3 | 0 | 549 | 0.00 | 0.00 | 0.00 | 0.00 | | | | |
| 28 - H Cairoli | 0 | 6 | 2 | 572 | 17.55 | 0.76 | 402.83 | 22.58 | | | | |
| 79 - F Gotzinger et al | 0 | 7 | 4 | 575 | 8.47 | 0.42 | 171.64 | 24.48 | | | | |
| PIMS-TS/MIS-C | 0 | 1 | 1 | 14 | 3.00 | 0.08 | 111.78 | 16.94 | 1.00 | 0.00 | 0.60 | 0.55 |
| 143 - M Riollano-Cruz et al | 0 | 1 | 1 | 14 | 3.00 | 0.08 | 111.78 | 16.94 | | | | |
| CKD | | | | | | | | | | | | |
| Invasive Ventilation | 32 | 145 | 767 | 8825 | 8.81 | 2.52 | 30.79 | 100.00 | 0.03 | 53.60 | 3.41 | 0.00 |
| COVID-19 | 32 | 144 | 765 | 8812 | 10.90 | 2.72 | 43.74 | 91.16 | 0.02 | 59.74 | 3.37 | 0.00 |
| 151 - B Sousa et al | 28 | 119 | 727 | 6817 | 2.58 | 1.68 | 3.96 | 27.12 | | | | |
| 15 - JP Armann et al | 3 | 5 | 3 | 97 | 47.00 | 5.60 | 394.17 | 15.48 | | | | |
| 21 - D Bayesheva et al | 0 | 3 | 1 | 549 | 52.24 | 1.80 | 1517.10 | 9.24 | | | | |
| 28 - H Cairoli | 0 | 5 | 1 | 575 | 34.82 | 1.27 | 952.48 | 9.46 | | | | |
| 67 - G Fisler et al | 1 | 1 | 7 | 76 | 27.80 | 1.04 | 744.79 | 9.54 | | | | |
| 75 - V Giacomet et al | 0 | 2 | 1 | 125 | 16.60 | 0.53 | 516.03 | 9.00 | | | | |
| 79 - F Gotzinger et al | 0 | 9 | 25 | 573 | 1.13 | 0.06 | 19.99 | 11.32 | | | | |
| PIMS-TS/MIS-C | 0 | 1 | 2 | 13 | 1.53 | 0.05 | 49.80 | 8.84 | 1.00 | 0.00 | 0.24 | 0.81 |
| 13 - AR Araujo da Silva et al | 0 | 1 | 2 | 13 | 1.53 | 0.05 | 49.80 | 8.84 | | | | |
| Cardiovascular Support | 0 | 19 | 28 | 1243 | 3.30 | 0.35 | 31.32 | 100.00 | 0.20 | 38.78 | 1.04 | 0.30 |

| Name | Events 1 | Total 1 | Events 2 | Total 2 | Effect Estimate | CI Start | CI End | Weight | P(Q) | I²(Q) | Z | P(Z) |
|---|---|---|---|---|---|---|---|---|---|---|---|---|
| COVID-19 | 0 | 19 | 28 | 1243 | 3.30 | 0.35 | 31.32 | 100.00 | 0.20 | 38.78 | 1.04 | 0.30 |
| 15 - JP Armann et al | 0 | 5 | 8 | 97 | 0.96 | 0.05 | 18.83 | 34.27 | | | | |
| 28 - H Cairoli | 0 | 5 | 1 | 573 | 34.70 | 1.27 | 949.17 | 30.05 | | | | |
| 79 - F Gotzinger et al | 0 | 9 | 19 | 573 | 1.50 | 0.08 | 26.65 | 35.68 | | | | |
| PIMS-TS/MIS-C | 0 | 0 | 0 | 0 | 0.00 | 0.00 | 0.00 | 0.00 | 0.00 | 0.00 | 0.00 | 0.00 |
| Admission to Critical Care | 61 | 160 | 2004 | 9388 | 2.38 | 1.58 | 3.59 | 100.00 | 0.41 | 3.21 | 4.16 | 0.00 |
| COVID-19 | 60 | 159 | 1993 | 9375 | 2.63 | 1.58 | 4.39 | 98.63 | 0.35 | 9.85 | 3.70 | 0.00 |
| 151 - B Sousa et al | 51 | 119 | 1816 | 6817 | 2.07 | 1.43 | 2.98 | 73.79 | | | | |
| 157 - O Swann et al | 2 | 14 | 76 | 566 | 1.07 | 0.24 | 4.90 | 7.01 | | | | |
| 15 - JP Armann et al | 3 | 5 | 12 | 97 | 10.63 | 1.61 | 70.23 | 4.58 | | | | |
| 21 - D Bayesheva et al | 0 | 3 | 4 | 549 | 17.32 | 0.78 | 386.35 | 1.72 | | | | |
| 28 - H Cairoli | 0 | 5 | 3 | 573 | 14.82 | 0.68 | 322.38 | 1.75 | | | | |
| 67 - G Fisler et al | 2 | 2 | 28 | 75 | 8.33 | 0.39 | 179.82 | 1.76 | | | | |
| 75 - V Giacomet et al | 0 | 2 | 8 | 125 | 2.76 | 0.12 | 62.31 | 1.71 | | | | |
| 79 - F Gotzinger et al | 2 | 9 | 46 | 573 | 3.27 | 0.66 | 16.21 | 6.32 | | | | |
| PIMS-TS/MIS-C | 1 | 1 | 11 | 13 | 0.65 | 0.02 | 21.18 | 1.37 | 1.00 | 0.00 | 0.24 | 0.81 |
| 13 - AR Araujo da Silva et al | 1 | 1 | 11 | 13 | 0.65 | 0.02 | 21.18 | 1.37 | | | | |
| Death | 28 | 142 | 553 | 8622 | 3.72 | 2.44 | 5.67 | 100.00 | 0.70 | 0.00 | 6.12 | 0.00 |
| COVID-19 | 28 | 141 | 553 | 8609 | 3.72 | 2.44 | 5.67 | 100.00 | 0.70 | 0.00 | 6.12 | 0.00 |
| 151 - B Sousa et al | 28 | 119 | 546 | 6817 | 3.53 | 2.29 | 5.45 | 94.63 | | | | |
| 15 - JP Armann et al | 0 | 5 | 1 | 97 | 5.85 | 0.21 | 160.79 | 1.61 | | | | |
| 21 - D Bayesheva et al | 0 | 3 | 0 | 549 | 0.00 | 0.00 | 0.00 | 0.00 | | | | |
| 28 - H Cairoli | 0 | 5 | 2 | 573 | 20.78 | 0.89 | 485.13 | 1.78 | | | | |
| 79 - F Gotzinger et al | 0 | 9 | 4 | 573 | 6.66 | 0.33 | 132.62 | 1.98 | | | | |
| PIMS-TS/MIS-C | 0 | 1 | 0 | 13 | 0.00 | 0.00 | 0.00 | 0.00 | 0.00 | 0.00 | 0.00 | 0.00 |
| 13 - AR Araujo da Silva et al | 0 | 1 | 0 | 13 | 0.00 | 0.00 | 0.00 | 0.00 | | | | |
| Age 0 - <1 year and severe outcome | | | | | | | | | | | | |
| Invasive Ventilation | 340 | 3439 | 175 | 2482 | 0.99 | 0.56 | 1.75 | 100.00 | 0.01 | 51.30 | 0.04 | 0.97 |
| COVID-19 | 338 | 3433 | 168 | 2443 | 0.91 | 0.51 | 1.63 | 94.96 | 0.01 | 53.98 | 0.32 | 0.75 |
| 12 - OY Antunez-Montes et al | 8 | 138 | 2 | 153 | 4.65 | 0.97 | 22.27 | 8.49 | | | | |

| Name | Events 1 | Total 1 | Events 2 | Total 2 | Effect Estimate | CI Start | CI End | Weight | P(Q) | I²(Q) | Z | P(Z) |
|---|---|---|---|---|---|---|---|---|---|---|---|---|
| 13 - AR Araujo da Silva et al | 0 | 11 | 1 | 13 | 0.36 | 0.01 | 9.82 | 2.67 | | | | |
| 142 - S Richardson et al | 7 | 53 | 1 | 6 | 0.76 | 0.08 | 7.51 | 4.94 | | | | |
| 151 - B Sousa et al | 267 | 1957 | 119 | 1499 | 1.83 | 1.46 | 2.30 | 22.39 | | | | |
| 155 - Kyung Sung | 0 | 1 | 0 | 8 | 0.00 | 0.00 | 0.00 | 0.00 | | | | |
| 15 - JP Armann et al | 1 | 36 | 3 | 28 | 0.24 | 0.02 | 2.42 | 4.83 | | | | |
| 163 - S Verma et al | 1 | 33 | 1 | 9 | 0.25 | 0.01 | 4.44 | 3.39 | | | | |
| 21 - D Bayesheva et al | 0 | 100 | 0 | 137 | 0.00 | 0.00 | 0.00 | 0.00 | | | | |
| 221 - Preston et al | 43 | 632 | 22 | 245 | 0.74 | 0.43 | 1.27 | 19.29 | | | | |
| 28 - H Cairoli | 0 | 189 | 1 | 108 | 0.19 | 0.01 | 4.68 | 2.81 | | | | |
| 46 - M Ceano-Vivas et al | 1 | 15 | 0 | 2 | 0.52 | 0.02 | 16.63 | 2.44 | | | | |
| 54 - A Desai et al | 7 | 69 | 13 | 109 | 0.83 | 0.32 | 2.21 | 13.92 | | | | |
| 58 - H Du et al | 1 | 30 | 1 | 58 | 1.97 | 0.12 | 32.57 | 3.54 | | | | |
| 72 - S Garazzino et al | 2 | 52 | 0 | 24 | 2.43 | 0.11 | 52.49 | 3.03 | | | | |
| 75 - V Giacomet et al | 0 | 56 | 0 | 20 | 0.00 | 0.00 | 0.00 | 0.00 | | | | |
| 99 - A Kambhampati | 0 | 61 | 4 | 24 | 0.04 | 0.00 | 0.72 | 3.22 | | | | |
| PIMS-TS/MIS-C | 2 | 6 | 7 | 39 | 4.58 | 0.20 | 106.62 | 5.04 | 0.20 | 40.38 | 0.95 | 0.34 |
| 12 - OY Antunez-Montes et al | 2 | 2 | 6 | 31 | 19.62 | 0.84 | 460.56 | 2.89 | | | | |
| 13 - AR Araujo da Silva et al | 0 | 2 | 0 | 4 | 0.00 | 0.00 | 0.00 | 0.00 | | | | |
| 187 - R Carbajal et al | 0 | 1 | 0 | 0 | 0.00 | 0.00 | 0.00 | 0.00 | | | | |
| 89 - S Jain et al | 0 | 1 | 1 | 4 | 0.78 | 0.02 | 32.37 | 2.15 | | | | |
| Cardiovascular Support | 9 | 609 | 13 | 510 | 0.76 | 0.08 | 7.64 | 100.00 | 0.00 | 74.67 | 0.24 | 0.81 |
| COVID-19 | 3 | 467 | 5 | 471 | 1.53 | 0.03 | 80.51 | 62.71 | 0.00 | 81.55 | 0.21 | 0.83 |
| 12 - OY Antunez-Montes et al | 2 | 2 | 0 | 153 | 1535.00 | 25.08 | 93956.13 | 13.61 | | | | |
| 13 - AR Araujo da Silva et al | 0 | 11 | 0 | 13 | 0.00 | 0.00 | 0.00 | 0.00 | | | | |
| 142 - S Richardson et al | 0 | 53 | 0 | 6 | 0.00 | 0.00 | 0.00 | 0.00 | | | | |
| 15 - JP Armann et al | 0 | 36 | 4 | 28 | 0.07 | 0.00 | 1.45 | 17.17 | | | | |
| 21 - D Bayesheva et al | 0 | 100 | 0 | 137 | 0.00 | 0.00 | 0.00 | 0.00 | | | | |
| 28 - H Cairoli | 0 | 189 | 1 | 108 | 0.19 | 0.01 | 4.68 | 16.38 | | | | |
| 46 - M Ceano-Vivas et al | 1 | 15 | 0 | 2 | 0.52 | 0.02 | 16.63 | 15.55 | | | | |
| 99 - A Kambhampati | 0 | 61 | 0 | 24 | 0.00 | 0.00 | 0.00 | 0.00 | | | | |

| Name | Events 1 | Total 1 | Events 2 | Total 2 | Effect Estimate | CI Start | CI End | Weight | P(Q) | I²(Q) | Z | P(Z) |
|---|---|---|---|---|---|---|---|---|---|---|---|---|
| PIMS-TS/MIS-C | 6 | 142 | 8 | 39 | 0.18 | 0.06 | 0.55 | 37.29 | 0.42 | 0.00 | 3.00 | 0.00 |
| 12 - OY Antunez-Montes et al | 6 | 138 | 7 | 31 | 0.16 | 0.05 | 0.50 | 22.54 | | | | |
| 13 - AR Araujo da Silva et al | 0 | 2 | 0 | 4 | 0.00 | 0.00 | 0.00 | 0.00 | | | | |
| 187 - R Carbajal et al | 0 | 1 | 0 | 0 | 0.00 | 0.00 | 0.00 | 0.00 | | | | |
| 89 - S Jain et al | 0 | 1 | 1 | 4 | 0.78 | 0.02 | 32.37 | 14.75 | | | | |
| Admission to Critical Care | 947 | 4162 | 523 | 2727 | 1.15 | 0.72 | 1.85 | 100.00 | 0.00 | 72.38 | 0.60 | 0.55 |
| COVID-19 | 935 | 4144 | 499 | 2665 | 1.11 | 0.67 | 1.83 | 91.00 | 0.00 | 76.20 | 0.40 | 0.69 |
| 12 - OY Antunez-Montes et al | 16 | 138 | 0 | 153 | 41.35 | 2.46 | 696.17 | 2.19 | | | | |
| 136 - G Qian et al | 5 | 28 | 1 | 16 | 3.26 | 0.35 | 30.73 | 3.07 | | | | |
| 13 - AR Araujo da Silva et al | 6 | 11 | 0 | 13 | 31.91 | 1.52 | 668.75 | 1.94 | | | | |
| 142 - S Richardson et al | 17 | 53 | 3 | 6 | 0.47 | 0.09 | 2.59 | 4.36 | | | | |
| 151 - B Sousa et al | 626 | 1957 | 342 | 1499 | 1.59 | 1.36 | 1.86 | 9.88 | | | | |
| 153 - P Storch de Gracia et al | 2 | 12 | 1 | 5 | 0.80 | 0.06 | 11.50 | 2.39 | | | | |
| 155 - Kyung Sung | 0 | 1 | 0 | 8 | 0.00 | 0.00 | 0.00 | 0.00 | | | | |
| 157 - O Swann et al | 31 | 217 | 15 | 89 | 0.82 | 0.42 | 1.61 | 8.31 | | | | |
| 15 - JP Armann et al | 3 | 36 | 4 | 28 | 0.55 | 0.11 | 2.67 | 4.70 | | | | |
| 162 - MM van der Zalm et al | 5 | 30 | 3 | 21 | 1.20 | 0.25 | 5.68 | 4.80 | | | | |
| 163 - S Verma et al | 7 | 33 | 1 | 9 | 2.15 | 0.23 | 20.23 | 3.08 | | | | |
| 19 - S Bellino et al | 5 | 193 | 8 | 84 | 0.25 | 0.08 | 0.80 | 6.28 | | | | |
| 21 - D Bayesheva et al | 0 | 100 | 1 | 137 | 0.45 | 0.02 | 11.23 | 1.78 | | | | |
| 221 - Preston et al | 152 | 632 | 103 | 245 | 0.44 | 0.32 | 0.60 | 9.57 | | | | |
| 224 - Graff et al | 1 | 15 | 0 | 10 | 2.17 | 0.08 | 58.76 | 1.70 | | | | |
| 26 - CE Bolaños-Almeida et al | 7 | 58 | 2 | 103 | 6.93 | 1.39 | 34.58 | 4.64 | | | | |
| 28 - H Cairoli | 0 | 189 | 1 | 108 | 0.19 | 0.01 | 4.68 | 1.78 | | | | |
| 2 - Centre for Disease Control | 5 | 60 | 0 | 15 | 3.07 | 0.16 | 58.65 | 2.04 | | | | |
| 46 - M Ceano-Vivas et al | 2 | 15 | 0 | 2 | 0.93 | 0.03 | 25.68 | 1.68 | | | | |
| 75 - V Giacomet et al | 3 | 56 | 1 | 20 | 1.08 | 0.11 | 10.98 | 2.93 | | | | |
| 79 - F Gotzinger et al | 23 | 230 | 2 | 62 | 3.33 | 0.76 | 14.54 | 5.07 | | | | |
| 91 - M Kainth et al | 0 | 19 | 2 | 8 | 0.07 | 0.00 | 1.58 | 1.83 | | | | |
| 99 - A Kambhampati | 19 | 61 | 9 | 24 | 0.75 | 0.28 | 2.03 | 6.95 | | | | |

| Name | Events 1 | Total 1 | Events 2 | Total 2 | Effect Estimate | CI Start | CI End | Weight | P(Q) | I²(Q) | Z | P(Z) |
|---|---|---|---|---|---|---|---|---|---|---|---|---|
| PIMS-TS/MIS-C | 12 | 18 | 24 | 62 | 1.77 | 0.45 | 6.94 | 9.00 | 0.58 | 0.00 | 0.82 | 0.41 |
| 12 - OY Antunez-Montes et al | 2 | 2 | 11 | 31 | 8.91 | 0.39 | 202.06 | 1.87 | | | | |
| 13 - AR Araujo da Silva et al | 2 | 2 | 3 | 4 | 2.14 | 0.06 | 77.54 | 1.48 | | | | |
| 179 - J Pang et al | 4 | 5 | 0 | 0 | 0.00 | 0.00 | 0.00 | 0.00 | | | | |
| 187 - R Carbajal et al | 1 | 1 | 0 | 0 | 0.00 | 0.00 | 0.00 | 0.00 | | | | |
| 38 - BC Clark et al | 2 | 4 | 6 | 17 | 1.83 | 0.20 | 16.51 | 3.16 | | | | |
| 5 - N Abdel-Haq et al | 1 | 4 | 4 | 10 | 0.50 | 0.04 | 6.68 | 2.49 | | | | |
| Death | 399 | 4380 | 209 | 4272 | 2.10 | 1.54 | 2.85 | 100.00 | 0.24 | 20.42 | 4.73 | 0.00 |
| COVID-19 | 398 | 4375 | 207 | 4237 | 2.07 | 1.54 | 2.78 | 99.05 | 0.26 | 19.42 | 4.84 | 0.00 |
| 12 - OY Antunez-Montes et al | 3 | 138 | 0 | 153 | 7.93 | 0.41 | 154.91 | 1.05 | | | | |
| 13 - AR Araujo da Silva et al | 0 | 11 | 0 | 13 | 0.00 | 0.00 | 0.00 | 0.00 | | | | |
| 142 - S Richardson et al | 0 | 53 | 1 | 6 | 0.03 | 0.00 | 0.95 | 0.84 | | | | |
| 151 - B Sousa et al | 181 | 1957 | 67 | 1499 | 2.18 | 1.63 | 2.91 | 38.44 | | | | |
| 155 - Kyung Sung | 0 | 1 | 0 | 8 | 0.00 | 0.00 | 0.00 | 0.00 | | | | |
| 157 - O Swann et al | 3 | 217 | 0 | 89 | 2.92 | 0.15 | 57.13 | 1.05 | | | | |
| 15 - JP Armann et al | 0 | 36 | 1 | 28 | 0.25 | 0.01 | 6.40 | 0.88 | | | | |
| 21 - D Bayesheva et al | 0 | 100 | 0 | 137 | 0.00 | 0.00 | 0.00 | 0.00 | | | | |
| 227 - Sena et al | 15 | 111 | 6 | 67 | 1.59 | 0.58 | 4.32 | 8.11 | | | | |
| 26 - CE Bolaños-Almeida et al | 3 | 58 | 2 | 103 | 2.75 | 0.45 | 16.99 | 2.71 | | | | |
| 28 - H Cairoli | 0 | 189 | 1 | 108 | 0.19 | 0.01 | 4.68 | 0.90 | | | | |
| 46 - M Ceano-Vivas et al | 1 | 15 | 0 | 2 | 0.52 | 0.02 | 16.63 | 0.77 | | | | |
| 58 - H Du et al | 1 | 30 | 0 | 58 | 5.95 | 0.24 | 150.55 | 0.89 | | | | |
| 75 - V Giacomet et al | 0 | 56 | 0 | 20 | 0.00 | 0.00 | 0.00 | 0.00 | | | | |
| 86 - D Hillesheim et al | 191 | 1341 | 129 | 1918 | 2.30 | 1.82 | 2.91 | 43.41 | | | | |
| 89 - S Jain et al | 0 | 1 | 0 | 4 | 0.00 | 0.00 | 0.00 | 0.00 | | | | |
| 99 - A Kambhampati | 0 | 61 | 0 | 24 | 0.00 | 0.00 | 0.00 | 0.00 | | | | |
| PIMS-TS/MIS-C | 1 | 5 | 2 | 35 | 14.50 | 0.64 | 328.46 | 0.95 | 1.00 | 0.00 | 1.68 | 0.09 |
| 12 - OY Antunez-Montes et al | 1 | 2 | 2 | 31 | 14.50 | 0.64 | 328.46 | 0.95 | | | | |
| 13 - AR Araujo da Silva et al | 0 | 2 | 0 | 4 | 0.00 | 0.00 | 0.00 | 0.00 | | | | |
| 187 - R Carbajal et al | 0 | 1 | 0 | 0 | 0.00 | 0.00 | 0.00 | 0.00 | | | | |

| Name | Events 1 | Total 1 | Events 2 | Total 2 | Effect Estimate | CI Start | CI End | Weight | P(Q) | I²(Q) | Z | P(Z) |
|---|---|---|---|---|---|---|---|---|---|---|---|---|
| Age 5-9 years | | | | | | | | | | | | |
| Invasive Ventilation | 134 | 1846 | 170 | 2454 | 1.05 | 0.82 | 1.34 | 100.00 | 0.96 | 0.00 | 0.39 | 0.70 |
| COVID-19 | 111 | 1770 | 163 | 2410 | 0.99 | 0.77 | 1.28 | 92.50 | 0.89 | 0.00 | 0.06 | 0.95 |
| 12 - OY Antunez-Montes et al | 0 | 128 | 2 | 153 | 0.24 | 0.01 | 4.96 | 0.66 | | | | |
| 13 - AR Araujo da Silva et al | 3 | 12 | 1 | 13 | 4.00 | 0.35 | 45.10 | 1.04 | | | | |
| 142 - S Richardson et al | 1 | 6 | 1 | 6 | 1.00 | 0.05 | 20.83 | 0.66 | | | | |
| 151 - B Sousa et al | 81 | 964 | 119 | 1499 | 1.06 | 0.79 | 1.43 | 69.97 | | | | |
| 155 - Kyung Sung | 0 | 12 | 0 | 8 | 0.00 | 0.00 | 0.00 | 0.00 | | | | |
| 15 - JP Armann et al | 1 | 12 | 3 | 28 | 0.76 | 0.07 | 8.12 | 1.08 | | | | |
| 187 - R Carbajal et al | 2 | 4 | 0 | 0 | 0.00 | 0.00 | 0.00 | 0.00 | | | | |
| 21 - D Bayesheva et al | 0 | 128 | 0 | 137 | 0.00 | 0.00 | 0.00 | 0.00 | | | | |
| 221 - Preston et al | 20 | 274 | 22 | 245 | 0.80 | 0.42 | 1.50 | 15.23 | | | | |
| 28 - H Cairoli | 0 | 109 | 1 | 108 | 0.33 | 0.01 | 8.12 | 0.59 | | | | |
| 46 - M Ceano-Vivas et al | 0 | 4 | 0 | 2 | 0.00 | 0.00 | 0.00 | 0.00 | | | | |
| 54 - A Desai et al | 2 | 26 | 13 | 109 | 0.62 | 0.13 | 2.91 | 2.51 | | | | |
| 58 - H Du et al | 1 | 54 | 1 | 58 | 1.08 | 0.07 | 17.63 | 0.78 | | | | |
| 72 - S Garazzino et al | 0 | 13 | 0 | 24 | 0.00 | 0.00 | 0.00 | 0.00 | | | | |
| 75 - V Giacomet et al | 0 | 24 | 0 | 20 | 0.00 | 0.00 | 0.00 | 0.00 | | | | |
| PIMS-TS/MIS-C | 23 | 76 | 7 | 44 | 2.11 | 0.86 | 5.19 | 7.50 | 1.00 | 0.00 | 1.62 | 0.10 |
| 10 - Z Almoussa et al | 1 | 4 | 0 | 0 | 0.00 | 0.00 | 0.00 | 0.00 | | | | |
| 12 - OY Antunez-Montes et al | 11 | 33 | 6 | 31 | 2.08 | 0.66 | 6.57 | 4.61 | | | | |
| 13 - AR Araujo da Silva et al | 1 | 6 | 0 | 4 | 2.45 | 0.08 | 76.13 | 0.52 | | | | |
| 143 - M Riollano-Cruz et al | 1 | 3 | 0 | 1 | 1.80 | 0.04 | 79.42 | 0.42 | | | | |
| 147 - L Shahbaznejad et al | 1 | 4 | 0 | 0 | 0.00 | 0.00 | 0.00 | 0.00 | | | | |
| 183 - Rekhtman et al | 2 | 6 | 0 | 2 | 2.78 | 0.09 | 83.84 | 0.52 | | | | |
| 228 - Hasan et al | 1 | 5 | 0 | 2 | 1.67 | 0.05 | 58.28 | 0.48 | | | | |
| 41 - RL Crawford et al | 0 | 3 | 0 | 0 | 0.00 | 0.00 | 0.00 | 0.00 | | | | |
| 89 - S Jain et al | 5 | 12 | 1 | 4 | 2.14 | 0.17 | 27.10 | 0.94 | | | | |
| Cardiovascular Support | 31 | 340 | 15 | 356 | 1.53 | 0.70 | 3.36 | 100.00 | 0.81 | 0.00 | 1.07 | 0.29 |
| COVID-19 | 3 | 271 | 5 | 310 | 1.35 | 0.31 | 6.01 | 27.76 | 0.32 | 0.00 | 0.40 | 0.69 |

| Name | Events 1 | Total 1 | Events 2 | Total 2 | Effect Estimate | CI Start | CI End | Weight | P(Q) | I²(Q) | Z | P(Z) |
|---|---|---|---|---|---|---|---|---|---|---|---|---|
| 12 - OY Antunez-Montes et al | 0 | 128 | 0 | 153 | 0.00 | 0.00 | 0.00 | 0.00 | | | | |
| 13 - AR Araujo da Silva et al | 0 | 12 | 0 | 13 | 0.00 | 0.00 | 0.00 | 0.00 | | | | |
| 142 - S Richardson et al | 0 | 6 | 0 | 6 | 0.00 | 0.00 | 0.00 | 0.00 | | | | |
| 15 - JP Armann et al | 3 | 12 | 4 | 28 | 2.00 | 0.37 | 10.75 | 21.79 | | | | |
| 28 - H Cairoli | 0 | 109 | 1 | 108 | 0.33 | 0.01 | 8.12 | 5.97 | | | | |
| 46 - M Ceano-Vivas et al | 0 | 4 | 0 | 2 | 0.00 | 0.00 | 0.00 | 0.00 | | | | |
| PIMS-TS/MIS-C | 28 | 69 | 10 | 46 | 1.61 | 0.64 | 4.05 | 72.24 | 0.74 | 0.00 | 1.01 | 0.31 |
| 10 - Z Almoussa et al | 2 | 4 | 1 | 2 | 1.00 | 0.03 | 29.81 | 5.35 | | | | |
| 12 - OY Antunez-Montes et al | 9 | 33 | 7 | 31 | 1.29 | 0.41 | 4.01 | 47.55 | | | | |
| 13 - AR Araujo da Silva et al | 0 | 6 | 0 | 4 | 0.00 | 0.00 | 0.00 | 0.00 | | | | |
| 143 - M Riollano-Cruz et al | 1 | 3 | 0 | 1 | 1.80 | 0.04 | 79.42 | 4.30 | | | | |
| 147 - L Shahbaznejad et al | 1 | 4 | 1 | 4 | 1.00 | 0.04 | 24.55 | 6.02 | | | | |
| 187 - R Carbajal et al | 4 | 4 | 0 | 0 | 0.00 | 0.00 | 0.00 | 0.00 | | | | |
| 41 - RL Crawford et al | 2 | 3 | 0 | 0 | 0.00 | 0.00 | 0.00 | 0.00 | | | | |
| 89 - S Jain et al | 9 | 12 | 1 | 4 | 9.00 | 0.66 | 122.79 | 9.02 | | | | |
| Admission to Critical Care | 1119 | 3335 | 911 | 3582 | 1.46 | 1.19 | 1.78 | 100.00 | 0.13 | 22.17 | 3.68 | 0.00 |
| COVID-19 | 437 | 2184 | 508 | 2682 | 1.17 | 1.00 | 1.36 | 56.90 | 0.63 | 0.00 | 1.97 | 0.05 |
| 100 - M Korkomaz et al | 0 | 24 | 2 | 20 | 0.15 | 0.01 | 3.34 | 0.41 | | | | |
| 12 - OY Antunez-Montes et al | 6 | 128 | 3 | 153 | 2.46 | 0.60 | 10.04 | 1.88 | | | | |
| 136 - G Qian et al | 1 | 45 | 1 | 16 | 0.34 | 0.02 | 5.79 | 0.49 | | | | |
| 13 - AR Araujo da Silva et al | 7 | 12 | 6 | 13 | 1.63 | 0.34 | 7.95 | 1.51 | | | | |
| 142 - S Richardson et al | 2 | 6 | 3 | 6 | 0.50 | 0.05 | 5.15 | 0.72 | | | | |
| 151 - B Sousa et al | 243 | 964 | 342 | 1499 | 1.14 | 0.94 | 1.38 | 20.73 | | | | |
| 153 - P Storch de Gracia et al | 6 | 9 | 1 | 5 | 8.00 | 0.60 | 106.94 | 0.58 | | | | |
| 155 - Kyung Sung | 0 | 12 | 0 | 8 | 0.00 | 0.00 | 0.00 | 0.00 | | | | |
| 157 - O Swann et al | 18 | 73 | 15 | 89 | 1.61 | 0.75 | 3.48 | 5.35 | | | | |
| 15 - JP Armann et al | 4 | 12 | 4 | 28 | 3.00 | 0.61 | 14.86 | 1.48 | | | | |
| 162 - MM van der Zalm et al | 3 | 11 | 3 | 21 | 2.25 | 0.37 | 13.67 | 1.18 | | | | |
| 187 - R Carbajal et al | 4 | 4 | 0 | 0 | 0.00 | 0.00 | 0.00 | 0.00 | | | | |
| 19 - S Bellino et al | 1 | 97 | 8 | 84 | 0.10 | 0.01 | 0.81 | 0.88 | | | | |

| Name | Events 1 | Total 1 | Events 2 | Total 2 | Effect Estimate | CI Start | CI End | Weight | P(Q) | I²(Q) | Z | P(Z) |
|---|---|---|---|---|---|---|---|---|---|---|---|---|
| 21 - D Bayesheva et al | 1 | 128 | 1 | 137 | 1.07 | 0.07 | 17.30 | 0.51 | | | | |
| 221 - Preston et al | 119 | 274 | 103 | 245 | 1.06 | 0.75 | 1.50 | 14.37 | | | | |
| 224 - Graff et al | 1 | 7 | 0 | 10 | 4.85 | 0.17 | 137.68 | 0.35 | | | | |
| 26 - CE Bolaños-Almeida et al | 3 | 115 | 2 | 103 | 1.35 | 0.22 | 8.26 | 1.17 | | | | |
| 28 - H Cairoli | 2 | 109 | 1 | 108 | 2.00 | 0.18 | 22.39 | 0.67 | | | | |
| 2 - Centre for Disease Control | 2 | 16 | 0 | 15 | 5.34 | 0.24 | 121.00 | 0.41 | | | | |
| 46 - M Ceano-Vivas et al | 1 | 4 | 0 | 2 | 2.14 | 0.06 | 77.54 | 0.31 | | | | |
| 67 - G Fisler et al | 2 | 9 | 8 | 30 | 0.79 | 0.13 | 4.60 | 1.22 | | | | |
| 75 - V Giacomet et al | 2 | 24 | 1 | 20 | 1.73 | 0.14 | 20.58 | 0.64 | | | | |
| 79 - F Gotzinger et al | 8 | 94 | 2 | 62 | 2.79 | 0.57 | 13.61 | 1.50 | | | | |
| 91 - M Kainth et al | 1 | 7 | 2 | 8 | 0.50 | 0.04 | 7.10 | 0.56 | | | | |
| PIMS-TS/MIS-C | 682 | 1151 | 403 | 900 | 1.81 | 1.51 | 2.16 | 43.10 | 0.49 | 0.00 | 6.48 | 0.00 |
| 10 - Z Almoussa et al | 4 | 4 | 2 | 2 | 0.00 | 0.00 | 0.00 | 0.00 | | | | |
| 12 - OY Antunez-Montes et al | 12 | 33 | 11 | 31 | 1.04 | 0.37 | 2.89 | 3.34 | | | | |
| 13 - AR Araujo da Silva et al | 5 | 6 | 3 | 4 | 1.67 | 0.07 | 37.73 | 0.41 | | | | |
| 143 - M Riollano-Cruz et al | 1 | 3 | 0 | 1 | 1.80 | 0.04 | 79.42 | 0.28 | | | | |
| 147 - L Shahbaznejad et al | 3 | 4 | 4 | 4 | 0.26 | 0.01 | 8.52 | 0.32 | | | | |
| 183 - Rekhtman et al | 2 | 6 | 1 | 2 | 0.50 | 0.02 | 12.90 | 0.37 | | | | |
| 222 - Belay et al | 349 | 588 | 197 | 445 | 1.84 | 1.43 | 2.36 | 18.24 | | | | |
| 225 - Alkan et al | 0 | 14 | 0 | 9 | 0.00 | 0.00 | 0.00 | 0.00 | | | | |
| 226 - Abrams et al | 278 | 456 | 175 | 373 | 1.77 | 1.34 | 2.33 | 17.10 | | | | |
| 228 - Hasan et al | 5 | 5 | 0 | 2 | 55.00 | 0.83 | 3650.69 | 0.23 | | | | |
| 38 - BC Clark et al | 9 | 15 | 6 | 17 | 2.75 | 0.66 | 11.54 | 1.81 | | | | |
| 41 - RL Crawford et al | 2 | 3 | 0 | 0 | 0.00 | 0.00 | 0.00 | 0.00 | | | | |
| 5 - N Abdel-Haq et al | 12 | 14 | 4 | 10 | 9.00 | 1.27 | 63.89 | 1.00 | | | | |
| Death | 140 | 3284 | 215 | 4565 | 1.07 | 0.86 | 1.33 | 100.00 | 0.70 | 0.00 | 0.57 | 0.57 |
| COVID-19 | 133 | 2637 | 207 | 4076 | 1.09 | 0.87 | 1.37 | 95.57 | 0.60 | 0.00 | 0.77 | 0.44 |
| 12 - OY Antunez-Montes et al | 2 | 128 | 0 | 153 | 6.07 | 0.29 | 127.53 | 0.52 | | | | |
| 13 - AR Araujo da Silva et al | 1 | 12 | 0 | 13 | 3.52 | 0.13 | 95.09 | 0.45 | | | | |
| 142 - S Richardson et al | 0 | 6 | 1 | 6 | 0.28 | 0.01 | 8.42 | 0.42 | | | | |

| Name | Events 1 | Total 1 | Events 2 | Total 2 | Effect Estimate | CI Start | CI End | Weight | P(Q) | I²(Q) | Z | P(Z) |
|---|---|---|---|---|---|---|---|---|---|---|---|---|
| 151 - B Sousa et al | 56 | 964 | 67 | 1499 | 1.32 | 0.92 | 1.90 | 36.60 | | | | |
| 155 - Kyung Sung | 0 | 12 | 0 | 8 | 0.00 | 0.00 | 0.00 | 0.00 | | | | |
| 157 - O Swann et al | 0 | 73 | 0 | 89 | 0.00 | 0.00 | 0.00 | 0.00 | | | | |
| 15 - JP Armann et al | 0 | 12 | 1 | 28 | 0.73 | 0.03 | 19.29 | 0.45 | | | | |
| 187 - R Carbajal et al | 0 | 4 | 0 | 0 | 0.00 | 0.00 | 0.00 | 0.00 | | | | |
| 227 - Sena et al | 5 | 33 | 6 | 67 | 1.82 | 0.51 | 6.45 | 3.02 | | | | |
| 26 - CE Bolaños-Almeida et al | 0 | 115 | 2 | 103 | 0.18 | 0.01 | 3.70 | 0.52 | | | | |
| 28 - H Cairoli | 0 | 109 | 1 | 108 | 0.33 | 0.01 | 8.12 | 0.47 | | | | |
| 46 - M Ceano-Vivas et al | 0 | 4 | 0 | 2 | 0.00 | 0.00 | 0.00 | 0.00 | | | | |
| 58 - H Du et al | 0 | 64 | 0 | 58 | 0.00 | 0.00 | 0.00 | 0.00 | | | | |
| 75 - V Giacomet et al | 0 | 24 | 0 | 20 | 0.00 | 0.00 | 0.00 | 0.00 | | | | |
| 86 - D Hillesheim et al | 68 | 1073 | 129 | 1918 | 0.94 | 0.69 | 1.27 | 52.71 | | | | |
| 89 - S Jain et al | 1 | 4 | 0 | 4 | 3.86 | 0.12 | 126.73 | 0.40 | | | | |
| PIMS-TS/MIS-C | 7 | 647 | 8 | 489 | 0.63 | 0.22 | 1.78 | 4.43 | 0.67 | 0.00 | 0.88 | 0.38 |
| 10 - Z Almoussa et al | 1 | 4 | 0 | 2 | 2.14 | 0.06 | 77.54 | 0.38 | | | | |
| 12 - OY Antunez-Montes et al | 1 | 33 | 2 | 31 | 0.45 | 0.04 | 5.26 | 0.81 | | | | |
| 13 - AR Araujo da Silva et al | 0 | 6 | 0 | 4 | 0.00 | 0.00 | 0.00 | 0.00 | | | | |
| 143 - M Riollano-Cruz et al | 1 | 3 | 0 | 1 | 1.80 | 0.04 | 79.42 | 0.34 | | | | |
| 147 - L Shahbaznejad et al | 1 | 4 | 0 | 4 | 3.86 | 0.12 | 126.73 | 0.40 | | | | |
| 183 - Rekhtman et al | 0 | 6 | 0 | 2 | 0.00 | 0.00 | 0.00 | 0.00 | | | | |
| 222 - Belay et al | 3 | 588 | 6 | 445 | 0.38 | 0.09 | 1.51 | 2.51 | | | | |
| 41 - RL Crawford et al | 0 | 3 | 0 | 0 | 0.00 | 0.00 | 0.00 | 0.00 | | | | |
| Age 10-14 years | | | | | | | | | | | | |
| Invasive Ventilation | 134 | 1424 | 157 | 2140 | 1.35 | 1.05 | 1.74 | 100.00 | 0.63 | 0.00 | 2.37 | 0.02 |
| COVID-19 | 113 | 1332 | 146 | 2061 | 1.30 | 1.00 | 1.68 | 91.24 | 0.50 | 0.00 | 1.93 | 0.05 |
| 12 - OY Antunez-Montes et al | 0 | 80 | 2 | 153 | 0.38 | 0.02 | 7.93 | 0.68 | | | | |
| 13 - AR Araujo da Silva et al | 0 | 8 | 1 | 13 | 0.49 | 0.02 | 13.52 | 0.57 | | | | |
| 142 - S Richardson et al | 2 | 11 | 1 | 6 | 1.11 | 0.08 | 15.53 | 0.91 | | | | |
| 151 - B Sousa et al | 97 | 859 | 119 | 1499 | 1.48 | 1.11 | 1.96 | 79.09 | | | | |
| 155 - Kyung Sung | 0 | 18 | 0 | 8 | 0.00 | 0.00 | 0.00 | 0.00 | | | | |

| Name | Events 1 | Total 1 | Events 2 | Total 2 | Effect Estimate | CI Start | CI End | Weight | P(Q) | I²(Q) | Z | P(Z) |
|---|---|---|---|---|---|---|---|---|---|---|---|---|
| 15 - JP Armann et al | 0 | 10 | 3 | 28 | 0.35 | 0.02 | 7.32 | 0.68 | | | | |
| 163 - S Verma et al | 3 | 19 | 1 | 9 | 1.50 | 0.13 | 16.82 | 1.08 | | | | |
| 28 - H Cairoli | 0 | 101 | 1 | 108 | 0.35 | 0.01 | 8.77 | 0.61 | | | | |
| 46 - M Ceano-Vivas et al | 0 | 5 | 0 | 2 | 0.00 | 0.00 | 0.00 | 0.00 | | | | |
| 54 - A Desai et al | 2 | 32 | 13 | 109 | 0.49 | 0.11 | 2.31 | 2.64 | | | | |
| 58 - H Du et al | 0 | 30 | 1 | 58 | 0.63 | 0.02 | 15.90 | 0.60 | | | | |
| 72 - S Garazzino et al | 0 | 21 | 0 | 24 | 0.00 | 0.00 | 0.00 | 0.00 | | | | |
| 75 - V Giacomet et al | 1 | 15 | 0 | 20 | 4.24 | 0.16 | 111.65 | 0.59 | | | | |
| 99 - A Kambhampati | 8 | 123 | 4 | 24 | 0.35 | 0.10 | 1.26 | 3.78 | | | | |
| PIMS-TS/MIS-C | 21 | 92 | 11 | 79 | 2.15 | 0.92 | 5.02 | 8.76 | 0.69 | 0.00 | 1.77 | 0.08 |
| 10 - Z Almoussa et al | 0 | 4 | 0 | 2 | 0.00 | 0.00 | 0.00 | 0.00 | | | | |
| 12 - OY Antunez-Montes et al | 8 | 18 | 6 | 31 | 3.33 | 0.92 | 12.08 | 3.80 | | | | |
| 13 - AR Araujo da Silva et al | 1 | 1 | 0 | 4 | 27.00 | 0.35 | 2057.98 | 0.34 | | | | |
| 143 - M Riollano-Cruz et al | 1 | 7 | 0 | 1 | 0.69 | 0.02 | 26.90 | 0.47 | | | | |
| 147 - L Shahbaznejad et al | 1 | 2 | 1 | 4 | 3.00 | 0.08 | 107.45 | 0.49 | | | | |
| 183 - Rekhtman et al | 3 | 10 | 0 | 2 | 2.33 | 0.09 | 62.68 | 0.58 | | | | |
| 187 - R Carbajal et al | 1 | 2 | 0 | 0 | 0.00 | 0.00 | 0.00 | 0.00 | | | | |
| 60 - EM Dufort et al | 3 | 42 | 3 | 31 | 0.72 | 0.13 | 3.82 | 2.25 | | | | |
| 89 - S Jain et al | 3 | 6 | 1 | 4 | 3.00 | 0.19 | 47.96 | 0.82 | | | | |
| Cardiovascular Support | 53 | 420 | 30 | 411 | 1.84 | 0.96 | 3.54 | 100.00 | 0.79 | 0.00 | 1.84 | 0.07 |
| COVID-19 | 11 | 338 | 5 | 334 | 1.02 | 0.21 | 4.91 | 17.22 | 0.44 | 0.00 | 0.02 | 0.98 |
| 12 - OY Antunez-Montes et al | 0 | 80 | 0 | 153 | 0.00 | 0.00 | 0.00 | 0.00 | | | | |
| 13 - AR Araujo da Silva et al | 0 | 8 | 0 | 13 | 0.00 | 0.00 | 0.00 | 0.00 | | | | |
| 142 - S Richardson et al | 0 | 11 | 0 | 6 | 0.00 | 0.00 | 0.00 | 0.00 | | | | |
| 15 - JP Armann et al | 1 | 10 | 4 | 28 | 0.67 | 0.07 | 6.79 | 7.91 | | | | |
| 28 - H Cairoli | 0 | 101 | 1 | 108 | 0.35 | 0.01 | 8.77 | 4.13 | | | | |
| 46 - M Ceano-Vivas et al | 0 | 5 | 0 | 2 | 0.00 | 0.00 | 0.00 | 0.00 | | | | |
| 99 - A Kambhampati | 10 | 123 | 0 | 24 | 4.53 | 0.26 | 79.98 | 5.18 | | | | |
| PIMS-TS/MIS-C | 42 | 82 | 25 | 77 | 2.09 | 1.02 | 4.28 | 82.78 | 0.80 | 0.00 | 2.01 | 0.04 |
| 10 - Z Almoussa et al | 2 | 4 | 1 | 2 | 1.00 | 0.03 | 29.81 | 3.70 | | | | |

| Name | Events 1 | Total 1 | Events 2 | Total 2 | Effect Estimate | CI Start | CI End | Weight | P(Q) | I²(Q) | Z | P(Z) |
|---|---|---|---|---|---|---|---|---|---|---|---|---|
| 12 - OY Antunez-Montes et al | 5 | 18 | 7 | 31 | 1.32 | 0.35 | 4.99 | 24.06 | | | | |
| 13 - AR Araujo da Silva et al | 0 | 1 | 0 | 4 | 0.00 | 0.00 | 0.00 | 0.00 | | | | |
| 143 - M Riollano-Cruz et al | 0 | 7 | 0 | 1 | 0.00 | 0.00 | 0.00 | 0.00 | | | | |
| 147 - L Shahbaznejad et al | 2 | 2 | 1 | 4 | 11.67 | 0.32 | 422.14 | 3.31 | | | | |
| 187 - R Carbajal et al | 1 | 2 | 0 | 0 | 0.00 | 0.00 | 0.00 | 0.00 | | | | |
| 60 - EM Dufort et al | 29 | 42 | 15 | 31 | 2.38 | 0.91 | 6.22 | 46.15 | | | | |
| 89 - S Jain et al | 3 | 6 | 1 | 4 | 3.00 | 0.19 | 47.96 | 5.55 | | | | |
| Admission to Critical Care | 783 | 2476 | 649 | 2901 | 2.02 | 1.45 | 2.81 | 100.00 | 0.01 | 40.94 | 4.18 | 0.00 |
| COVID-19 | 410 | 1910 | 402 | 2345 | 1.78 | 1.19 | 2.68 | 68.25 | 0.10 | 30.53 | 2.78 | 0.01 |
| 12 - OY Antunez-Montes et al | 0 | 80 | 3 | 153 | 0.27 | 0.01 | 5.23 | 1.15 | | | | |
| 136 - G Qian et al | 0 | 38 | 1 | 16 | 0.13 | 0.01 | 3.48 | 0.97 | | | | |
| 13 - AR Araujo da Silva et al | 5 | 8 | 6 | 13 | 1.94 | 0.32 | 11.76 | 2.82 | | | | |
| 142 - S Richardson et al | 6 | 11 | 3 | 6 | 1.20 | 0.16 | 8.80 | 2.37 | | | | |
| 151 - B Sousa et al | 257 | 859 | 342 | 1499 | 1.44 | 1.20 | 1.75 | 16.49 | | | | |
| 153 - P Storch de Gracia et al | 6 | 13 | 1 | 5 | 3.43 | 0.30 | 39.64 | 1.65 | | | | |
| 155 - Kyung Sung | 0 | 18 | 0 | 8 | 0.00 | 0.00 | 0.00 | 0.00 | | | | |
| 157 - O Swann et al | 33 | 58 | 15 | 89 | 6.51 | 3.04 | 13.93 | 9.06 | | | | |
| 15 - JP Armann et al | 1 | 10 | 4 | 28 | 0.67 | 0.07 | 6.79 | 1.81 | | | | |
| 163 - S Verma et al | 6 | 19 | 1 | 9 | 3.69 | 0.37 | 36.57 | 1.85 | | | | |
| 21 - D Bayesheva et al | 2 | 101 | 1 | 137 | 2.75 | 0.25 | 30.72 | 1.69 | | | | |
| 224 - Graff et al | 1 | 11 | 0 | 10 | 3.00 | 0.11 | 82.40 | 0.94 | | | | |
| 26 - CE Bolaños-Almeida et al | 2 | 148 | 2 | 103 | 0.69 | 0.10 | 4.99 | 2.40 | | | | |
| 28 - H Cairoli | 0 | 101 | 1 | 108 | 0.35 | 0.01 | 8.77 | 1.00 | | | | |
| 2 - Centre for Disease Control | 6 | 27 | 0 | 15 | 9.37 | 0.49 | 179.00 | 1.17 | | | | |
| 46 - M Ceano-Vivas et al | 1 | 5 | 0 | 2 | 1.67 | 0.05 | 58.28 | 0.82 | | | | |
| 67 - G Fisler et al | 20 | 38 | 8 | 30 | 3.06 | 1.09 | 8.56 | 6.46 | | | | |
| 75 - V Giacomet et al | 2 | 15 | 1 | 20 | 2.92 | 0.24 | 35.68 | 1.58 | | | | |
| 79 - F Gotzinger et al | 15 | 196 | 2 | 62 | 2.49 | 0.55 | 11.19 | 3.77 | | | | |
| 91 - M Kainth et al | 6 | 31 | 2 | 8 | 0.72 | 0.12 | 4.50 | 2.74 | | | | |
| 99 - A Kambhampati | 41 | 123 | 9 | 24 | 0.83 | 0.34 | 2.06 | 7.52 | | | | |

| Name | Events 1 | Total 1 | Events 2 | Total 2 | Effect Estimate | CI Start | CI End | Weight | P(Q) | I²(Q) | Z | P(Z) |
|---|---|---|---|---|---|---|---|---|---|---|---|---|
| PIMS-TS/MIS-C | 373 | 566 | 247 | 556 | 2.65 | 1.48 | 4.74 | 31.75 | 0.25 | 21.32 | 3.27 | 0.00 |
| 10 - Z Almoussa et al | 3 | 4 | 2 | 2 | 0.47 | 0.01 | 16.89 | 0.81 | | | | |
| 12 - OY Antunez-Montes et al | 10 | 18 | 11 | 31 | 2.27 | 0.69 | 7.44 | 5.36 | | | | |
| 13 - AR Araujo da Silva et al | 1 | 1 | 3 | 4 | 1.29 | 0.03 | 53.51 | 0.75 | | | | |
| 143 - M Riollano-Cruz et al | 0 | 7 | 0 | 1 | 0.00 | 0.00 | 0.00 | 0.00 | | | | |
| 147 - L Shahbaznejad et al | 2 | 2 | 4 | 4 | 0.00 | 0.00 | 0.00 | 0.00 | | | | |
| 183 - Rekhtman et al | 8 | 10 | 1 | 2 | 4.00 | 0.17 | 95.76 | 1.02 | | | | |
| 187 - R Carbajal et al | 2 | 2 | 0 | 0 | 0.00 | 0.00 | 0.00 | 0.00 | | | | |
| 222 - Belay et al | 300 | 451 | 197 | 445 | 2.50 | 1.91 | 3.28 | 15.60 | | | | |
| 225 - Alkan et al | 4 | 13 | 0 | 9 | 9.00 | 0.42 | 191.37 | 1.09 | | | | |
| 38 - BC Clark et al | 0 | 11 | 6 | 17 | 0.08 | 0.00 | 1.53 | 1.14 | | | | |
| 5 - N Abdel-Haq et al | 5 | 5 | 4 | 10 | 15.89 | 0.69 | 365.14 | 1.04 | | | | |
| 60 - EM Dufort et al | 38 | 42 | 19 | 31 | 6.00 | 1.70 | 21.12 | 4.93 | | | | |
| Death | 179 | 3001 | 216 | 4620 | 1.38 | 0.97 | 1.96 | 100.00 | 0.25 | 18.44 | 1.81 | 0.07 |
| COVID-19 | 171 | 2458 | 207 | 4096 | 1.40 | 0.88 | 2.21 | 87.71 | 0.11 | 40.48 | 1.43 | 0.15 |
| 12 - OY Antunez-Montes et al | 0 | 80 | 0 | 153 | 0.00 | 0.00 | 0.00 | 0.00 | | | | |
| 13 - AR Araujo da Silva et al | 0 | 8 | 0 | 13 | 0.00 | 0.00 | 0.00 | 0.00 | | | | |
| 142 - S Richardson et al | 0 | 11 | 1 | 6 | 0.16 | 0.01 | 4.58 | 1.07 | | | | |
| 151 - B Sousa et al | 82 | 859 | 67 | 1499 | 2.26 | 1.61 | 3.15 | 36.84 | | | | |
| 155 - Kyung Sung | 0 | 18 | 0 | 8 | 0.00 | 0.00 | 0.00 | 0.00 | | | | |
| 157 - O Swann et al | 0 | 58 | 0 | 89 | 0.00 | 0.00 | 0.00 | 0.00 | | | | |
| 15 - JP Armann et al | 0 | 10 | 1 | 28 | 0.87 | 0.03 | 23.17 | 1.12 | | | | |
| 227 - Sena et al | 2 | 38 | 6 | 67 | 0.56 | 0.11 | 2.95 | 4.17 | | | | |
| 26 - CE Bolaños-Almeida et al | 0 | 148 | 2 | 103 | 0.14 | 0.01 | 2.88 | 1.30 | | | | |
| 28 - H Cairoli | 1 | 101 | 1 | 108 | 1.07 | 0.07 | 17.34 | 1.54 | | | | |
| 46 - M Ceano-Vivas et al | 0 | 5 | 0 | 2 | 0.00 | 0.00 | 0.00 | 0.00 | | | | |
| 58 - H Du et al | 0 | 30 | 0 | 58 | 0.00 | 0.00 | 0.00 | 0.00 | | | | |
| 75 - V Giacomet et al | 0 | 15 | 0 | 20 | 0.00 | 0.00 | 0.00 | 0.00 | | | | |
| 86 - D Hillesheim et al | 85 | 954 | 129 | 1918 | 1.36 | 1.02 | 1.80 | 40.50 | | | | |
| 99 - A Kambhampati | 1 | 123 | 0 | 24 | 0.60 | 0.02 | 15.17 | 1.16 | | | | |

| Name | Events 1 | Total 1 | Events 2 | Total 2 | Effect Estimate | CI Start | CI End | Weight | P(Q) | I²(Q) | Z | P(Z) |
|---|---|---|---|---|---|---|---|---|---|---|---|---|
| PIMS-TS/MIS-C | 8 | 543 | 9 | 524 | 0.82 | 0.31 | 2.13 | 12.29 | 0.80 | 0.00 | 0.41 | 0.68 |
| 10 - Z Almoussa et al | 1 | 4 | 0 | 2 | 2.14 | 0.06 | 77.54 | 0.94 | | | | |
| 12 - OY Antunez-Montes et al | 0 | 18 | 2 | 31 | 0.32 | 0.01 | 7.02 | 1.26 | | | | |
| 13 - AR Araujo da Silva et al | 0 | 1 | 0 | 4 | 0.00 | 0.00 | 0.00 | 0.00 | | | | |
| 143 - M Riollano-Cruz et al | 0 | 7 | 0 | 1 | 0.00 | 0.00 | 0.00 | 0.00 | | | | |
| 147 - L Shahbaznejad et al | 1 | 2 | 0 | 4 | 9.00 | 0.22 | 362.48 | 0.89 | | | | |
| 183 - Rekhtman et al | 1 | 10 | 0 | 2 | 0.79 | 0.02 | 25.90 | 0.99 | | | | |
| 187 - R Carbajal et al | 0 | 2 | 0 | 0 | 0.00 | 0.00 | 0.00 | 0.00 | | | | |
| 222 - Belay et al | 4 | 451 | 6 | 445 | 0.65 | 0.18 | 2.34 | 6.70 | | | | |
| 60 - EM Dufort et al | 1 | 42 | 1 | 31 | 0.73 | 0.04 | 12.17 | 1.52 | | | | |
| 89 - S Jain et al | 0 | 6 | 0 | 4 | 0.00 | 0.00 | 0.00 | 0.00 | | | | |
| Age >= 15 years | | | | | | | | | | | | |
| Invasive Ventilation | 190 | 2413 | 164 | 2222 | 1.20 | 0.95 | 1.53 | 100.00 | 0.67 | 0.00 | 1.54 | 0.12 |
| COVID-19 | 185 | 2379 | 161 | 2184 | 1.19 | 0.94 | 1.52 | 97.41 | 0.49 | 0.00 | 1.46 | 0.14 |
| 13 - AR Araujo da Silva et al | 0 | 6 | 1 | 13 | 0.64 | 0.02 | 18.05 | 0.50 | | | | |
| 142 - S Richardson et al | 3 | 20 | 1 | 6 | 0.88 | 0.07 | 10.46 | 0.91 | | | | |
| 151 - B Sousa et al | 85 | 757 | 119 | 1499 | 1.47 | 1.09 | 1.97 | 64.95 | | | | |
| 155 - Kyung Sung | 0 | 62 | 0 | 8 | 0.00 | 0.00 | 0.00 | 0.00 | | | | |
| 15 - JP Armann et al | 1 | 16 | 3 | 28 | 0.56 | 0.05 | 5.84 | 1.01 | | | | |
| 163 - S Verma et al | 3 | 20 | 1 | 9 | 1.41 | 0.13 | 15.78 | 0.96 | | | | |
| 21 - D Bayesheva et al | 1 | 86 | 0 | 137 | 4.82 | 0.19 | 119.78 | 0.54 | | | | |
| 221 - Preston et al | 87 | 1279 | 22 | 245 | 0.74 | 0.45 | 1.21 | 23.34 | | | | |
| 28 - H Cairoli | 0 | 72 | 1 | 108 | 0.49 | 0.02 | 12.30 | 0.54 | | | | |
| 46 - M Ceano-Vivas et al | 0 | 6 | 0 | 2 | 0.00 | 0.00 | 0.00 | 0.00 | | | | |
| 54 - A Desai et al | 5 | 45 | 13 | 109 | 0.92 | 0.31 | 2.76 | 4.65 | | | | |
| 75 - V Giacomet et al | 0 | 10 | 0 | 20 | 0.00 | 0.00 | 0.00 | 0.00 | | | | |
| PIMS-TS/MIS-C | 5 | 34 | 3 | 38 | 1.63 | 0.37 | 7.06 | 2.59 | 0.89 | 0.00 | 0.65 | 0.52 |
| 13 - AR Araujo da Silva et al | 0 | 1 | 0 | 4 | 0.00 | 0.00 | 0.00 | 0.00 | | | | |
| 143 - M Riollano-Cruz et al | 1 | 4 | 0 | 1 | 1.29 | 0.03 | 53.51 | 0.40 | | | | |
| 183 - Rekhtman et al | 0 | 1 | 0 | 2 | 0.00 | 0.00 | 0.00 | 0.00 | | | | |

| Name | Events 1 | Total 1 | Events 2 | Total 2 | Effect Estimate | CI Start | CI End | Weight | P(Q) | I²(Q) | Z | P(Z) |
|---|---|---|---|---|---|---|---|---|---|---|---|---|
| 41 - RL Crawford et al | 0 | 2 | 0 | 0 | 0.00 | 0.00 | 0.00 | 0.00 | | | | |
| 60 - EM Dufort et al | 4 | 26 | 3 | 31 | 1.70 | 0.34 | 8.39 | 2.19 | | | | |
| Cardiovascular Support | 19 | 153 | 20 | 193 | 0.95 | 0.21 | 4.34 | 100.00 | 0.23 | 31.54 | 0.07 | 0.94 |
| COVID-19 | 0 | 120 | 5 | 157 | 0.27 | 0.03 | 2.45 | 38.01 | 0.62 | 0.00 | 1.16 | 0.25 |
| 13 - AR Araujo da Silva et al | 0 | 6 | 0 | 13 | 0.00 | 0.00 | 0.00 | 0.00 | | | | |
| 142 - S Richardson et al | 0 | 20 | 0 | 6 | 0.00 | 0.00 | 0.00 | 0.00 | | | | |
| 15 - JP Armann et al | 0 | 16 | 4 | 28 | 0.16 | 0.01 | 3.27 | 20.10 | | | | |
| 28 - H Cairoli | 0 | 72 | 1 | 108 | 0.49 | 0.02 | 12.30 | 17.91 | | | | |
| 46 - M Ceano-Vivas et al | 0 | 6 | 0 | 2 | 0.00 | 0.00 | 0.00 | 0.00 | | | | |
| PIMS-TS/MIS-C | 19 | 33 | 15 | 36 | 2.01 | 0.69 | 5.89 | 61.99 | 1.00 | 0.00 | 1.28 | 0.20 |
| 13 - AR Araujo da Silva et al | 0 | 1 | 0 | 4 | 0.00 | 0.00 | 0.00 | 0.00 | | | | |
| 143 - M Riollano-Cruz et al | 0 | 4 | 0 | 1 | 0.00 | 0.00 | 0.00 | 0.00 | | | | |
| 41 - RL Crawford et al | 2 | 2 | 0 | 0 | 0.00 | 0.00 | 0.00 | 0.00 | | | | |
| 60 - EM Dufort et al | 17 | 26 | 15 | 31 | 2.01 | 0.69 | 5.89 | 61.99 | | | | |
| Admission to Critical Care | 1232 | 4793 | 982 | 4038 | 1.35 | 0.95 | 1.93 | 100.00 | 0.00 | 74.33 | 1.66 | 0.10 |
| COVID-19 | 866 | 4259 | 581 | 3165 | 0.98 | 0.69 | 1.40 | 69.19 | 0.00 | 58.83 | 0.09 | 0.92 |
| 122 - A Moreira et al | 236 | 1450 | 94 | 789 | 1.44 | 1.11 | 1.86 | 11.80 | | | | |
| 13 - AR Araujo da Silva et al | 2 | 6 | 6 | 13 | 0.58 | 0.08 | 4.39 | 2.49 | | | | |
| 142 - S Richardson et al | 9 | 20 | 3 | 6 | 0.82 | 0.13 | 5.08 | 2.91 | | | | |
| 151 - B Sousa et al | 197 | 757 | 342 | 1499 | 1.19 | 0.97 | 1.46 | 12.09 | | | | |
| 155 - Kyung Sung | 0 | 62 | 0 | 8 | 0.00 | 0.00 | 0.00 | 0.00 | | | | |
| 157 - O Swann et al | 19 | 109 | 15 | 89 | 1.04 | 0.50 | 2.19 | 8.11 | | | | |
| 15 - JP Armann et al | 3 | 16 | 4 | 28 | 1.38 | 0.27 | 7.15 | 3.41 | | | | |
| 163 - S Verma et al | 9 | 20 | 1 | 9 | 6.55 | 0.68 | 62.59 | 2.07 | | | | |
| 19 - S Bellino et al | 4 | 137 | 8 | 84 | 0.29 | 0.08 | 0.98 | 5.00 | | | | |
| 21 - D Bayesheva et al | 0 | 86 | 1 | 137 | 0.53 | 0.02 | 13.06 | 1.11 | | | | |
| 221 - Preston et al | 373 | 1279 | 103 | 245 | 0.57 | 0.43 | 0.75 | 11.66 | | | | |
| 224 - Graff et al | 8 | 23 | 0 | 10 | 11.52 | 0.60 | 221.75 | 1.29 | | | | |
| 26 - CE Bolaños-Almeida et al | 3 | 176 | 2 | 103 | 0.88 | 0.14 | 5.33 | 2.96 | | | | |
| 28 - H Cairoli | 0 | 72 | 1 | 108 | 0.49 | 0.02 | 12.30 | 1.11 | | | | |

| Name | Events 1 | Total 1 | Events 2 | Total 2 | Effect Estimate | CI Start | CI End | Weight | P(Q) | I²(Q) | Z | P(Z) |
|---|---|---|---|---|---|---|---|---|---|---|---|---|
| 2 - Centre for Disease Control | 2 | 30 | 0 | 15 | 2.72 | 0.12 | 60.29 | 1.19 | | | | |
| 46 - M Ceano-Vivas et al | 1 | 6 | 0 | 2 | 1.36 | 0.04 | 46.65 | 0.94 | | | | |
| 75 - V Giacomet et al | 0 | 10 | 1 | 20 | 0.62 | 0.02 | 16.57 | 1.07 | | | | |
| PIMS-TS/MIS-C | 366 | 534 | 401 | 873 | 2.56 | 2.03 | 3.21 | 30.81 | 0.96 | 0.00 | 8.06 | 0.00 |
| 13 - AR Araujo da Silva et al | 1 | 1 | 3 | 4 | 1.29 | 0.03 | 53.51 | 0.85 | | | | |
| 143 - M Riollano-Cruz et al | 0 | 4 | 0 | 1 | 0.00 | 0.00 | 0.00 | 0.00 | | | | |
| 179 - J Pang et al | 1 | 1 | 0 | 0 | 0.00 | 0.00 | 0.00 | 0.00 | | | | |
| 183 - Rekhtman et al | 1 | 1 | 1 | 2 | 3.00 | 0.06 | 151.19 | 0.77 | | | | |
| 222 - Belay et al | 161 | 246 | 197 | 445 | 2.38 | 1.73 | 3.29 | 11.40 | | | | |
| 226 - Abrams et al | 175 | 249 | 175 | 373 | 2.68 | 1.91 | 3.76 | 11.28 | | | | |
| 38 - BC Clark et al | 3 | 4 | 6 | 17 | 5.50 | 0.46 | 65.16 | 1.77 | | | | |
| 41 - RL Crawford et al | 2 | 2 | 0 | 0 | 0.00 | 0.00 | 0.00 | 0.00 | | | | |
| 60 - EM Dufort et al | 22 | 26 | 19 | 31 | 3.47 | 0.96 | 12.59 | 4.74 | | | | |
| Death | 311 | 4732 | 227 | 8286 | 1.81 | 1.00 | 3.24 | 100.00 | 0.00 | 74.82 | 1.98 | 0.05 |
| COVID-19 | 300 | 4453 | 220 | 7803 | 1.69 | 0.87 | 3.27 | 84.12 | 0.00 | 78.95 | 1.55 | 0.12 |
| 122 - A Moreira et al | 25 | 1450 | 13 | 789 | 1.05 | 0.53 | 2.06 | 16.56 | | | | |
| 13 - AR Araujo da Silva et al | 0 | 6 | 0 | 13 | 0.00 | 0.00 | 0.00 | 0.00 | | | | |
| 142 - S Richardson et al | 0 | 20 | 1 | 6 | 0.09 | 0.00 | 2.51 | 2.69 | | | | |
| 151 - B Sousa et al | 74 | 757 | 67 | 1499 | 2.32 | 1.64 | 3.26 | 19.80 | | | | |
| 155 - Kyung Sung | 0 | 62 | 0 | 8 | 0.00 | 0.00 | 0.00 | 0.00 | | | | |
| 157 - O Swann et al | 3 | 109 | 0 | 89 | 5.88 | 0.30 | 115.41 | 3.28 | | | | |
| 15 - JP Armann et al | 0 | 16 | 1 | 28 | 0.56 | 0.02 | 14.45 | 2.81 | | | | |
| 227 - Sena et al | 10 | 66 | 6 | 67 | 1.82 | 0.62 | 5.32 | 12.39 | | | | |
| 26 - CE Bolaños-Almeida et al | 0 | 176 | 2 | 103 | 0.12 | 0.01 | 2.42 | 3.15 | | | | |
| 28 - H Cairoli | 0 | 72 | 1 | 108 | 0.49 | 0.02 | 12.30 | 2.87 | | | | |
| 46 - M Ceano-Vivas et al | 0 | 6 | 0 | 2 | 0.00 | 0.00 | 0.00 | 0.00 | | | | |
| 75 - V Giacomet et al | 0 | 10 | 0 | 20 | 0.00 | 0.00 | 0.00 | 0.00 | | | | |
| 86 - D Hillesheim et al | 188 | 1703 | 129 | 5071 | 4.75 | 3.77 | 5.99 | 20.56 | | | | |
| PIMS-TS/MIS-C | 11 | 279 | 7 | 483 | 2.01 | 0.31 | 12.87 | 15.88 | 0.20 | 38.13 | 0.74 | 0.46 |
| 13 - AR Araujo da Silva et al | 0 | 1 | 0 | 4 | 0.00 | 0.00 | 0.00 | 0.00 | | | | |

| Name | Events 1 | Total 1 | Events 2 | Total 2 | Effect Estimate | CI Start | CI End | Weight | P(Q) | I²(Q) | Z | P(Z) |
|---|---|---|---|---|---|---|---|---|---|---|---|---|
| 143 - M Riollano-Cruz et al | 0 | 4 | 0 | 1 | 0.00 | 0.00 | 0.00 | 0.00 | | | | |
| 183 - Rekhtman et al | 0 | 1 | 0 | 2 | 0.00 | 0.00 | 0.00 | 0.00 | | | | |
| 222 - Belay et al | 11 | 245 | 6 | 445 | 3.44 | 1.26 | 9.42 | 13.05 | | | | |
| 41 - RL Crawford et al | 0 | 2 | 0 | 0 | 0.00 | 0.00 | 0.00 | 0.00 | | | | |
| 60 - EM Dufort et al | 0 | 26 | 1 | 31 | 0.38 | 0.01 | 9.82 | 2.83 | | | | |
| Association between asian race and severe outcomes | | | | | | | | | | | | |
| Invasive Ventilation | 18 | 134 | 1395 | 13687 | 1.09 | 0.63 | 1.88 | 100.00 | 0.68 | 0.00 | 0.30 | 0.77 |
| COVID-19 | 12 | 120 | 1372 | 13633 | 1.20 | 0.65 | 2.21 | 80.66 | 0.58 | 0.00 | 0.58 | 0.56 |
| 142 - S Richardson et al | 3 | 12 | 10 | 95 | 2.83 | 0.66 | 12.22 | 14.19 | | | | |
| 151 - B Sousa et al | 5 | 45 | 750 | 6891 | 1.02 | 0.40 | 2.60 | 34.85 | | | | |
| 183 - Rekhtman et al | 0 | 0 | 5 | 19 | 0.00 | 0.00 | 0.00 | 0.00 | | | | |
| 75 - V Giacomet et al | 0 | 15 | 1 | 112 | 2.40 | 0.09 | 61.50 | 2.88 | | | | |
| 86 - D Hillesheim et al | 4 | 48 | 606 | 6516 | 0.89 | 0.32 | 2.48 | 28.75 | | | | |
| PIMS-TS/MIS-C | 6 | 14 | 23 | 54 | 0.72 | 0.21 | 2.52 | 19.34 | 0.41 | 0.00 | 0.51 | 0.61 |
| 167 - E Whittaker et al | 4 | 12 | 21 | 46 | 0.60 | 0.16 | 2.26 | 17.06 | | | | |
| 179 - J Pang et al | 2 | 2 | 2 | 3 | 3.00 | 0.08 | 115.34 | 2.28 | | | | |
| 41 - RL Crawford et al | 0 | 0 | 0 | 5 | 0.00 | 0.00 | 0.00 | 0.00 | | | | |
| CV support | 6 | 12 | 26 | 84 | 1.19 | 0.33 | 4.25 | 100.00 | 1.00 | 0.00 | 0.27 | 0.79 |
| COVID-19 | 0 | 0 | 1 | 33 | 0.00 | 0.00 | 0.00 | 0.00 | 0.00 | 0.00 | 0.00 | 0.00 |
| 46 - M Ceano-Vivas et al | 0 | 0 | 1 | 33 | 0.00 | 0.00 | 0.00 | 0.00 | | | | |
| PIMS-TS/MIS-C | 6 | 12 | 25 | 51 | 1.19 | 0.33 | 4.25 | 100.00 | 1.00 | 0.00 | 0.27 | 0.79 |
| 167 - E Whittaker et al | 6 | 12 | 21 | 46 | 1.19 | 0.33 | 4.25 | 100.00 | | | | |
| 41 - RL Crawford et al | 0 | 0 | 4 | 5 | 0.00 | 0.00 | 0.00 | 0.00 | | | | |
| Admission to critical care | 68 | 234 | 2878 | 10652 | 1.25 | 0.92 | 1.69 | 100.00 | 0.76 | 0.00 | 1.40 | 0.16 |
| COVID-19 | 56 | 210 | 2821 | 10543 | 1.27 | 0.92 | 1.76 | 88.51 | 0.57 | 0.00 | 1.43 | 0.15 |
| 122 - A Moreira et al | 4 | 15 | 101 | 430 | 1.18 | 0.37 | 3.80 | 6.95 | | | | |
| 142 - S Richardson et al | 6 | 12 | 30 | 95 | 2.17 | 0.65 | 7.28 | 6.44 | | | | |
| 151 - B Sousa et al | 9 | 45 | 1859 | 6891 | 0.68 | 0.33 | 1.41 | 17.63 | | | | |
| 157 - O Swann et al | 10 | 58 | 68 | 522 | 1.39 | 0.67 | 2.88 | 17.87 | | | | |
| 221 - Preston et al | 21 | 56 | 726 | 2374 | 1.36 | 0.79 | 2.36 | 31.48 | | | | |

| Name | Events 1 | Total 1 | Events 2 | Total 2 | Effect Estimate | CI Start | CI End | Weight | P(Q) | I²(Q) | Z | P(Z) |
|---|---|---|---|---|---|---|---|---|---|---|---|---|
| 46 - M Ceano-Vivas et al | 0 | 0 | 5 | 33 | 0.00 | 0.00 | 0.00 | 0.00 | | | | |
| 67 - G Fisler et al | 4 | 9 | 26 | 86 | 1.85 | 0.46 | 7.43 | 4.87 | | | | |
| 75 - V Giacomet et al | 2 | 15 | 6 | 112 | 2.72 | 0.50 | 14.89 | 3.27 | | | | |
| PIMS-TS/MIS-C | 12 | 24 | 57 | 109 | 1.08 | 0.44 | 2.67 | 11.49 | 0.87 | 0.00 | 0.16 | 0.87 |
| 167 - E Whittaker et al | 6 | 12 | 23 | 46 | 1.00 | 0.28 | 3.56 | 5.86 | | | | |
| 183 - Rekhtman et al | 0 | 0 | 12 | 19 | 0.00 | 0.00 | 0.00 | 0.00 | | | | |
| 38 - BC Clark et al | 6 | 12 | 18 | 39 | 1.17 | 0.32 | 4.26 | 5.64 | | | | |
| 41 - RL Crawford et al | 0 | 0 | 4 | 5 | 0.00 | 0.00 | 0.00 | 0.00 | | | | |
| Death | 6 | 86 | 577 | 7605 | 1.85 | 0.88 | 3.90 | 100.00 | 0.97 | 0.00 | 1.63 | 0.10 |
| COVID-19 | 6 | 73 | 574 | 7526 | 1.84 | 0.84 | 4.01 | 90.53 | 0.88 | 0.00 | 1.52 | 0.13 |
| 122 - A Moreira et al | 0 | 15 | 12 | 430 | 1.08 | 0.06 | 19.08 | 6.72 | | | | |
| 13 - AR Araujo da Silva et al | 0 | 0 | 1 | 46 | 0.00 | 0.00 | 0.00 | 0.00 | | | | |
| 142 - S Richardson et al | 0 | 12 | 1 | 95 | 2.52 | 0.10 | 65.29 | 5.23 | | | | |
| 151 - B Sousa et al | 6 | 45 | 558 | 6891 | 1.75 | 0.74 | 4.14 | 74.29 | | | | |
| 46 - M Ceano-Vivas et al | 0 | 0 | 1 | 33 | 0.00 | 0.00 | 0.00 | 0.00 | | | | |
| 92 - R Marcello et al | 0 | 1 | 1 | 31 | 6.78 | 0.19 | 247.05 | 4.29 | | | | |
| PIMS-TS/MIS-C | 0 | 13 | 3 | 79 | 2.05 | 0.18 | 23.03 | 9.47 | 0.64 | 0.00 | 0.58 | 0.56 |
| 143 - M Riollano-Cruz et al | 0 | 0 | 1 | 15 | 0.00 | 0.00 | 0.00 | 0.00 | | | | |
| 167 - E Whittaker et al | 0 | 12 | 1 | 46 | 1.21 | 0.05 | 31.65 | 5.21 | | | | |
| 183 - Rekhtman et al | 0 | 1 | 1 | 18 | 3.89 | 0.11 | 143.60 | 4.26 | | | | |
| Association between black race and severe outcomes | | | | | | | | | | | | |
| Invasive Ventilation | 123 | 1028 | 1499 | 15538 | 1.38 | 1.11 | 1.70 | 100.00 | 0.58 | 0.00 | 2.93 | 0.00 |
| COVID-19 | 107 | 995 | 1474 | 15423 | 1.32 | 1.05 | 1.67 | 94.28 | 0.39 | 4.52 | 2.36 | 0.02 |
| 13 - AR Araujo da Silva et al | 0 | 1 | 2 | 14 | 1.67 | 0.05 | 53.92 | 0.38 | | | | |
| 142 - S Richardson et al | 3 | 27 | 10 | 80 | 0.88 | 0.22 | 3.45 | 2.42 | | | | |
| 151 - B Sousa et al | 28 | 240 | 727 | 6696 | 1.08 | 0.73 | 1.62 | 28.27 | | | | |
| 221 - Preston et al | 50 | 478 | 122 | 1952 | 1.75 | 1.24 | 2.48 | 38.17 | | | | |
| 54 - A Desai et al | 8 | 54 | 20 | 185 | 1.43 | 0.59 | 3.47 | 5.85 | | | | |
| 75 - V Giacomet et al | 0 | 4 | 1 | 123 | 9.07 | 0.32 | 255.04 | 0.41 | | | | |
| 86 - D Hillesheim et al | 18 | 191 | 592 | 6373 | 1.02 | 0.62 | 1.66 | 18.77 | | | | |

| Name | Events 1 | Total 1 | Events 2 | Total 2 | Effect Estimate | CI Start | CI End | Weight | P(Q) | I²(Q) | Z | P(Z) |
|---|---|---|---|---|---|---|---|---|---|---|---|---|
| PIMS-TS/MIS-C | 16 | 33 | 25 | 115 | 2.39 | 0.98 | 5.84 | 5.72 | 0.80 | 0.00 | 1.92 | 0.06 |
| 13 - AR Araujo da Silva et al | 0 | 1 | 4 | 45 | 3.07 | 0.11 | 87.20 | 0.41 | | | | |
| 143 - M Riollano-Cruz et al | 1 | 2 | 2 | 13 | 5.50 | 0.23 | 128.97 | 0.46 | | | | |
| 167 - E Whittaker et al | 13 | 22 | 12 | 36 | 2.89 | 0.96 | 8.65 | 3.79 | | | | |
| 179 - J Pang et al | 1 | 1 | 3 | 4 | 1.29 | 0.03 | 53.51 | 0.33 | | | | |
| 183 - Rekhtman et al | 1 | 5 | 4 | 14 | 0.63 | 0.05 | 7.46 | 0.74 | | | | |
| 41 - RL Crawford et al | 0 | 2 | 0 | 3 | 0.00 | 0.00 | 0.00 | 0.00 | | | | |
| CV support | 16 | 24 | 16 | 72 | 3.09 | 1.07 | 8.88 | 100.00 | 0.99 | 0.00 | 2.09 | 0.04 |
| COVID-19 | 0 | 0 | 1 | 33 | 0.00 | 0.00 | 0.00 | 0.00 | 0.00 | 0.00 | 0.00 | 0.00 |
| 46 - M Ceano-Vivas et al | 0 | 0 | 1 | 33 | 0.00 | 0.00 | 0.00 | 0.00 | | | | |
| PIMS-TS/MIS-C | 16 | 24 | 15 | 39 | 3.09 | 1.07 | 8.88 | 100.00 | 0.99 | 0.00 | 2.09 | 0.04 |
| 167 - E Whittaker et al | 14 | 22 | 13 | 36 | 3.10 | 1.03 | 9.33 | 91.63 | | | | |
| 41 - RL Crawford et al | 2 | 2 | 2 | 3 | 3.00 | 0.08 | 115.34 | 8.37 | | | | |
| Admission to Critical Care | 352 | 1010 | 2676 | 10082 | 1.61 | 1.16 | 2.23 | 100.00 | 0.01 | 50.51 | 2.87 | 0.00 |
| COVID-19 | 305 | 940 | 2621 | 9971 | 1.51 | 1.06 | 2.15 | 82.10 | 0.01 | 59.32 | 2.28 | 0.02 |
| 122 - A Moreira et al | 30 | 114 | 75 | 331 | 1.22 | 0.75 | 1.99 | 14.05 | | | | |
| 13 - AR Araujo da Silva et al | 1 | 1 | 25 | 45 | 2.41 | 0.09 | 62.39 | 0.95 | | | | |
| 142 - S Richardson et al | 12 | 27 | 24 | 80 | 1.87 | 0.76 | 4.58 | 8.03 | | | | |
| 151 - B Sousa et al | 53 | 240 | 1814 | 6696 | 0.76 | 0.56 | 1.04 | 17.36 | | | | |
| 157 - O Swann et al | 12 | 46 | 66 | 534 | 2.50 | 1.23 | 5.07 | 10.45 | | | | |
| 221 - Preston et al | 182 | 478 | 565 | 1952 | 1.51 | 1.22 | 1.86 | 18.99 | | | | |
| 224 - Graff et al | 1 | 3 | 10 | 63 | 2.65 | 0.22 | 32.08 | 1.57 | | | | |
| 32 - J Chao et al | 2 | 5 | 11 | 41 | 1.82 | 0.27 | 12.38 | 2.53 | | | | |
| 46 - M Ceano-Vivas et al | 0 | 0 | 5 | 33 | 0.00 | 0.00 | 0.00 | 0.00 | | | | |
| 67 - G Fisler et al | 12 | 22 | 18 | 73 | 3.67 | 1.36 | 9.90 | 7.05 | | | | |
| 75 - V Giacomet et al | 0 | 4 | 8 | 123 | 1.51 | 0.07 | 30.44 | 1.11 | | | | |
| PIMS-TS/MIS-C | 47 | 70 | 55 | 111 | 2.13 | 0.96 | 4.75 | 17.90 | 0.29 | 18.72 | 1.85 | 0.06 |
| 13 - AR Araujo da Silva et al | 1 | 1 | 11 | 14 | 0.91 | 0.03 | 27.83 | 0.87 | | | | |
| 167 - E Whittaker et al | 13 | 22 | 16 | 36 | 1.81 | 0.62 | 5.29 | 6.35 | | | | |
| 183 - Rekhtman et al | 2 | 5 | 10 | 14 | 0.27 | 0.03 | 2.25 | 2.09 | | | | |

| Name | Events 1 | Total 1 | Events 2 | Total 2 | Effect Estimate | CI Start | CI End | Weight | P(Q) | I²(Q) | Z | P(Z) |
|---|---|---|---|---|---|---|---|---|---|---|---|---|
| 38 - BC Clark et al | 11 | 15 | 13 | 36 | 4.87 | 1.29 | 18.42 | 4.63 | | | | |
| 41 - RL Crawford et al | 2 | 2 | 2 | 3 | 3.00 | 0.08 | 115.34 | 0.77 | | | | |
| 5 - N Abdel-Haq et al | 18 | 25 | 3 | 8 | 4.29 | 0.80 | 22.92 | 3.19 | | | | |
| Death | 25 | 415 | 558 | 7276 | 1.50 | 0.79 | 2.83 | 100.00 | 0.34 | 12.18 | 1.25 | 0.21 |
| COVID-19 | 24 | 386 | 556 | 7213 | 1.91 | 0.71 | 5.12 | 89.69 | 0.13 | 43.60 | 1.29 | 0.20 |
| 122 - A Moreira et al | 6 | 114 | 6 | 331 | 3.01 | 0.95 | 9.53 | 22.55 | | | | |
| 13 - AR Araujo da Silva et al | 0 | 1 | 1 | 45 | 9.89 | 0.27 | 358.47 | 3.01 | | | | |
| 142 - S Richardson et al | 1 | 27 | 0 | 80 | 9.11 | 0.36 | 230.51 | 3.69 | | | | |
| 151 - B Sousa et al | 17 | 240 | 547 | 6696 | 0.86 | 0.52 | 1.41 | 57.00 | | | | |
| 46 - M Ceano-Vivas et al | 0 | 0 | 1 | 33 | 0.00 | 0.00 | 0.00 | 0.00 | | | | |
| 92 - R Marcello et al | 0 | 4 | 1 | 28 | 2.04 | 0.07 | 58.19 | 3.44 | | | | |
| PIMS-TS/MIS-C | 1 | 29 | 2 | 63 | 1.96 | 0.28 | 13.56 | 10.31 | 0.74 | 0.00 | 0.68 | 0.50 |
| 143 - M Riollano-Cruz et al | 0 | 2 | 1 | 13 | 1.67 | 0.05 | 53.92 | 3.20 | | | | |
| 167 - E Whittaker et al | 1 | 22 | 0 | 36 | 5.09 | 0.20 | 130.65 | 3.66 | | | | |
| 183 - Rekhtman et al | 0 | 5 | 1 | 14 | 0.82 | 0.03 | 23.34 | 3.44 | | | | |
| Association between white race and severe outcome | | | | | | | | | | | | |
| Invasive Ventilation | 428 | 4873 | 1203 | 11796 | 0.85 | 0.67 | 1.07 | 100.00 | 0.12 | 33.25 | 1.42 | 0.16 |
| COVID-19 | 420 | 4843 | 1172 | 11710 | 0.85 | 0.71 | 1.03 | 94.91 | 0.22 | 26.21 | 1.67 | 0.09 |
| 13 - AR Araujo da Silva et al | 3 | 24 | 1 | 22 | 3.00 | 0.29 | 31.22 | 0.95 | | | | |
| 142 - S Richardson et al | 0 | 20 | 13 | 87 | 0.13 | 0.01 | 2.36 | 0.64 | | | | |
| 151 - B Sousa et al | 224 | 2110 | 531 | 4826 | 0.96 | 0.81 | 1.13 | 33.07 | | | | |
| 174 - P Zachariah et al | 5 | 27 | 4 | 23 | 1.08 | 0.25 | 4.61 | 2.38 | | | | |
| 221 - Preston et al | 38 | 639 | 134 | 1791 | 0.78 | 0.54 | 1.13 | 19.55 | | | | |
| 54 - A Desai et al | 12 | 109 | 16 | 184 | 1.30 | 0.59 | 2.86 | 7.03 | | | | |
| 75 - V Giacomet et al | 0 | 95 | 1 | 32 | 0.11 | 0.00 | 2.77 | 0.50 | | | | |
| 86 - D Hillesheim et al | 138 | 1819 | 472 | 4745 | 0.74 | 0.61 | 0.91 | 30.80 | | | | |
| PIMS-TS/MIS-C | 8 | 30 | 31 | 86 | 1.02 | 0.16 | 6.34 | 5.09 | 0.08 | 51.54 | 0.02 | 0.98 |
| 13 - AR Araujo da Silva et al | 2 | 5 | 0 | 9 | 13.57 | 0.51 | 358.64 | 0.49 | | | | |
| 143 - M Riollano-Cruz et al | 0 | 2 | 3 | 13 | 0.60 | 0.02 | 15.76 | 0.49 | | | | |
| 167 - E Whittaker et al | 5 | 18 | 20 | 40 | 0.38 | 0.12 | 1.28 | 3.36 | | | | |

| Name | Events 1 | Total 1 | Events 2 | Total 2 | Effect Estimate | CI Start | CI End | Weight | P(Q) | I²(Q) | Z | P(Z) |
|---|---|---|---|---|---|---|---|---|---|---|---|---|
| 179 - J Pang et al | 0 | 1 | 4 | 4 | 0.04 | 0.00 | 2.82 | 0.28 | | | | |
| 183 - Rekhtman et al | 1 | 1 | 4 | 18 | 9.67 | 0.33 | 281.33 | 0.46 | | | | |
| 41 - RL Crawford et al | 0 | 3 | 0 | 2 | 0.00 | 0.00 | 0.00 | 0.00 | | | | |
| CV support | 6 | 49 | 26 | 47 | 0.19 | 0.06 | 0.58 | 100.00 | 0.71 | 0.00 | 2.89 | 0.00 |
| COVID-19 | 0 | 28 | 1 | 5 | 0.05 | 0.00 | 1.50 | 11.44 | 0.00 | 100.00 | 1.72 | 0.09 |
| 46 - M Ceano-Vivas et al | 0 | 28 | 1 | 5 | 0.05 | 0.00 | 1.50 | 11.44 | | | | |
| PIMS-TS/MIS-C | 6 | 21 | 25 | 42 | 0.22 | 0.07 | 0.74 | 88.56 | 0.82 | 0.00 | 2.45 | 0.01 |
| 167 - E Whittaker et al | 4 | 18 | 23 | 40 | 0.21 | 0.06 | 0.76 | 78.92 | | | | |
| 41 - RL Crawford et al | 2 | 3 | 2 | 2 | 0.33 | 0.01 | 12.82 | 9.65 | | | | |
| Admission to Critical Care | 889 | 3408 | 2143 | 7683 | 0.63 | 0.46 | 0.87 | 100.00 | 0.00 | 65.88 | 2.81 | 0.00 |
| COVID-19 | 870 | 3356 | 2060 | 7555 | 0.72 | 0.53 | 1.00 | 85.80 | 0.00 | 70.05 | 1.96 | 0.05 |
| 122 - A Moreira et al | 17 | 102 | 88 | 343 | 0.58 | 0.33 | 1.03 | 11.81 | | | | |
| 13 - AR Araujo da Silva et al | 16 | 24 | 10 | 22 | 2.40 | 0.73 | 7.92 | 5.23 | | | | |
| 142 - S Richardson et al | 3 | 20 | 33 | 87 | 0.29 | 0.08 | 1.06 | 4.60 | | | | |
| 151 - B Sousa et al | 598 | 2110 | 1269 | 4826 | 1.11 | 0.99 | 1.24 | 18.58 | | | | |
| 157 - O Swann et al | 32 | 307 | 46 | 273 | 0.57 | 0.35 | 0.93 | 13.27 | | | | |
| 221 - Preston et al | 191 | 639 | 556 | 1791 | 0.95 | 0.78 | 1.15 | 17.76 | | | | |
| 224 - Graff et al | 1 | 13 | 10 | 53 | 0.36 | 0.04 | 3.09 | 1.98 | | | | |
| 32 - J Chao et al | 2 | 3 | 11 | 43 | 5.82 | 0.48 | 70.62 | 1.52 | | | | |
| 46 - M Ceano-Vivas et al | 3 | 28 | 2 | 5 | 0.18 | 0.02 | 1.55 | 1.98 | | | | |
| 67 - G Fisler et al | 3 | 15 | 27 | 80 | 0.49 | 0.13 | 1.89 | 4.36 | | | | |
| 75 - V Giacomet et al | 4 | 95 | 8 | 32 | 0.13 | 0.04 | 0.48 | 4.71 | | | | |
| PIMS-TS/MIS-C | 19 | 52 | 83 | 128 | 0.31 | 0.15 | 0.66 | 14.20 | 0.51 | 0.00 | 3.05 | 0.00 |
| 13 - AR Araujo da Silva et al | 4 | 5 | 8 | 9 | 0.50 | 0.02 | 10.25 | 1.06 | | | | |
| 167 - E Whittaker et al | 7 | 18 | 22 | 40 | 0.52 | 0.17 | 1.62 | 5.63 | | | | |
| 183 - Rekhtman et al | 1 | 1 | 11 | 18 | 1.96 | 0.07 | 54.67 | 0.88 | | | | |
| 38 - BC Clark et al | 5 | 22 | 19 | 29 | 0.15 | 0.04 | 0.54 | 4.84 | | | | |
| 41 - RL Crawford et al | 2 | 3 | 2 | 2 | 0.33 | 0.01 | 12.82 | 0.74 | | | | |
| 5 - N Abdel-Haq et al | 0 | 3 | 21 | 30 | 0.06 | 0.00 | 1.35 | 1.04 | | | | |
| Death | 131 | 2326 | 452 | 5370 | 0.65 | 0.54 | 0.80 | 100.00 | 0.84 | 0.00 | 4.16 | 0.00 |

| Name | Events 1 | Total 1 | Events 2 | Total 2 | Effect Estimate | CI Start | CI End | Weight | P(Q) | I²(Q) | Z | P(Z) |
|---|---|---|---|---|---|---|---|---|---|---|---|---|
| COVID-19 | 131 | 2302 | 449 | 5297 | 0.65 | 0.53 | 0.79 | 98.99 | 0.70 | 0.00 | 4.23 | 0.00 |
| 122 - A Moreira et al | 2 | 102 | 10 | 343 | 0.67 | 0.14 | 3.09 | 1.69 | | | | |
| 13 - AR Araujo da Silva et al | 0 | 24 | 1 | 22 | 0.29 | 0.01 | 7.56 | 0.38 | | | | |
| 142 - S Richardson et al | 0 | 20 | 1 | 87 | 1.41 | 0.06 | 35.79 | 0.38 | | | | |
| 151 - B Sousa et al | 129 | 2110 | 435 | 4826 | 0.66 | 0.54 | 0.81 | 95.83 | | | | |
| 46 - M Ceano-Vivas et al | 0 | 28 | 1 | 5 | 0.05 | 0.00 | 1.50 | 0.35 | | | | |
| 92 - R Marcello et al | 0 | 18 | 1 | 14 | 0.24 | 0.01 | 6.44 | 0.37 | | | | |
| PIMS-TS/MIS-C | 0 | 24 | 3 | 73 | 1.57 | 0.22 | 11.39 | 1.01 | 0.79 | 0.00 | 0.44 | 0.66 |
| 143 - M Riollano-Cruz et al | 0 | 2 | 1 | 13 | 1.67 | 0.05 | 53.92 | 0.33 | | | | |
| 167 - E Whittaker et al | 0 | 18 | 1 | 40 | 0.71 | 0.03 | 18.32 | 0.38 | | | | |
| 183 - Rekhtman et al | 0 | 1 | 1 | 18 | 3.89 | 0.11 | 143.60 | 0.30 | | | | |
| 41 - RL Crawford et al | 0 | 3 | 0 | 2 | 0.00 | 0.00 | 0.00 | 0.00 | | | | |
| Comparison between hispanic ethnicity and severe disease | | | | | | | | | | | | |
| Invasive Ventilation | 64 | 1001 | 141 | 1725 | 0.76 | 0.35 | 1.68 | 100.00 | 0.16 | 37.59 | 0.67 | 0.50 |
| COVID-19 | 62 | 985 | 135 | 1707 | 0.98 | 0.36 | 2.67 | 84.30 | 0.08 | 55.15 | 0.04 | 0.97 |
| 174 - P Zachariah et al | 4 | 25 | 5 | 25 | 0.76 | 0.18 | 3.25 | 18.39 | | | | |
| 221 - Preston et al | 55 | 936 | 117 | 1494 | 0.73 | 0.53 | 1.02 | 44.50 | | | | |
| 54 - A Desai et al | 2 | 17 | 13 | 68 | 0.56 | 0.11 | 2.78 | 16.28 | | | | |
| 75 - V Giacomet et al | 1 | 7 | 0 | 120 | 55.62 | 2.06 | 1502.58 | 5.14 | | | | |
| PIMS-TS/MIS-C | 2 | 16 | 6 | 18 | 0.29 | 0.05 | 1.79 | 15.70 | 0.59 | 0.00 | 1.33 | 0.18 |
| 143 - M Riollano-Cruz et al | 1 | 10 | 2 | 5 | 0.17 | 0.01 | 2.56 | 7.13 | | | | |
| 183 - Rekhtman et al | 1 | 6 | 4 | 13 | 0.45 | 0.04 | 5.21 | 8.57 | | | | |
| Cardiovascular Support | 0 | 2 | 0 | 31 | 0.00 | 0.00 | 0.00 | 0.00 | 0.00 | 0.00 | 0.00 | 0.00 |
| COVID-19 | 0 | 0 | 0 | 0 | 0.00 | 0.00 | 0.00 | 0.00 | 0.00 | 0.00 | 0.00 | 0.00 |
| PIMS-TS/MIS-C | 0 | 2 | 0 | 31 | 0.00 | 0.00 | 0.00 | 0.00 | 0.00 | 0.00 | 0.00 | 0.00 |
| 5 - N Abdel-Haq et al | 0 | 2 | 0 | 31 | 0.00 | 0.00 | 0.00 | 0.00 | | | | |
| Admission to critical care | 279 | 1038 | 560 | 1756 | 0.72 | 0.32 | 1.63 | 100.00 | 0.01 | 63.66 | 0.78 | 0.43 |
| COVID-19 | 276 | 1030 | 530 | 1712 | 0.88 | 0.35 | 2.23 | 84.15 | 0.01 | 71.81 | 0.26 | 0.79 |
| 221 - Preston et al | 255 | 936 | 492 | 1494 | 0.76 | 0.64 | 0.91 | 27.52 | | | | |
| 224 - Graff et al | 8 | 40 | 3 | 26 | 1.92 | 0.46 | 8.02 | 15.00 | | | | |

| Name | Events 1 | Total 1 | Events 2 | Total 2 | Effect Estimate | CI Start | CI End | Weight | P(Q) | I²(Q) | Z | P(Z) |
|---|---|---|---|---|---|---|---|---|---|---|---|---|
| 32 - J Chao et al | 3 | 29 | 10 | 17 | 0.08 | 0.02 | 0.38 | 14.01 | | | | |
| 67 - G Fisler et al | 9 | 18 | 18 | 55 | 2.06 | 0.70 | 6.07 | 18.70 | | | | |
| 75 - V Giacomet et al | 1 | 7 | 7 | 120 | 2.69 | 0.28 | 25.54 | 8.92 | | | | |
| 10PIMS-TS/MIS-C | 3 | 8 | 30 | 44 | 0.24 | 0.04 | 1.32 | 15.85 | 0.48 | 0.00 | 1.64 | 0.10 |
| 183 - Rekhtman et al | 2 | 6 | 10 | 13 | 0.15 | 0.02 | 1.26 | 9.59 | | | | |
| 5 - N Abdel-Haq et al | 1 | 2 | 20 | 31 | 0.55 | 0.03 | 9.68 | 6.26 | | | | |
| Death | 6 | 209 | 9 | 298 | 0.94 | 0.30 | 2.93 | 100.00 | 0.36 | 6.42 | 0.11 | 0.91 |
| COVID-19 | 5 | 193 | 8 | 280 | 0.88 | 0.28 | 2.74 | 78.06 | 0.46 | 0.00 | 0.22 | 0.82 |
| 122 - A Moreira et al | 4 | 176 | 8 | 265 | 0.75 | 0.22 | 2.52 | 66.49 | | | | |
| 92 - R Marcello et al | 1 | 17 | 0 | 15 | 2.82 | 0.11 | 74.51 | 11.58 | | | | |
| PIMS-TS/MIS-C | 1 | 16 | 1 | 18 | 1.03 | 0.02 | 49.24 | 21.94 | 0.10 | 62.00 | 0.02 | 0.99 |
| 143 - M Riollano-Cruz et al | 0 | 10 | 1 | 5 | 0.14 | 0.00 | 4.22 | 10.86 | | | | |
| 183 - Rekhtman et al | 1 | 6 | 0 | 13 | 7.36 | 0.26 | 210.02 | 11.08 | | | | |

**Acknowledgements**: Thanks to the following authors of studies who provided individual patient data or additional aggregated data for inclusion in the meta-analysis.

Ho Kyung Sung, Jin Yong Kim, Jeonghun Heo, Haesook Seo, Young soo Jan, Hyewon Kim, Bo Ram Koh, Neungsun Jo, Hong Sang Oh, Young Mi Baek, Kyung-Haw Park, Jeung A Shon, Min-Chul Kim, Joon Ho Kim, Hyun-Ha Chang, Yukyung Park, Yu Min Kang, Dong Hyun Lee, Dong Hyun Oh, Hyun Jung Park, Kyoung-Ho Song, Eun Kyoung Lee, Hyeongseok Jeong, Ji Yeon Lee, Ja-Young Ko, Johee Choi, Eun Hwa Ryu, Ki-hyun Chung, Myoung-don Oh, Dinagul Bayesheva , Riza Boranbayeva , Bayan Turdalina , Ildar Fakhradiyev, Timur Saliev , Shynar Tanabayeva , Baurzhan Zhussupov, Talgat Nurgozhin, S Jain, S Sen, S Lakshmivenkateshiah, P Bobhate, S Venkatesh, S Udani, L Shobhavat, P Andankar, T Karande, S Kulkarni, Jakob Peter Armann, Natalie Diffloth, Arne Simon, Maren Doenhardt, Markus Hufnagel, Andreas Trotter, Dominik Schneider, Johannes Hübner, Reinhard Berner, Vania Giacomet, Lucia Barcellini, Marta Stracuzzi, Emma Longoni, Laura Folgori, Alessandro Leone, Gian Vincenzo Zuccotti, María de Ceano-Vivas, Irene Martín-Espín,Teresa del Rosal, Marta Bueno-Barriocanal, Marta Plata-Gallardo, José Antonio Ruiz-Domínguez, Rosario López-López, Miguel Ángel Molina-Gutiérrez, Patricia Bote-Gascón,, Isabel González-Bertolín, Paula García-Sánchez, Julia Martín-Sánchez, Begoña de Miguel-Lavisier,Talía Sainz,Fernando Baquero-Artigao, Ana Méndez-Echevarría, Cristina Calvo2, Jasmin Pfefferle, Angela Zacharasiewicz, Matthias Bogyi, Florian Götzinger, Angelika Berger, Roland Berger, Volker Strenger, Daniela S. Kohlfürst, Anna Zschocke, Benoît Bernar; Burkhard Simma, Edda Haberlandt, Christina Thir, Ariane Biebl, Christelle Christiaens, Marine Creuven, Koen Vanden Driessche, Tine Boiy, Daan Van Brusselen, An Bael, Sara Debulpaep, Petra Schelstraete, Natalia Gabrovska, Svetlana Velizarova, Nina Krajcar, Srđan Roglić, Ivan Pavić, Ulrikka Nygaard, Jonathan Peter Glenthoej, Lise Heilmann Jensen, Ilona Lind, Mihhail Tistsenko, Ülle Uustalu, Folke Brinkmann, Laura Buchtala, Stephanie Thee, Robin Kobbe, Cornelius Rau, Nicolaus Schwerk, Michael Barker, Maria Tsolia, Irini Eleftheriou, Patrick Gavin, Oksana Kozdoba, Borbàla Zsigmond, Laura Lancella, Francesca I Calò Carducci, Danilo Buonsenso, Piero Valentini, Andrea Lo Vecchio, Marcello Lanari, Luca Pierantoni, Inga Ivaškevičienė, Rimvydas Ivaškevičius, Valentina Vilc, Elisabeth Schölvinck, Astrid Rojahn, Anastasios Smyrnaios, Claus Klingenberg, Isabel Carvalho, Andreia Ribeiro, Anna Starshinova, Ivan Solovic, Petra Prunk, Veronika Osterman, Uros Krivec, Begoña Santiago-Garcia, Mar Santos, Antoni Noguera-Julian, Miguel Lanaspa, Antoni Soriano Arandes, Susana Melendo, Lola Falcón, Olaf Neth, Mario Pérez-Butragueño, Laura Minguell, Matilde Bustillo, Aida María Gutiérrez-Sánchez, Borja Guarch Ibáñez, Dr. Josep Trueta, Francesc Ripoll, Beatriz Soto, Karsten Kötz, Noémie Wagner, Arnaud G. L'Huillier, Ulrich Heininger, Hanna Schmid, Nicole Ritz, Petra Zimmermann, Franziska Zucol, Anita Niederer, Michael Buettcher, Benhur Sirvan Cetin, Olga Bilogortseva, Vera Chechenyeva, Marc Tebruegge, Delane Shingadia, Alicia Demirjian, Srini Bandi, Steven B Welch, Fiona Shackley, Lynne McFetridge, Lynne Speirs, Conor Doherty, Laura Jones, Paddy McMaster, Clare Murray, Frances Child, Yvonne Beuvink, Nick Makwana, Elizabeth Whittaker, Amanda Williams, Katy Fidler, Jolanta Bernatoniene, Rinn Song, Zoe Oliver, Andrew Riordan, Alasdair Bamford, Julia Kenny, Myrsini Kaforou, Christime E. Jones, Priyen Shah, Padmanabhan Ramnarayan, Alain Fraisse, Owen Miller, Patrick Davies, Filip Kucera, Joe Brierley, Marilyn McDougall, Michael Carter, Adriana Tremoulet, Chisato Shimizu, Jethro Herberg, Jane C Burns, Hermione Lyall, Michael Levin, Roopa Marcello, Johanna Dolle, Sheila Grami, Richard Adule, Zeyu Li, Kathleen Tatem, Chinyere Anyaogu, Stephen Apfelroth, Raji Ayinla, Noella Boma, Terence Brady, Braulio F. Cosme-Thormann, Roseann Costarella, Kenra Ford, Kecia Gaither, Jessica Jacobson, Marc Kanter, Stuart Kessler, Ross B. Kristal, Joseph J. Lieber, Vikramjit Mukherjee, Vincent Rizzo, Jr., Madden Rowell, David Stevens, Elana Sydney, Andrew Wallach, Dave A. Chokshi, Nichola Davis, Alvaro Moreira, Kevin Chorath, Karthik Rajesekaran, Fiona Burmeister, Mubbasheer Ahmed, Axel Moreira, Safiya Richardson, Jamie S Hirsch, Mangala Narasimhan, James M Crawford, Thomas McGinn, Karina W Davidson, Maraiawy Riollano-Cruz, Esra Akkoyun, Eudos Briceno-Brito, Shanna Kowalsky, Roberto Posada, Emilia Mia Sordillo, Michael Tosi, Rebecca Trachtman, Alberto Paniz-Mondolfi, Sergey Rekhtman, Rachel tennenbaum, Andrew Strunk, Morgan Birabaharan, Shari Wright, Amit Garg, AR Araujo da Silva, CGB Fonseca, JLPS Miranda, BV Travassos, CR Baião, KD Silva, LBAE dos Santos, MMR de Britto, PALS Cerqueira, SNB Pereira, RBJ Rios, CS Vieira, IA Leal, NC Martins, LMAC dr Carvalho, AB Pereira, CH Teixeira, Braian LA Sousa, Magda Sampaio-Carneiro, Werther B de Carvalho,

Clovis A Silva, Alexandre A Ferraro, CE Bolaños-Almeida, OM Espitia Segura, Héctor Cairoli, Silvana Riden, María J Chiolo, Sandra Di Lalla, Fernando Ferrero, Jb Vitoeereo, Edgardo Checacci, Carolina Davenport, Paula Dominguez, Horacio Planovsky, Fabian Gambarruta, Mariano Ibarra, Maria Fabiana Ossorio, Javier Potasnik, Norma Schenone, Milagros, Torrents and Fernando Torres.
